# Menopause modulates the circulating metabolome: evidence from a prospective cohort study

**DOI:** 10.1101/2021.12.17.21266891

**Authors:** Jari E. Karppinen, Timo Törmäkangas, Urho M. Kujala, Sarianna Sipilä, Jari Laukkanen, Pauliina Aukee, Vuokko Kovanen, Eija K. Laakkonen

## Abstract

**Aims:** We studied the changes in the circulating metabolome and their relation to the menopausal hormonal shift in 17β-oestradiol and follicle-stimulating hormone levels among women transitioning from perimenopause to early postmenopause.

**Methods and Results:** We analysed longitudinal data from 218 Finnish women, 35 of whom started menopausal hormone therapy during the study. The menopausal transition was monitored with menstrual diaries and serum hormone measurements. The median follow-up was 14 months (interquartile range: 8–20). Serum metabolites were quantified with targeted nuclear magnetic resonance metabolomics. The model results were adjusted for age, follow-up duration, education, lifestyle, and multiple comparisons. Menopause was associated with 84 metabolite measures. The concentration of apoB (0.17 standard deviation [SD], 99.5% confidence interval [CI] 0.03–0.31), VLDL triglycerides (0.25 SD, CI 0.05–0.45) and particles (0.21 SD, CI 0.05–0.36), LDL cholesterol (0.17 SD, CI 0.01–0.34) and particles (0.17 SD, CI 0.03–0.31), HDL triglycerides (0.24 SD, CI 0.02–0.46), glycerol (0.32 SD, CI 0.07–0.58) and leucine increased (0.25 SD, CI 0.02–0.49). Citrate (−0.36 SD, CI -0.57 to - 0.14) and 3-hydroxybutyrate concentrations decreased (−0.46 SD, CI -0.75 to -0.17). Most metabolite changes were associated with the menopausal hormonal shift. This explained 10% and 9% of the LDL cholesterol and particle concentration increase, respectively. Menopausal hormone therapy was associated with increased medium-to-large HDL particle count and decreased small-to-medium LDL particle and glycine concentration.

**Conclusions:** Menopause is associated with proatherogenic circulating metabolome alterations. Female sex hormones levels are connected to the alterations, highlighting their impact on women’s cardiovascular health.

**‘One-sentence Summary’:** Female sex hormone alterations induced by menopause altered the levels of circulating metabolites, leading to a higher risk profile for cardiovascular diseases.

## Introduction

Most women face menopause at the age of 48–52 years,^1^ resulting from the cessation of ovarian follicular activity and diagnosed 12 months after the final menstrual period.^2^ The characteristic menopausal hormonal shift consists of a decline in 17β-oestradiol (E2) and a concomitant increase in follicle-stimulating hormone (FSH) levels.^1^ Oestrogen-containing menopausal hormone therapy (MHT) alleviates menopausal symptoms and restores systemic E2 without lowering FSH to premenopausal levels.^3^

Menopause is thought to predispose women to atherosclerotic cardiovascular disease (ACVD) since they develop obstructive coronary artery disease 7–10 years later than men, and this risk rises after menopause.^4^ Also, premature menopause is associated with an increased ACVD risk.^5^ The causality between these phenomena is difficult to prove, as the menopause-driven metabolic changes may predispose women to present ACVD at an older age, making it difficult to distinguish their effects from other ageing-related changes.^6^ Moreover, as it is generally not possible to predict the timing of menopause, establishing appropriate longitudinal study settings is challenging. Perimenopause (the menopausal transition phase) also differs in timing and duration among individuals.^1^ Therefore, individual information on menopause progression benefits the investigation of menopausal effects.

Research on clinical biomarkers supports the relationship between menopause and ACVD risk. In longitudinal studies, menopause is associated with increases in circulating triglyceride and low-density lipoprotein (LDL) cholesterol levels. However, the effect of menopause on high-density lipoprotein (HDL) cholesterol concentration and its direction is controversial.^7–10^ The biomarker changes may result directly from the menopausal hormonal shift or indirectly via increased adiposity.^11^ The role of female sex hormones is strengthened by studies on MHT in postmenopausal women. Oestrogen-only MHT lowers LDL cholesterol and increases HDL cholesterol concentrations accompanied by a rise in triglyceride levels when administered orally.^12^ In combination MHT, the effects on HDL cholesterol are modulated by selected progestogen.^12^ MHT may also improve blood glucose regulation,^13^ reflecting broad systemic effects on metabolism.

Compared with clinical methods, metabolomics offer a wider lens to investigate menopausal effects on the circulating metabolome. Two population-level nuclear magnetic resonance (NMR) metabolomics studies on menopause have been conducted. The cross-sectional study by Auro et al.^15^ examined associations between age, sex, and menopause and the circulating metabolome in 26 065 Finnish and Estonian individuals, including 10 083 women. Later, Wang et al.^16^ investigated cross-sectional associations in the UK among 3 312 women, with 1 492 longitudinal samples taken 2.5 years apart. Their results were similar; menopause was associated with a proatherogenic shift in lipoprotein measurements and non-lipid metabolites, such as amino acids.^15,16^ Both studies relied on self-reported menopausal status and did not associate the findings to the female sex hormone levels.

Therefore, here, we investigated whether the menopause-related hormonal shift modulates the circulating metabolome in a longitudinal design, where the follow-up of the menopausal transition progression was individualised and monitored with repeated FSH level measurements. The premise was that menopause has an identifiable metabolomic fingerprint resulting from the shift of female sex hormone levels.

## Methods

### Study design and participants

This study used data from the Estrogenic Regulation of Muscle Apoptosis (ERMA) prospective cohort study^17^ and was approved by the ethics committee of the Central Finland Health Care District (KSSHP Dnro 8U/2014). The study complied with the Declaration of Helsinki, and participants gave informed consent.

The recruitment and the first measurements were performed between 2014 and 2015. The follow-up data were collected between 2015 and 2019. The sample was randomly drawn from the Population Information System. An invitation and a prequestionnaire were sent to 6 878 women aged 47–55 living in the Jyväskylä area. The response rate was 47%. Based on the prequestionnaire data, 1 627 women were invited to menopausal status determination. Exclusion criteria were self-reported body mass index > 35 kg/m^2^ and medical conditions or use of medication affecting the ovaries, the hormone or inflammatory profile, or daily functioning. The menopausal status of 1 393 women was determined, and 1 158 women were called to the Health and Sports Laboratory of the University of Jyväskylä for physiological and psychological measurements.

The ERMA study aimed to create a cohort for the investigation of menopausal effects with a minimum influence of ageing. Therefore, 381 perimenopausal women were invited to participate in the longitudinal Core-ERMA study (**Fig. 1**). Women kept a menstrual diary during follow-up and visited the laboratory for the measurement of FSH levels every 3 or 6 months until early postmenopause. The follow-up was scheduled according to the participant’s menopausal transition progression.

**Figure 1.**
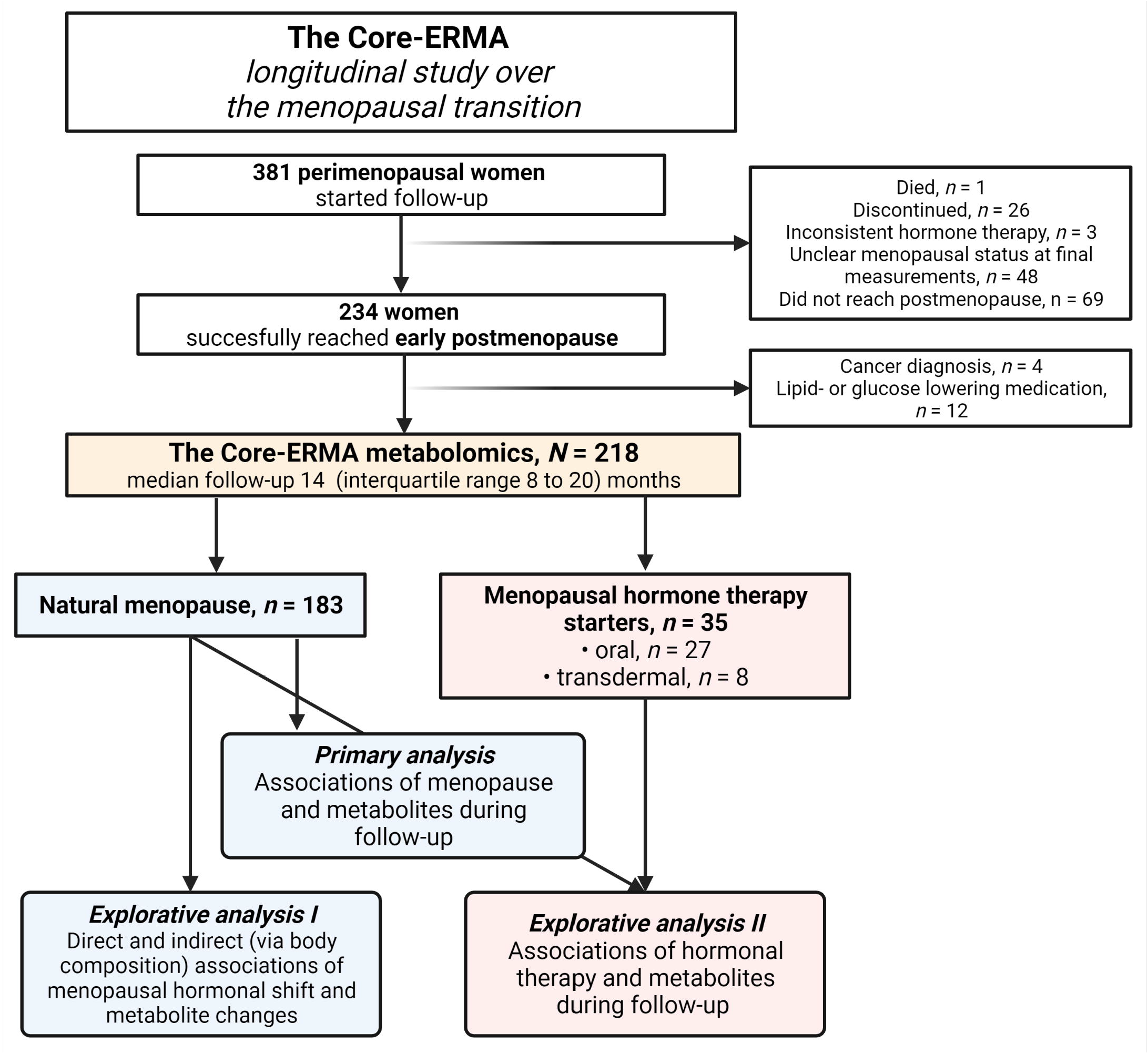
Study flowchart and statistical analysis approach.

Of the 381 participants, the follow-up was completed by 234 women. One participant died, and 26 dropped out. Three women reported inconsistent MHT use and 117 women did not reach postmenopause or their menopausal status was uncertain at the last measurement, leading to exclusion. From this study, we excluded 12 women using lipid or glucose-lowering medication and 4 women diagnosed with cancer. Therefore, the metabolomics analyses were performed for 218 participants, with 35 (15%) starting MHT during follow-up.

### Hormone and metabolites profile

Blood samples were collected between 7–10 AM after overnight fasting and processed for serum collection with a standard procedure. The samples were aliquoted and stored at –80 °C until analysis. E2 and FSH levels were measured with IMMULITE 2000 XPi (Siemens Healthcare Diagnostics, UK). Metabolites were analysed with a targeted proton nuclear magnetic resonance (^1^H-NMR) spectroscopy platform (Nightingale Health Ltd., Helsinki, Finland; biomarker quantification version 2020).^18^ The platform quantifies 250 metabolite measures and 180 were selected as outcomes. The lipoprotein lipid ratios were not included due to the limited sample size and because they were unlikely to provide substantial added value.

### Menopausal status and menopausal hormone therapy use

The guidelines Stages of Reproductive Aging Workshop (STRAW) + 10 with slight modifications were used to determine the menopausal status.^2^ Women kept a menstrual diary for 3 months before the first blood sampling. Perimenopausal women were required to have irregular or no menstrual bleeding and FSH levels of 17–30 IU/l. During follow-up, women were determined postmenopausal after two consecutive FSH measurements of > 30 IU/l and at least 6 months of amenorrhea. However, the absence of menses was slightly shorter in 16 participants. The criterion differs from the guidelines where postmenopause begins after 12 months of amenorrhea due to practical limitations. We decided to compromise the length of the follow-up period to maximise the number of women with valid end measurements inside a limited project funding period. The median amenorrhea duration was 8.1 months (interquartile range: 6.5–10.4). The postmenopausal status of 25 participants was determined solely based on FSH levels because of prior hysterectomy (*n* = 11) or ambiguity in menstrual diary reporting (*n* = 14).

MHT was queried at each follow-up meeting, and MHT participants were invited to the last measurement 6 months after the initiation of the treatment to allow the medication to exert its effects. One woman reported starting MHT 17 days before and another participant 2 days before the last measurement. We only classified the former as an MHT user because a 2-day use was unlikely to substantially affect metabolism. Her metabolite levels did not significantly differ from the other participants’ levels. Of the 35 MHT starters, 27 used oral products containing either E2-only (*n* = 7), E2 in combination with dydrogesterone (*n* = 16) or norethisterone acetate (*n* = 4). Concerning the E2-only MHT starters, 5 were users of levonorgestrel-releasing intrauterine devices, and 2 did not need progestogen because of past hysterectomy. The rest of the sample (*n* = 8) used transdermal MHT, 7 of whom used E2-gel with a levonorgestrel-releasing intrauterine device and one E2 and norethisterone acetate patches.

### Education level, lifestyle factors and body composition

**The Method Supplement** includes a detailed description of the study covariates. Briefly, the education level (primary, secondary or tertiary) and lifestyle factors were recorded with structured questionnaires. Due to the low number of participants with primary education, primary and secondary were combined in the statistical analyses. Smoking status was classified as ‘never’, ‘quitter’, or ‘current smoker’. As the number of current smokers was low, dummy variables were created to describe whether the participant had ever smoked and smoked currently. Alcohol use was calculated as portions per week. Physical activity was calculated as metabolic equivalent hours per day (MET-h/d) by assessing the duration, frequency and intensity of leisure-time physical activity and the time spent on active commuting.^19^ Use of common foods in Finnish food culture were recorded with a 45-item food-frequency questionnaire and the diet quality was measured with an 11-element diet quality sum score (DQS) adapted from a validated tool,^20^ where a higher score reflects a healthier diet. Height was measured with a stadiometer at the first measurement. Body mass and body fat percentage were measured with InBody720 (Biospace, Seoul, Korea).

### Statistical analyses

The statistical analyses are described in detail in **the Method Supplement**. Questionnaire-based data were missing from 1 participant at the first and 2 participants at the last measurement. We completed the missing values by inspecting participants’ answers from other visits or mean imputation. Metabolite data were nearly complete, and we did not impute the rare missing values. R version 4.0.0 or newer was used for statistical analyses unless stated otherwise.

The primary results are the associations between menopause and metabolite measurements in 183 women experiencing natural menopause, i.e., who did not start MHT during follow-up (**Fig. 1**). Analyses were performed using linear mixed-effect models with random intercept after metabolite Box-Cox transformation and standardisation with respect to the first measurements. The metabolites were treated as outcomes and the menopausal status as exposure. Two adjusted models were built in addition to a crude model. The first adjusted model (the main study results) included the covariates of age at the first measurement, follow-up duration, education level and lifestyle factors. In the second adjusted model, the body fat percentage (potential mediator between menopause and metabolites) was included as a covariate. The K_eff_-Šidák correction was used to account for multiple testing.

Two exploratory analyses were performed. First, the direct and indirect associations (via body fat percentage change) between the menopausal hormonal shift and metabolite changes in the women experiencing natural menopause (*n* = 183) were investigated using latent change score modelling. Both E2 and FSH were included in the model simultaneously because this combination better characterises an individual’s sex hormone profile as E2 levels fluctuate during perimenopause. The used model (**Method Supplement Fig. 1**) can be seen as an extension of the paired *t*-test, where change is controlled for metabolite concentrations at the first measurement. The calculated effect sizes are interpreted similarly to squared semi-partial correlations and indicate how much of the metabolite change is explained by the menopausal hormonal shift. Mplus version 7.4 was used to estimate the model parameters. False discovery rate adjustment was used to correct for multiple testing. The association of MHT initiation during follow-up with the metabolite measurements was studied in the whole sample (*N* = 218, *N*_MHT starters_ = 35) using menopausal status and MHT interaction as the exposure. The model structures and multiple testing corrections were performed as in the primary analysis.

## Results

Participant characteristics can be found in **Table 1**. The mean age of participants was 51.7 (SD = 1.9) at the first measurement. Hypertension was the most commonly diagnosed condition in the cohort. Participants did not use lipid-lowering agents, even though more than half had elevated LDL cholesterol levels (> 3 mmol/l). MHT starters were younger than non-users at the first measurement. Moreover, they had lower FSH levels and systolic/diastolic blood pressure. As for the rest of the variables, the groups showed similar characteristics at the beginning of the study.

**Table 1.** Characteristics of the participants at the first and last measurements.

|  | Natural menopause (*n* = 183) |  | MHT starters (*n* = 35) |  |
| --- | --- | --- | --- | --- |
|  | Perimenopause | Postmenopause | Perimenopause | Postmenopause |
| **Age** (years) | 51.9 (1.9) | 53.1 (1.9) | 50.6 (1.8) | 52.0 (1.9) |
| **Follow-up** (days) | - | 397 (243–603) | - | 464 (334–707) |
| **HT use** (days) | - | - | - | 220 (191–242) |
| **Sex hormones** |  |  |  |  |
| E2 (nmol/l) | 0.25 (0.17–0.41) | 0.16 (0.11–0.27) | 0.25 (0.18–0.42) | 0.28 (0.20–0.61) |
| FSH (IU/l) | 30.0 (23.4–48.5) | 69.8 (46.8–89.7) | 23.0 (16.5–36.1) | 36.1 (18.4–60.0) |
| **Education** |  |  |  |  |
| Primary | 5 (3%) | 5 (3%) | 1 (3%) | 1 (3%) |
| Secondary | 100 (55%) | 100 (55%) | 15 (43%) | 15 (43%) |
| Tertiary | 78 (42%) | 78 (43%) | 19 (54%) | 19 (54%) |
| **Body composition** |  |  |  |  |
| Height (cm) | 165.1 (5.7) | - | 166.3 (5.5) | - |
| Body mass (kg) | 69.5 (11.3) | 70.2 (11.6) | 70.4 (10.2) | 70.5 (10.8) |
| Body mass index categories |  |  |  |  |
| Healthy weight (< 25 kg/m<sup>2</sup>) | 92 (50%) | 85 (46%) | 20 (57%) | 19 (54%) |
| Overweight (25–29.9 kg/m<sup>2</sup>) | 66 (36%) | 71 (39%) | 10 (29%) | 12 (34%) |
| Obesity (≥ 30 kg/m<sup>2</sup>) | 25 (14%) | 27 (15%) | 5 (14%) | 4 (11%) |
| Percent body fat (%) | 30.9 (8.3) | 32.1 (8.0) | 30.5 (7.1) | 31.4 (6.3) |
| **Smoking** |  |  |  |  |
| Never | 122 (67%) | 122 (67%) | 26 (74%) | 26 (74%) |
| Quitter | 49 (27%) | 50 (28%) | 6 (17%) | 5 (14%) |
| Smoker | 12 (7%) | 11 (6%) | 3 (9%) | 4 (11%) |
| **Alcohol use** (portions/week) | 3.0 (1.0–5.0) | 2.5 (1.0–5.0) | 4.5 (2.5–5.8) | 2.5 (1.0–4.9) |
| **Diet quality score** (0–11) | 5.7 (2.3) | 5.5 (2.1) | 5.5 (2.4) | 5.6 (2.4) |
| **Physical activity** (MET-h/d) | 3.6 (1.7–4.8) | 3.8 (2.3–5.6) | 4.5 (1.8–7.5) | 4.8 (2.5–7.5) |
| **Blood pressure** |  |  |  |  |
| Diastolic (mmHg) | 85 (10) | 82 (10) | 80 (9) | 77 (11) |
| Systolic (mmHg) | 134 (19) | 131 (18) | 126 (16) | 121 (14) |
| Diagnosed hypertension | 24 (13%) | 29 (16%) | 4 (11%) | 4 (11%) |
| ATC1 class C medication | 25 (14%) | 34 (19%) | 5 (14%) | 6 (17%) |
| **Dyslipidaemia and blood glucose** |  |  |  |  |
| LDL cholesterol > 3 mmol/l <sup>a</sup> | 99 (54%) | 119 (65%) | 22 (62%) | 20 (57%) |
| HDL cholesterol < 1.2 mmol/l | 5 (3%) | 4 (2%) | 1 (3%) | 0 (0%) |
| Triglycerides |  |  |  |  |
| Optimal (< 1.2 mmol/l) | 123 (67%) | 107 (58%) | 24 (69%) | 24 (69%) |
| Elevated (> 1.7 mmol/l) | 22 (12%) | 27 (15%) | 1 (3%) | 5 (14%) |
| Fasting glucose > 6 mmol/l | 8 (4%) | 12 (7%) | 2 (6%) | 3 (9%) |

Data as means (standard deviation), medians (interquartile range) or counts (%).
<sup>a</sup> Calculated with Friedewald formula.
E2, 17β-oestradiol; FSH, follicle-stimulating hormone; MET, metabolic equivalent of task; MHT, menopausal hormone therapy.

The median follow-up duration was 14 months (interquartile range: 8–15), ranging from 4 months to 3.5 years. Expectedly, E2 decreased and FSH increased in women not using MHT during follow-up. Among MHT starters, E2 increased without an apparent change in FSH levels. As for lifestyle factors, alcohol use decreased in both groups and body fat percentage increased by 1% for the total sample with a similar trend in both groups.

### Menopause and metabolite associations

In **Results Supplement Table 1**, the absolute metabolite concentrations during the study and the unstandardised metabolite change scores of the 183 women experiencing natural menopause are shown. Menopause was associated with a statistically significant change in 85 metabolite measures (**Result Supplement Table 3**; the key findings are summarised in **Fig. 2**). The apolipoprotein B (apoB)-containing particle count increased by 0.17 SD (99.95% confidence interval [CI] 0.03–0.31), resulting from the increased very low-density lipoprotein (VLDL; 0.21 SD, CI 0.05–0.36) and LDL particle counts (0.17 SD, CI 0.03–0.31). The VLDL increase favoured the increase of VLDL particle size (0.22 SD, CI 0.03–0.41). The apolipoprotein A-I (apoA-I) and total HDL particle counts did not change markedly. However, it is uncertain whether small HDL subclass is affected by menopause (0.19 SD, CI -0.01 to 0.40). From the lipid measurements, cholesterol concentrations in all apoB-containing lipoprotein classes increased from 0.17 to 0.20 SD. The increase in VLDL triglyceride (0.25 SD, CI 0.05–0.45) and HDL triglyceride concentrations (0.24 SD, CI 0.02– 0.46) was even more pronounced. Total serum fatty acid concentration did not change but the fatty acid profile shifted from polyunsaturated to saturated direction. As for the other metabolites, citrate concentration decreased by -0.36 SD (CI -0.57 to -0.14) and glycerol concentration increased by 0.32 SD (CI 0.07–0.58). Glutamine concentration decreased by - 0.46 SD (CI -0.72 to -0.20) and leucine concentration increased by 0.25 SD (CI 0.02–0.49). Acetoacetate and 3-hydroxybutyrate levels decreased by -0.44 SD (CI −0.72 to −0.15) and -0.46 (CI −0.75 to −0.17), respectively. When body fat percentage was included as a covariate, the association sizes of lipoprotein, lipid, glycerol and amino acid concentrations were roughly 0.03–0.05 SD smaller (**Result Supplement Table 3**). The adjustment had a negligible effect on the associations of menopause with citrate and ketone levels.

**Figure 2.**
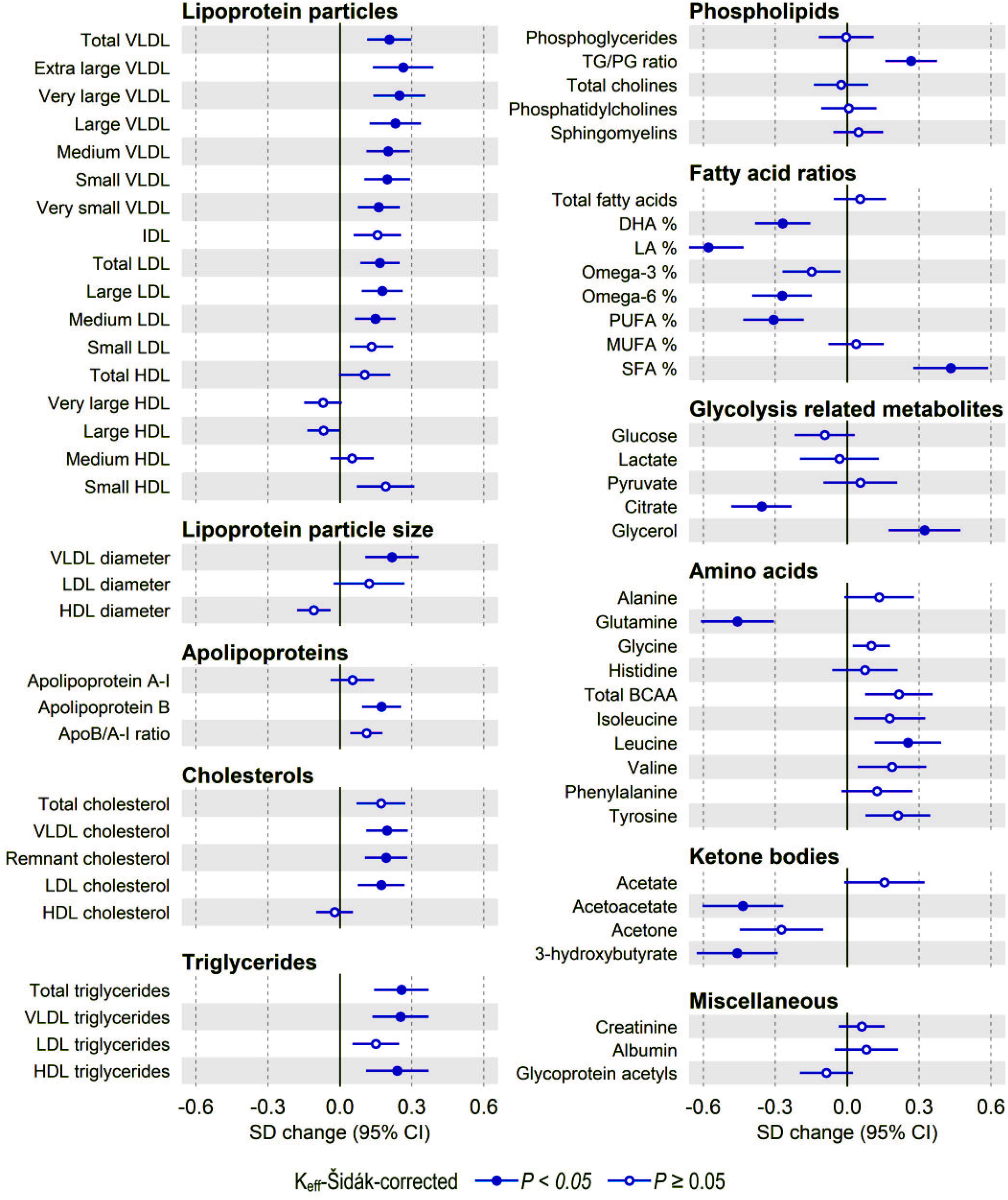
The associations of the menopausal transition and key metabolite measurements in women experiencing natural menopause (*n* = 183) adjusted for age at the first measurement, follow-up duration, education level, smoking status, alcohol use, physical activity and diet quality. The figure describes the standardised change (expressed as standard deviation [SD] units) in the metabolite levels during follow-up with respect to the first measurement values, allowing the comparison between metabolite measures with different units and concentrations.

### Menopausal hormonal shift and metabolite changes

The menopausal hormonal shift directly explained the change in 64 of the 85 metabolites identified as menopause-responsive in the primary analysis, with effect sizes ranging from 2.1% to 11.2% (**Result Supplement Table 4**). Based on the size of the standardised regression estimates of the two hormones included in the model, the decline of E2 levels had more impact on the results than the increased FSH levels. The hormonal shift was not associated with indirect metabolite changes since the shift was not associated with body fat percentage change.

The key findings are summarised in **Fig. 3**. The menopausal hormonal shift directly explained the concentration increase in apoB (7.4%), LDL particles (8.5%) and LDL cholesterol (10.6%). The effect sizes concerning the total VLDL particle count (4.0%) and triglyceride levels (2.9%) were more modest. Even though the association of menopause and the increased small HDL particle count was inconclusive in the primary analysis, the exploratory analysis strongly linked them with an effect size of 10.9%.

**Figure 3.**
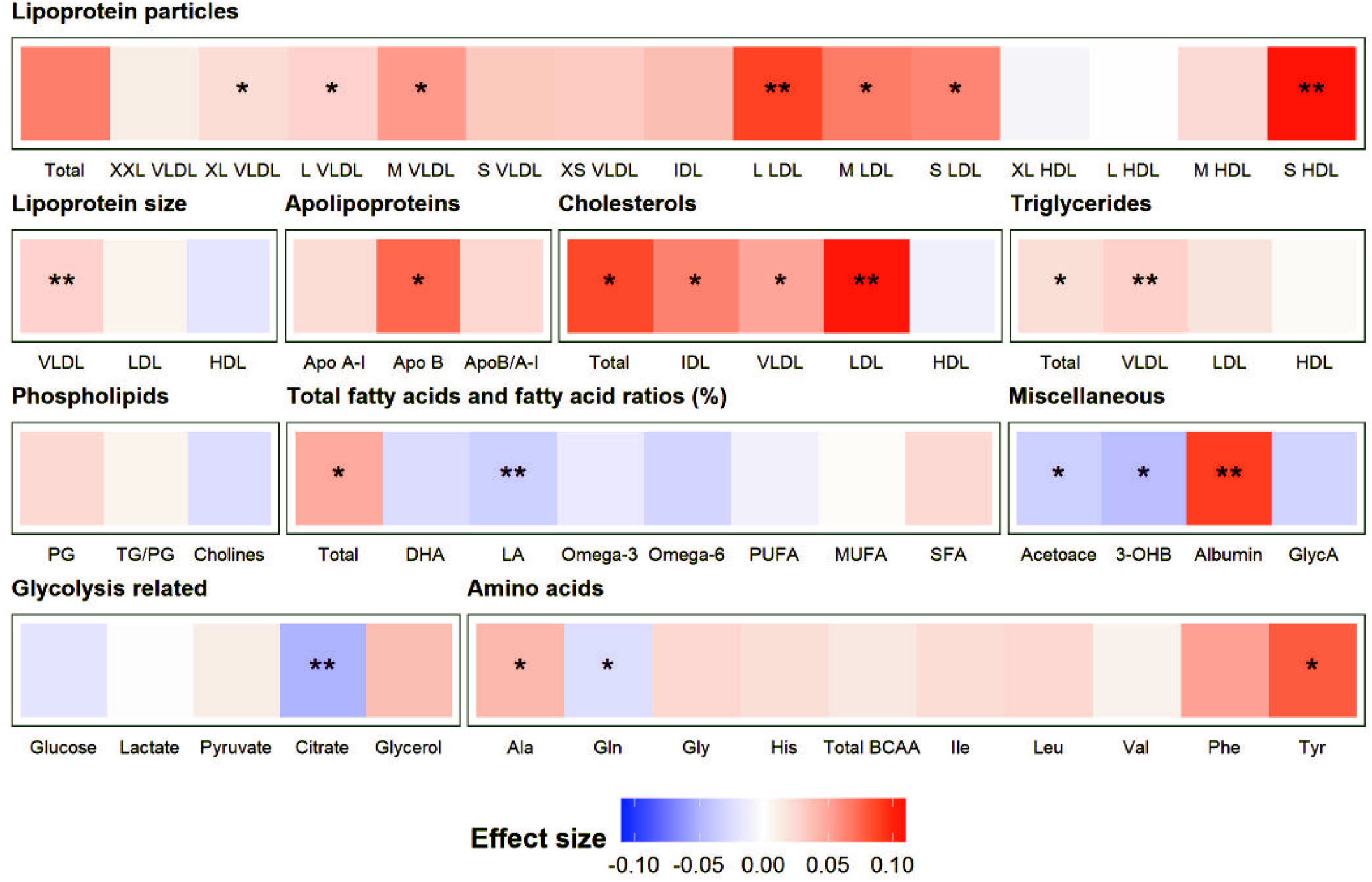
The associations of menopausal hormonal change (the decline in E2 and increase in FSH levels) with key metabolite changes. The colours and their intensity show the association direction and the effect size. The models were adjusted for age at the first measurement, follow-up duration, education level, smoking status, alcohol use, physical activity and diet quality. False discovery corrected *P*-values: * − 0.05; ** − 0.01. 3-OHB, 3-hydroxybutyrate; Ala, alanine; Gln, glutamine; Gly, glycine; His, histidine; Ile, isoleucine; Leu, leucine; Val, valine; Phe, phenylalanine; Tyr, tyrosine.

From the results concerning the association of non-lipoprotein and lipid with menopause in the primary analysis, a direct association with the hormonal shift was confirmed for glutamine (2.4%), citrate (5.0%), 3-hydroxybutyrate (4.4%) and acetoacetate (3.4%) concentrations. The largest effect size of the hormonal shift was found for tyrosine concentration (7.8%), although menopause and tyrosine level association did not remain statistically significant in the primary analysis after multiple comparison corrections.

### MHT and metabolite changes

In **Results Supplement Table 2**, the absolute metabolite concentrations for MHT starters during the study including the unstandardised change scores are presented. MHT initiation during follow-up was characterised by a decreased apoB/apoA-I ratio (**Result Supplement Table 5**; the key statistically significant findings are summarised in **Table 2**), resulting from the increased particle count in medium and large HDL subclasses and the decreased small and medium LDL subclass particle counts. VLDL was less affected by MHT than LDL and HDL. As for non-lipid metabolites, we found an inverse association between MHT and glycine concentration. Adjusting for body fat percentage did not influence these associations.

**Table 2.** Menopause and menopausal hormone therapy interactions with key metabolite measurements.

| Metabolite | Crude model |  |  | Adjusted model<sup>a</sup> |  |  |
| --- | --- | --- | --- | --- | --- | --- |
|  | Interaction | 99.95% CI | *P* | Interaction | 99.95% CI | *P* |
| ApoB/apoA-I ratio | -0.54 | -0.86 to -0.21 | < 0.001 | -0.55 | -0.88 to -0.22 | < 0.001 |
| ApoA-I | 0.51 | 0.08 to 0.93 | 0.005 | 0.55 | 0.13 to 0.97 | 0.001 |
| Glycine | -0.46 | -0.80 to -0.11 | 0.001 | -0.46 | -0.80 to -0.12 | 0.001 |
| Medium HDL particles | 0.54 | 0.10 to 0.97 | 0.003 | 0.58 | 0.15 to 1.01 | 0.001 |
| Large HDL particles | 0.35 | 0.06 to 0.64 | 0.004 | 0.36 | 0.07 to 0.66 | 0.002 |
| HDL cholesterol | 0.36 | 0.02 to 0.70 | 0.028 | 0.39 | 0.05 to 0.74 | 0.008 |
| HDL diameter | 0.35 | 0.05 to 0.65 | 0.007 | 0.35 | 0.04 to 0.74 | 0.009 |
| Small LDL particles | -0.41 | -0.76 to -0.05 | 0.005 | -0.44 | -0.83 to -0.05 | 0.011 |
| LDL particles | 0.31 | -0.76 to -0.05 | 0.010 | -0.39 | -0.75 to -0.03 | 0.020 |
| Medium LDL particles | -0.39 | -0.74 to -0.03 | 0.017 | -0.37 | -0.77 to -0.01 | 0.039 |

<sup>a</sup> age at the first measurement, follow-up duration, education level, and lifestyle factors
Apo, apolipoprotein; HDL, high-density lipoprotein; LDL, low-density lipoprotein
The confidence intervals and P-values were Keff-Šidák-corrected for multiple comparisons.

## Discussion

This study investigated the associations of menopause and circulating metabolome in 183 women transitioning from perimenopause to early postmenopause. The study also explored whether the menopausal hormonal shift explains the observed changes and whether MHT initiated during the follow-up (*n* = 35) was associated with metabolite changes. The results showed that menopause-induced hormonal shift is associated with a proatherogenic metabolomic fingerprint. MHT specifically influences LDL, HDL and glycine metabolism. These findings broadly agree with earlier metabolomics studies on menopause,^15,16^ and now connect the previous and present observations to the female sex hormone levels.

### Menopause modulates lipoprotein and lipid metabolism towards a proatherogenic profile

ApoB-containing lipoproteins cause ACVD,^21^ with LDL being the primary disease driver.^22^ Studies using both clinical methods and metabolomics have identified LDL as one of the most menopause-responsive biomarkers.^8,9,15,16^ In agreement with this, we report increased apoB and LDL cholesterol and particle concentration, 7–10% of which are directly explained by the menopausal hormonal shift. The underlying mechanism is probably the well-established oestrogen-mediated LDL receptor modulation, ^23,24^ influencing LDL clearance from the circulation.^25^ However, similar to Auro et al.^15^ but contrary to Wang et al., ^16^ menopause did not alter LDL size distribution in our study. MHT was associated with decreased particle counts in medium and small LDL subclasses, providing further evidence that female sex hormones regulate LDL metabolism. These observations support the clinical guidelines to initiate MHT early into menopause as this timing offers the greatest cardioprotective benefits.^4^

Triglyceride-rich lipoproteins also contribute to the ACVD risk.^26^ Menopause was associated with an increased VLDL particle count, favouring larger particles possibly in the range of VLDL1 size. Moreover, VLDL triglyceride and cholesterol concentrations increased. The observed pattern was similar to the results of Auro et al.^15^ but different from Wang et al.^16^ Our findings also agree with a recent study^27^ on the sex differences in plasma metabolites at multiple life stages that identified VLDL triglycerides as the most menopause-responsive lipid measurement.

The menopause and VLDL associations observed in this study were robust against body fat percentage adjustment. Still, only ?4% of the VLDL changes were directly explained by the menopausal hormonal shift. Therefore, the potential causal pathway between menopause and VLDL increase probably entails mediators like insulin resistance, as overproduction of VLDL particles and hypertriglyceridemia are hallmarks of chronically elevated insulin levels.^26^ VLDL was less affected by MHT than the LDL variables, possibly resulting from the effect on the liver caused by the use of oral products (the majority of MHT starters in this study).^25^

Unlike apoB-containing lipoproteins, total HDL particle and cholesterol concentrations are associated with reduced ACVD risk in observational studies, but their causal role is debatable.^28^ Consistent with previous metabolomic studies,^15,16^ the apoA-I count and the HDL particle or cholesterol concentrations did not decrease during follow-up. However, in our previous study,^7^ using data obtained with clinical immunoassay from the same serum samples, HDL cholesterol levels increased. HDL cholesterol increase was also observed during a 4-year follow-up in premenopausal women transitioning into postmenopause.^10^ This discrepancy could result from the different measurement methods.

However, based on the present metabolomics analyses, the effects of menopause on HDL metabolism do not favour cardiovascular health. First, menopause was associated with increased HDL triglyceride concentration that, contrary to HDL cholesterol levels, is associated with a higher risk of coronary heart disease.^29^ Furthermore, the menopausal hormonal shift was associated with a particle count increase in the small HDL subclass, whereas MHT was associated with a particle count increase in medium and large HDL subclasses. HDL subclasses relate differently to ACVD risk as only larger particles are protective.^30^ Moreover, HDL size distribution is linked to insulin resistance as smaller particles accumulate when glucose tolerance worsens.^31^ Our results agree with the longitudinal The Study of Women’s Health Across the Nation (SWAN) study, where menopause was associated with an increase in the small HDL particle and HDL triglyceride concentrations.^32^ In the future, examining menopause-associated structural and functional HDL changes will provide more insight into the phenomenon.

### Menopause is associated with a metabolic signature indicative of deteriorating insulin sensitivity

Our findings from other metabolite classes link menopause to a broader deterioration of cardiovascular and metabolic health. Blood glucose levels did not increase during follow-up. However, glucose homeostasis is tightly regulated, and insulin sensitivity changes may influence other metabolites more rapidly. For example, menopause was associated with increased glycerol and decreased 3-hydroxybutyrate and acetoacetate concentrations.

Glycerol levels are inversely associated with insulin sensitivity and predict type 2 diabetes.^33^ Conversely, ketone levels are positively associated with insulin sensitivity in nondiabetic subjects^34^ like our participants. The menopause-associated decrease in ketone production fits into the picture with the observed VLDL triglyceride increase, indicating that fatty acids are rather esterified into triglycerides than used for oxidation in the liver after menopause.

Moreover, we noticed that the serum fatty acid profile changed from polyunsaturated to saturated, relating to elevated type 2 diabetes risk.^35^ One explanation for this profile change is increased *de novo* lipogenesis.^36^ Insulin resistance increases hepatic fatty acid synthesis, upscaling VLDL production.^37^ Altered diet is another explanation. Based on the diet assessment, participants’ dietary habits were stable during the follow-up. However, this method provides only a crude estimation of the overall diet quality and does not capture possible changes in energy intake.

Furthermore, menopause was associated with amino acid changes, as previously reported.^15,16,27^ Leucine concentration increased during follow-up, and the menopausal hormonal shift was associated with increased tyrosine levels. Elevated aromatic amino acids and branched-chained amino acid concentrations follow insulin resistance^36,38,39^ and predict type 2 diabetes risk.^40^ Higher levels of branched-chained amino acids were associated with higher ACVD risk in women.^41^ Conversely, higher glycine and glutamine concentrations predict a decreased risk of type 2 diabetes.^36,40^ In line with our other findings, glutamine concentration decreased during follow-up, contradicting previous observations.^15,16^ Even though ovariectomy in rats did not alter skeletal muscle glutamine synthetase expression or activity,^42^ the glutamine concentration decrease may be due to the reduced branched-chained amino acid catabolism because the primary nitrogen acceptor, α-ketoglutarate, is initially converted to glutamate and, then, partially to glutamine.^43^ We did not find a statistically significant association between menopause and glycine levels. However, MHT was associated with decreased glycine concentration, similar to previous mass spectrometry studies using randomised controlled trial samples.^44,45^ Therefore, all MHT effects may not favour cardiovascular health.

### Menopause-associated changes in the circulating metabolome are linked to bone health but not increased inflammation

Oestrogen receptors exist throughout the body, with menopause causing broad systemic effects on multiple organ systems.^46^ The decreased citrate concentration during follow-up is a notable finding that links the circulating metabolome possibly to the skeletal system, where about 90% of body citrate resides.^47^ In mice, ovariectomy led to bone mineral density loss and decreased bone and plasma citrate levels.^48^ This decrease may result from reduced citrate synthesis due to osteoblast loss and increased citrate consumption by osteoclasts and lipid synthesis.^49^ As menopause relates to a bone mass decrease,^50^ serum citrate concentration could be a novel imaging-free biomarker of bone health in the population. An alternative explanation for the citrate association is that the observed change results from the potential deterioration of insulin sensitivity, as citrate levels were associated with insulin resistance among Finnish bariatric surgery patients.^51^ MHT did not influence citrate levels in this study.

Concerning the rest of the quantified metabolites, we found no or inconsistent evidence on their relationship with menopause. Contrary to Wang et al.,^16^ we did not observe a positive association between menopause or the menopausal hormonal shift and the inflammation marker GlycA. Moreover, we did not observe an inverse association between MHT and GlycA levels as Auro et al. did.^15^ Our findings indicate that at least the short observation time around the final menstrual period (the median follow-up was 14 months) do not reveal a sharp increase in systemic inflammation. Although menopause was previously associated with increased albumin concentration,^16^ and MHT users had lower albumin levels than nonusers,^15^ neither menopause nor MHT was significantly associated with albumin concentration in this study. Nevertheless, the menopausal hormonal shift was still related to increased albumin levels.

### Strengths and limitations

The key strengths of this study are the longitudinal design and the detailed menopausal transition monitoring by repeated hormone measurements. Moreover, we focused on changes occurring around the final menstrual period. Hence, we individualised the follow-up time for each woman to ensure they were at the same early postmenopausal state at the last measurement instead of using a standard follow-up time for all the participants.

The main study limitation is the relatively small sample size for a metabolomics study. This probably led to the detection of the most notable associations only. Additionally, the NMR metabolomics platform focuses on lipoprotein and lipid measurements and does not capture all ACVD-relevant metabolites, such as clotting factors. In the exploratory analysis, we also divided participants into MHT non-starters and starters *post hoc*. We recommend caution when interpreting the findings as women who start MHT may differ from women not requiring medical assistance with menopausal symptoms. Moreover, the whole-body fat percentage was considered the adiposity variable for indirect associations between menopausal hormonal shift and metabolite changes. The hormonal shift did not associate with the body fat percentage change and, therefore, the indirect link with metabolite changes was discarded. In this aspect, visceral fat mass quantification might have been more informative. Last, the generalisability of our findings is limited to healthy white women.

In conclusion, menopause is associated with a proatherogenic circulating metabolome change that relates to the induced hormonal shift and is partially modified by MHT. These findings highlight the impact of female sex hormones on women’s cardiovascular health.

## Supporting information

Result Supplement

## Data Availability

All data produced in the present study are available upon reasonable request to the authors

## Acknowledgements

We thank the ERMA women for their participation and the Faculty of Sport and Health Sciences staff for their contribution to the data collection.

## Data availability

The data underlying this article cannot be shared publicly for the privacy of the participants. The data will be shared on reasonable request to the corresponding author.

