## Supplementary material for "Menopause modulates the circulating metabolome: evidence from a prospective cohort study": Method Supplement

### 1. Study covariates

***Age*** was calculated as a specific age (e.g. 51.3 years) according to the participant's date of birth, drawn from the Finnish National Registry, and measurement date.

Education level and lifestyle factors were recorded with a structured questionnaire in the first and last measurements, separated by 4 months to 3 ½ years. The participants filled the questionnaires (exluding food-frequency questionnaires) at every follow-up meeting.

***Education level*** was assessed by a single question, including seven answer options. Based on the question, a three-category variable (primary, secondary and tertiary) was constructed. The primary level included women whose only education was a primary school. Secondary level included women who had a higher education than that, up to a bachelor level. Women who achieved a master's or doctoral degree were categorised as reaching the tertiary education level. As only 6 women had primary level education, primary and secondary levels were combined in the statistical analyses.

***Smoking status*** classification was based mainly on two questions. First, women were asked whether they had smoked more than 5 to 10 cigarettes in their lifetime. If the participant answered no, she was classified as a 'never smoker'. Participants who answered yes were then asked if they currently smoke cigarettes regularly. Based on their answer (yes or no), they were categorised as 'current smokers' or 'quitters'. Missing data on smoking were imputed based on responses in previous or later measurements. As the number of 'current smokers' was low, two dummy variables were created to describe whether the participant had 'ever smoked' and whether she 'smoked currently'.

***Alcohol use*** was calculated based on reported weekly consumption of 1) beer, cider, and long drinks, 2) wine or similar mild beverages, and 3) strong alcoholic drinks. The answer included seven answer options. Total alcoholic beverage use was calculated and expressed as portions per week.

***Self-reported leisure-time physical activity* (LTPA)** was recorded by a 4-item questionnaire.[1] First, the participants were asked how many times per month they currently participate in LTPA. The question had 6 answer options ranging from 'less than once per month' to 'over 20 times per month'. The average LTPA intensity was then queried by asking whether the typical LTPA-intensity was similar to 1) walking, 2) walking with periodical jogging, 3) jogging or 4) running. The third question asked the respondent to rate an LTPA session's average duration with five answer options ranging from 'under 15 minutes' to 'over two hours'. The last question asked how much time the participant spent daily on active commuting. The six answer options ranged from 'no active commuting' and 'I do not currently work' to 'an hour or longer'. Based on the first three questions, daily LTPA volume (MET-h/d) was calculated with an equation: (monthly LTPA frequency x minute duration x metabolic equivalent of task (MET) -intensity) / 30 days. The MET-intensities were derived from Ainsworth et al.[2] and recoded as walking = 4 METs, walking with periodical running = 6 METs, jogging = 8 METs and running = 13 METs. Next, the active commuting volume (MET-h/d) was calculated with an equation: (average daily active commuting duration x 4 MET-intensity x 5 times a week frequency) / 7 days. Finally, summing the leisure-time and active commuting physical activity volumes together, the total self-reported LTPA (MET-h/d) was calculated.

***Diet quality score (DQS)*** was calculated based on a 45-item food-frequency questionnaire, which the participants filled only at the first and last measurements. The food-frequency questionnaire included typical Finnish food culture food items, with 6 answer options. The Diet Quality Score (DQS) was adapted from an earlier validated tool.[3] As the used food-frequency questionnaires differed partly from each other, direct use of the original tool was impossible. The DQS consisted of 11 elements (**Method Supplement Table 1**) determined characteristics to a healthy diet by the Nordic Nutrition Recommendations 2012.[4] Each item was scored 1 point, and the scores were summed into the DQS. Therefore, the maximum DQS was 11 points. Higher DQS reflects a healthier diet.

**Method Supplement Table 1.** The Diet Quality Score elements and their background rationale.

| Item | Response | Score | Rationale |
|---|---|---|---|
| **1. Whole grain products**<br>Dark bread<br>(rye or crisp bread) | not every day (1–5)<br>once a day/more often (6) | 0<br>1 | Daily use of whole-grain bread is recommended to ensure adequate carbohydrate and fibre intake. |
| **2. Highly processed grain products**<br>a) White bread AND/OR<br>b) Baked goods | twice a week or more (4–6)<br>once a week or less (1–3) | 0<br>1 | Limited use of processed grain products is recommended due to high energy density and low nutritional value. |
| **3. Low-fat dairy products**<br>a) Sour milk or yoghurt OR<br>b) Low-fat cheese (< 20%) | not every day (1–5)<br>once a day/more often (6) | 0<br>1 | Daily use of low-fat dairy products is recommended to aid protein, calcium and vitamin D intake. |
| **4. Vegetables**<br>a) Cooked vegetables or legumes OR<br>b) Fresh vegetables, root vegetables or lettuce | not every day (1–5)<br>once a day/more often (6) | 0<br>1 | Daily use of vegetables is recommended to ensure fibre and nutrient intake. The advice is consistent across various dietary guidelines |
| **5. Fruits and berries**<br>a) Fruits OR<br>b) Fresh or frozen berries | not every day (1–5)<br>once a day/more often (6) | 0<br>1 | Daily use of fruits and berries is recommended to ensure fibre and nutrient intake. |
| **6. Fish**<br>Fish or fish foods | once a week or less (1–3)<br>at least twice a week (4–6) | 0<br>1 | Eating fish 2–3 times per week is recommended to ensure adequate intake of omega-3 fatty acids. |
| **7. Processed meats**<br>a) Sausage foods or hot dogs AND/OR<br>b) Cured meat | once a week or more often (3–6)<br>only once or twice a month (1–2) | 0<br>1 | Consumption of processed meat should be limited due to high amounts of saturated fat and sodium. |
| **8. Sugar-sweetened beverages**<br>Sugar-sweetened juices or soft drinks | once a month or more often (2–6)<br>less than once a month or never (1) | 0<br>1 | Consumption of sugar-sweetened beverages should be limited to a minimum as they are energy-dense and have low nutritional value. |
| **9. Polyunsaturated fats**<br>a) Nuts OR<br>b) Seeds | twice a week or less (1–4)<br>at least almost every day (5–6) | 0<br>1 | Nuts and seeds contain polyunsaturated fatty acids and other beneficial nutrients, and their daily intake is recommended. |
| **10. Fast food**<br>a) Pizza<br>b) Hamburgers<br>c) Fried potato products | once a week or more often (2–6)<br>only once a month (1–2) | | The intake of fast food should be limited to a minimum due to high energy density and high amounts of saturated fat and salt |
| **11. Sweet or salty snacks**<br>a) Chocolate AND/OR<br>b) Candy AND/OR<br>c) Salty snacks | twice per week or more often (4–6)<br>once per week at most (1–3) | 0<br>1 | The intake of processed snacks should be limited due to high energy density and high amounts of saturated fat, salt, and sugar. |
| | Maximum score | 11 | |

Answer options (1) never or less than once per month (2) once or twice per month (3) once per week (4) twice per week (5) almost every day (6) once a day or more often

***Body composition,*** including body fat percentage, lean body mass, fat mass and body mass, was measured with a multifrequency bioelectrical impedance analyser (InBody720; Biospace, Seoul, Korea) after overnight fasting at each measurement. Height was measured with a stadiometer at first measurement with a 0.5 cm accuracy.

### 2. Missing data and outliers

Questionnaire-based data had only a few missing values, which were handled with mean imputation, also using values from follow-up measurements if available. Metabolite data were also mainly complete; the rare missing values were left missing.

**Method Supplement Table 2** shows the missing data for each variable and how the missingness was handled. The table also shows outliers excluded from the analysis. It should be noted that a low amount of the largest (XXL and XL) VLDLs were reported for most samples as they were drawn after overnight fasting, and our participants were generally healthy. Few extreme values were noted in some metabolite outcomes when calculating level changes during follow-up. None of the used transformation methods was able to solve their abnormality. Therefore, we excluded these values as outliers (reported in **Method Supplement Table 2.)**

**Method Supplement Table 2.** Missing data, outliers and their handling.

| **Variable** | **Missing** | **Handling method** |
|---|---|---|
| E2 | 0 | |
| FSH | 0 | |
| Body fat percentage | 0 | |
| Self-reported physical activity | 3 | Mean imputation calculated based on all measurements. |
| Smoking | 3 | Smoking status was determined based on follow-up questionnaires assuming that participants did not start smoking between the last follow-up measurement and the final measurement. |
| Alcohol use | 3 | Mean imputation calculated based on all measurements. |
| Education level | 0 | |
| DQS | 5 | The missing value was replaced by the value reported either at the first measurement (if the last measurement value was missing) or the last measurement (if the first measurement value was missing). |
| Total cholesterol | 0 | |
| Total cholesterol minus HDL-C | 0 | |
| Remnant cholesterol | 0 | |
| VLDL cholesterol | 0 | |
| Clinical LDL cholesterol | 0 | |
| LDL cholesterol | 0 | |
| HDL cholesterol | 0 | |
| Total triglycerides | 0 | |
| Triglycerides in VLDL | 0 | |
| Triglycerides in LDL | 0 | |
| Triglycerides in HDL | 0 | |
| Total phospholipids in lipoprotein particles | 0 | |
| Phospholipids in VLDL | 0 | |
| Phospholipids in LDL | 0 | |
| Phospholipids in HDL | 0 | |

| | | |
|---|---|---|
| Total esterified cholesterol | 0 | |
| Cholesteryl esters in VLDL | 0 | |
| Cholesteryl esters in LDL | 0 | |
| Cholesteryl esters in HDL | 0 | |
| Total free cholesterol | 0 | |
| Free cholesterol in VLDL | 0 | |
| Free cholesterol in LDL | 0 | |
| Free cholesterol in HDL | 0 | |
| Total lipids in lipoprotein particles | 0 | |
| Total lipids in VLDL | 0 | |
| Total lipids in LDL | 0 | |
| Total lipids in HDL | 0 | |
| Total concentration of lipoprotein particles | 0 | |
| Concentration of VLDL particles | 0 | |
| Concentration of LDL particles | 0 | |
| Concentration of HDL particles | 0 | |
| Average diameter for VLDL particles | 0 | |
| Average diameter for LDL particles | 0 | |
| Average diameter for HDL particles | 0 | |
| Phosphoglycerides | 0 | |
| Ratio of triglycerides to phosphoglycerides | 0 | |
| Total cholines | 0 | |
| Phosphatidylcholines | 0 | |
| Sphingomyelins | 0 | |
| Apolipoprotein B | 0 | |
| Apolipoprotein A1 | 0 | |
| Ratio of apolipoprotein B to apolipoprotein A1 | 0 | |
| Total fatty acids | 0 | |
| Degree of unsaturation | 0 | 1 outlier removed |
| Omega-3 fatty acids | 0 | |
| Omega-6 fatty acids | 0 | |
| Polyunsaturated fatty acids | 0 | |
| Monounsaturated fatty acids | 0 | |
| Saturated fatty acids | 0 | |
| Linoleic acid | 0 | |
| Docosahexaenoic acid | 0 | |
| Ratio of omega-3 fatty acids to total fatty acids | 0 | |
| Ratio of omega-6 fatty acids to total fatty acids | 0 | |
| Ratio of polyunsaturated fatty acids to total fatty acids | 0 | |
| Ratio of monounsaturated fatty acids to total fatty acids | 0 | |
| Ratio of saturated fatty acids to total fatty acids | 0 | |
| Ratio of linoleic acid to total fatty acids | 0 | |
| Ratio of docosahexaenoic acid to total fatty acids | 0 | 2 outliers removed |
| Ratio of polyunsaturated fatty acids to monounsaturated fatty acids | 0 | |
| Ratio of omega-6 fatty acids to omega-3 fatty acids | 0 | |
| Alanine | 0 | |

| | | |
|---|---|---|
| Glutamine | 0 | |
| Glycine | 0 | |
| Histidine | 0 | |
| Total BCAA | 0 | |
| Isoleucine | 0 | |
| Leucine | 0 | |
| Valine | 0 | |
| Phenylalanine | 0 | |
| Tyrosine | 0 | |
| Glucose | 0 | |
| Lactate | 0 | |
| Pyruvate | 0 | 2 outliers removed |
| Citrate | 0 | |
| Glycerol | 2 | Left missing |
| 3-Hydroxybutyrate | 1 | Left missing |
| Acetate | 0 | |
| Acetoacetate | 0 | |
| Acetone | 0 | |
| Creatinine | 0 | |
| Albumin | 0 | |
| Glycoprotein acetyls | 0 | |
| Concentration of chylomicrons and extremely large VLDL particles | 0 | |
| Total lipids in chylomicrons and extremely large VLDL | 0 | |
| Phospholipids in chylomicrons and extremely large VLDL | 0 | |
| Cholesterol in chylomicrons and extremely large VLDL | 0 | |
| Cholesteryl esters in chylomicrons and extremely large VLDL | 0 | |
| Free cholesterol in chylomicrons and extremely large VLDL | 0 | |
| Triglycerides in chylomicrons and extremely large VLDL | 0 | |
| Concentration of very large VLDL particles | 0 | |
| Total lipids in very large VLDL | 0 | |
| Phospholipids in very large VLDL | 0 | |
| Cholesterol in very large VLDL | 0 | |
| Cholesteryl esters in very large VLDL | 0 | |
| Free cholesterol in very large VLDL | 0 | |
| Triglycerides in very large VLDL | 0 | |
| Concentration of large VLDL particles | 0 | |
| Total lipids in large VLDL | 0 | |
| Phospholipids in large VLDL | 0 | |
| Cholesterol in large VLDL | 0 | |
| Cholesteryl esters in large VLDL | 0 | |
| Free cholesterol in large VLDL | 0 | |
| Triglycerides in large VLDL | 0 | 1 outlier removed |
| Concentration of medium VLDL particles | 0 | |
| Total lipids in medium VLDL | 0 | |
| Phospholipids in medium VLDL | 0 | |
| Cholesterol in medium VLDL | 0 | |

| | | |
|---|---|---|
| Cholesteryl esters in medium VLDL | 0 | |
| Free cholesterol in medium VLDL | 0 | |
| Triglycerides in medium VLDL | 0 | |
| Concentration of small VLDL particles | 0 | |
| Total lipids in small VLDL | 0 | |
| Phospholipids in small VLDL | 0 | |
| Cholesterol in small VLDL | 0 | |
| Cholesteryl esters in small VLDL | 0 | |
| Free cholesterol in small VLDL | 0 | |
| Triglycerides in small VLDL | 0 | |
| Concentration of very small VLDL particles | 0 | |
| Total lipids in very small VLDL | 0 | |
| Phospholipids in very small VLDL | 0 | |
| Cholesterol in very small VLDL | 0 | |
| Cholesteryl esters in very small VLDL | 0 | |
| Free cholesterol in very small VLDL | 0 | |
| Triglycerides in very small VLDL | 0 | |
| Concentration of IDL particles | 0 | |
| Total lipids in IDL | 0 | |
| Phospholipids in IDL | 0 | |
| Cholesterol in IDL | 0 | |
| Cholesteryl esters in IDL | 0 | |
| Free cholesterol in IDL | 0 | |
| Triglycerides in IDL | 0 | |
| Concentration of large LDL particles | 0 | |
| Total lipids in large LDL | 0 | |
| Phospholipids in large LDL | 0 | |
| Cholesterol in large LDL | 0 | |
| Cholesteryl esters in large LDL | 0 | |
| Free cholesterol in large LDL | 0 | |
| Triglycerides in large LDL | 0 | |
| Concentration of medium LDL particles | 0 | |
| Total lipids in medium LDL | 0 | |
| Phospholipids in medium LDL | 0 | |
| Cholesterol in medium LDL | 0 | |
| Cholesteryl esters in medium LDL | 0 | |
| Free cholesterol in medium LDL | 0 | |
| Triglycerides in medium LDL | 0 | |
| Concentration of small LDL particles | 0 | |
| Total lipids in small LDL | 0 | |
| Phospholipids in small LDL | 0 | |
| Cholesterol in small LDL | 0 | |
| Cholesteryl esters in small LDL | 0 | |
| Free cholesterol in small LDL | 0 | 1 outlier removed |
| Triglycerides in small LDL | 0 | |
| Concentration of very large HDL particles | 0 | |

| | | |
|---|---|---|
| Total lipids in very large HDL | 0 | |
| Phospholipids in very large HDL | 0 | 1 outlier removed |
| Cholesterol in very large HDL | 0 | |
| Cholesteryl esters in very large HDL | 0 | |
| Free cholesterol in very large HDL | 0 | |
| Triglycerides in very large HDL | 0 | |
| Concentration of large HDL particles | 0 | |
| Total lipids in large HDL | 0 | |
| Phospholipids in large HDL | 0 | |
| Cholesterol in large HDL | 0 | |
| Cholesteryl esters in large HDL | 0 | |
| Free cholesterol in large HDL | 0 | |
| Triglycerides in large HDL | 0 | |
| Concentration of medium HDL particles | 0 | |
| Total lipids in medium HDL | 0 | |
| Phospholipids in medium HDL | 0 | |
| Cholesterol in medium HDL | 0 | |
| Cholesteryl esters in medium HDL | 0 | |
| Free cholesterol in medium HDL | 0 | |
| Triglycerides in medium HDL | 0 | |
| Concentration of small HDL particles | 0 | |
| Total lipids in small HDL | 0 | |
| Phospholipids in small HDL | 0 | |
| Cholesterol in small HDL | 0 | |
| Cholesteryl esters in small HDL | 0 | |
| Free cholesterol in small HDL | 0 | |
| Triglycerides in small HDL | 0 | |

### 3. Statistical analyses

All statistical analyses were performed using R version 4.0.0. or higher unless stated otherwise.[5] As recommended by the STROBE statement, we do not report statistical significance testing of within-group changes or between-group comparison in descriptive statistics.[6]

#### 3.1. Primary analysis

Our primary results were associations between menopause and metabolite measures in women experiencing natural menopause, i.e. who did not start menopausal hormone therapy (MHT) during follow-up ($n$ = 183). Metabolite measure skewness and kurtosis were estimated using the *psych* package.[7] Metabolite distributions were mostly skewed as expected. To minimise the biasing impact from non-normality and to stabilise variance (e.g., with respect to outliers and extreme values), we transformed the metabolite measures using the Box-Cox transformation with lambda parameter estimated from data for each variable separately with the *MASS* package, version 7.3-54.[8] For the two longitudinal measurements of each variable, we performed a grid search to identify a lambda parameter that was optimal for the transformation of both measurement occasions. In six measures,

Box-Cox transformation was not fully able to correct variable distributions, and outliers remained. These outliers were removed from the analyses and are shown in **Method Supplement Table 2**.

Metabolite measures were standardised with respect to baseline mean and standard deviation to aid comparability between measures with different units and concentrations. We also reported absolute mean values of metabolite measures with standard deviations at the first and last measurement time points to aid clinical interpretation (**Result Supplement Table 1**).

We performed the analyses using linear mixed-effect models with random intercept using the *nlme* package.[9] We created three models to study the primary research question. Metabolite measures were the outcomes, and menopausal status (0 = pre, 1 = post) was a fixed-effect in all models. The second model included age at the first measurement, follow-up duration, education level, smoking status, physical activity, and diet quality as fixed effects. Even though some of the covariates cannot be strictly determined as confounders as they do not influence the menopausal status progression, we considered this model to answer our research question the best as it adjusts the estimates to variables potentially influencing metabolite levels. In the third model, body fat percentage was included as a fixed effect. Body composition is a known mediator between menopause and metabolites, and therefore, it should not be included in the model as it may mask the potential effects of menopause. However, as we wanted to examine the possible direct role menopausal hormonal shift plays, we performed the analyses also body fat percentage adjusted.

Metabolomics requires multiple testing, which should be taken into account. Due to the highly correlated nature of the metabolite measures provided by the used NMR platform,[10] Bonferroni correction is too conservative. Several other multiple testing methods have been suggested, but using simulation studies, we found that the optimal approach in our case was based on accounting the number of independent sources of variability (eigenvalues of the correlation matrix) in the data. Therefore, in this study, the $K_{eff}$-Šidák correction was used to account for multiple testing.[11] Because the adjustment involves evaluating a double integral, we performed it using a custom C-script with a wrapper function in R.[5] The Šidák-part of the correction was used instead of the Bonferroni approach because of better performance in simulations.

#### 3.2. Explorative analysis I – menopausal hormonal shift and metabolite changes

Our first explorative analysis investigated if the menopausal hormonal shift explained metabolite measure changes in women experiencing natural menopause ($n$ = 183). The direct and indirect associations from hormones to metabolites via fat percentage were examined using latent change score modelling with the model shown and explained in **Method Supplement Fig 1**.

The structure of the model imposes a different relationship for each repeated measurement of the variables so that the latent variable corresponds to the follow-up minus baseline difference adjusted for the baseline measurement. This model can be seen as an extension of the paired $t$-test,[12] where change can be controlled for baseline measurement. We used Mplus,[13] version 7.4, to estimate model parameters. The effect size for direct effect is the squared semi-partial correlation and for the indirect effect extension of upsilon, see Lachowicz et al.[14] Briefly, the new indirect effect size estimate removes spurious correlation from the estimate and therefore, leads to a consistent estimate that avoids problematic estimates to occur. We extended the effect size measure to account for more than a single variable. The associations can be interpreted similarly to squared semi-partial correlations: they describe how much of the metabolite change the menopausal hormonal shift explains. Multiple testing correction was performed using false discovery rate adjustment.

#### 3.3. Explorative analysis II – menopausal hormone therapy and metabolite trajectory

As 35 women started oestrogen-containing menopausal hormone therapy (MHT) during follow-up, we assessed using the whole study sample data ($N$ = 218) if MHT modulated metabolite trajectories. Model structures we as in primary analyses, except that menopausal status and MHT use (0 = no MHT, 1 = MHT user) interaction was included as a fixed effect.

#### 3.4. Figures

We produced **Fig. 2** using the *ggforestplot* package,[15] and Fig. 3 using the *ggplot2 package*.[16] Graphical abstract and **Fig. 1** were produced with BioRender.com.

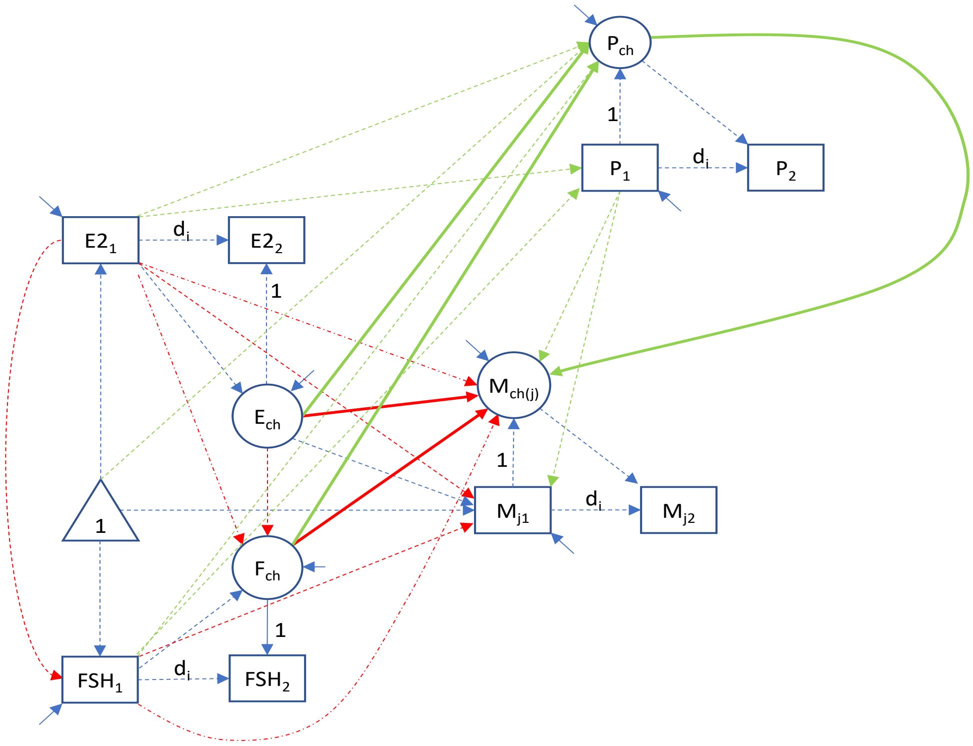


**Method Supplement Figure 1.** Latent change score model used to assess baseline-adjusted relationships among changes in oestradiol (E2), follicle-stimulating hormone (FSH), metabolite and fat percentage. Names enclosed in rectangles indicate observed variables, circles latent variables and triangles means/intercepts. Pathways of interest included associations between changes in oestradiol and follicle-stimulating hormone with change in metabolite j ($E_{ch} \rightarrow M_{ch(j)}$ and $F_{ch} \rightarrow M_{ch(j)}$, $j = 1, \ldots, 180$), as well as indirect associations from changes in estradiol and follicle-stimulating hormone on metabolite changes through changes in fat percentage ($E_{ch} \rightarrow P_{ch} \rightarrow M_{ch(j)}$ and $F_{ch} \rightarrow P_{ch} \rightarrow M_{ch(j)}$). The model was applied for each metabolite separately. Paths of interest are shown with a solid line, nuisance paths (i.e. paths necessary to estimate but not of substantive importance) are shown with dashed lines, fixed path coefficients are indicated with fixed parameter value, and mean and variance parameters constrained to zero are omitted from the figure. $E2_i$ = oestradiol, $FSH_i$ = follicle-stimulating hormone, $M_i$ = metabolite, $P_i$ = body fat percentage ($_i$ = 1 - baseline, 2 - follow-up), $d_i$ = time difference between baseline and follow-up for individual $_i$, $E_{ch}$ = change score for estradiol, $F_{ch}$ = change score for FSH, $M_{ch}$ = change score for metabolite, $P_{ch}$ = change score for body fat percentage.

4. Nordic Council of Ministers E. Nordic Nutrition Recommendations 2012. Volume 5. Nordisk Ministerråd; Kopenhagen, Denmark: 2012.

5. R Core Team. R: A language and environment for statistical computing. R Foundation for Statistical Computing, Vienna, Austria. URL https://www.R-project.org/: 2018-2021.

6. Vandenbroucke JP, Elm E von, Altman DG, Gøtzsche PC, Mulrow CD, Pocock SJ, Poole C, Schlesselman JJ, Egger M, STROBE initiative. Strengthening the Reporting of Observational Studies in Epidemiology (STROBE): explanation and elaboration. *Ann Intern Med* 2007;**147**:W163-194.

7. Revelle W. psych: Procedures for Personality and Psychological Research, Northwestern University, Evanston, Illinois, USA, 2020, https://CRAN.R-project.org/package=psych Version = 2.1.3.

8. Venables WN, Ripley BD. Modern Applied Statistics with S. Fourth Edition. Springer, New Yor k, 2020.

15. Scheinin I, Kalimeri M, Jagerroos V, Parkkinen J, Tikkanen E, Würtz P, Kangas A. (2020). ggforestplot: Forestplots of Measures of Effects and Their Confidence Intervals.

16. Wickham H. ggplot2: Elegant Graphics for Data Analysis. Springer-Verlag New York, 2016.
