## Supplementary material for "Menopause modulates the circulating metabolome: evidence from a prospective cohort study": Result Supplement

**Pages 1–6, Result Supplement Table 1.** Descriptive statistics for metabolite measures among women experiencing natural menopause (n = 183) during the study.

**Pages 7–12, Result Supplement Table 2.** Descriptive statistics for metabolite measures among women starting menopausal hormone therapy (*n* = 35) during the study.

**Pages 13–18, Result Supplement Table 3.** Menopause-associated metabolite changes.

**Pages 19–25, Result Supplement Table 4.** Direct and indirect associations (via body composition change) of menopausal hormonal shift and metabolite changes.

**Pages 26–31, Result Supplement Table 5.** Associations between menopausal hormone therapy and metabolite measures.

**Result Supplement Table 1**. Descriptive statistics for metabolite measures among women experiencing natural menopause (*n* = 183) during the study.

| | | First measurement | | | | Last measurement | | | | Change (last -first measurement) | | | |
|---|---|---|---|---|---|---|---|---|---|---|---|---|---|
| **Metabolite** | ***n*** | **Mean** | **SD** | **Median** | **IQR** | **Mean** | **SD** | **Median** | **IQR** | **Mean** | **SD** | **Median** | **IQR** |
| ***Cholesterols*** | | | | | | | | | | | | | |
| Total cholesterol | 183 | 5.580 | 0.821 | 5.527 | 0.959 | 5.732 | 0.855 | 5.692 | 1.068 | 0.152 | 0.593 | 0.088 | 0.682 |
| Non-HDL cholesterol | 183 | 3.830 | 0.801 | 3.740 | 0.966 | 3.991 | 0.830 | 3.902 | 1.087 | 0.160 | 0.507 | 0.103 | 0.615 |
| Remnant cholesterol | 183 | 1.678 | 0.364 | 1.647 | 0.456 | 1.757 | 0.389 | 1.727 | 0.486 | 0.079 | 0.225 | 0.057 | 0.283 |
| VLDL cholesterol | 183 | 0.675 | 0.228 | 0.639 | 0.286 | 0.724 | 0.246 | 0.707 | 0.280 | 0.049 | 0.140 | 0.037 | 0.161 |
| Clinical LDL cholesterol | 183 | 3.164 | 0.707 | 3.070 | 0.811 | 3.286 | 0.711 | 3.179 | 0.924 | 0.122 | 0.489 | 0.119 | 0.558 |
| LDL cholesterol | 183 | 2.153 | 0.453 | 2.069 | 0.518 | 2.234 | 0.457 | 2.173 | 0.584 | 0.081 | 0.306 | 0.074 | 0.337 |
| HDL cholesterol | 183 | 1.749 | 0.332 | 1.713 | 0.403 | 1.741 | 0.338 | 1.727 | 0.463 | -0.008 | 0.175 | -0.023 | 0.216 |
| ***Triglycerides*** | | | | | | | | | | | | | |
| Total triglycerides | 183 | 1.128 | 0.525 | 0.987 | 0.586 | 1.233 | 0.564 | 1.114 | 0.596 | 0.106 | 0.484 | 0.090 | 0.415 |
| VLDL triglycerides | 183 | 0.756 | 0.450 | 0.639 | 0.492 | 0.845 | 0.495 | 0.740 | 0.516 | 0.089 | 0.421 | 0.081 | 0.310 |
| LDL triglycerides | 183 | 0.152 | 0.033 | 0.146 | 0.036 | 0.157 | 0.033 | 0.150 | 0.038 | 0.005 | 0.025 | 0.005 | 0.026 |
| HDL triglycerides | 183 | 0.119 | 0.044 | 0.115 | 0.052 | 0.128 | 0.043 | 0.119 | 0.048 | 0.009 | 0.041 | 0.009 | 0.046 |
| ***Phospholipids*** | | | | | | | | | | | | | |
| Total phospholipids | 183 | 3.330 | 0.388 | 3.317 | 0.458 | 3.409 | 0.413 | 3.374 | 0.525 | 0.079 | 0.302 | 0.050 | 0.389 |
| VLDL phospholipids | 183 | 0.420 | 0.170 | 0.387 | 0.194 | 0.456 | 0.182 | 0.430 | 0.204 | 0.036 | 0.121 | 0.030 | 0.122 |
| LDL phospholipids | 183 | 0.729 | 0.138 | 0.707 | 0.160 | 0.752 | 0.140 | 0.737 | 0.177 | 0.023 | 0.091 | 0.018 | 0.106 |
| HDL phospholipids | 183 | 1.853 | 0.302 | 1.832 | 0.388 | 1.862 | 0.313 | 1.848 | 0.410 | 0.009 | 0.179 | -0.009 | 0.231 |
| ***Cholesteryl esters*** | | | | | | | | | | | | | |
| Total cholesteryl esters | 183 | 4.103 | 0.590 | 4.055 | 0.676 | 4.207 | 0.611 | 4.170 | 0.744 | 0.104 | 0.438 | 0.046 | 0.504 |
| VLDL cholesteryl esters | 183 | 0.413 | 0.129 | 0.396 | 0.164 | 0.439 | 0.139 | 0.430 | 0.177 | 0.027 | 0.076 | 0.019 | 0.094 |
| LDL cholesteryl esters | 183 | 1.562 | 0.340 | 1.496 | 0.390 | 1.626 | 0.345 | 1.591 | 0.439 | 0.064 | 0.223 | 0.053 | 0.257 |
| HDL cholesteryl esters | 183 | 1.369 | 0.260 | 1.353 | 0.332 | 1.361 | 0.264 | 1.344 | 0.378 | -0.008 | 0.138 | -0.017 | 0.169 |
| ***Free cholesterol*** | | | | | | | | | | | | | |
| Total free cholesterol | 183 | 1.477 | 0.237 | 1.460 | 0.294 | 1.525 | 0.250 | 1.506 | 0.319 | 0.048 | 0.159 | 0.034 | 0.191 |
| VLDL free cholesterol | 183 | 0.262 | 0.101 | 0.241 | 0.122 | 0.285 | 0.110 | 0.271 | 0.125 | 0.023 | 0.069 | 0.019 | 0.069 |
| LDL free cholesterol | 183 | 0.591 | 0.117 | 0.577 | 0.142 | 0.608 | 0.116 | 0.586 | 0.149 | 0.017 | 0.087 | 0.017 | 0.093 |
| HDL free cholesterol | 183 | 0.381 | 0.074 | 0.375 | 0.086 | 0.381 | 0.076 | 0.377 | 0.096 | 0.000 | 0.039 | -0.005 | 0.052 |
| ***Total lipids*** | | | | | | | | | | | | | |
| Total lipids | 183 | 10.037 | 1.445 | 9.922 | 1.865 | 10.374 | 1.541 | 10.258 | 1.872 | 0.337 | 1.083 | 0.194 | 1.211 |
| VLDL total lipids | 183 | 1.851 | 0.815 | 1.673 | 0.975 | 2.026 | 0.887 | 1.872 | 0.968 | 0.175 | 0.654 | 0.148 | 0.600 |
| LDL total lipids | 183 | 3.034 | 0.609 | 2.937 | 0.694 | 3.143 | 0.617 | 3.084 | 0.782 | 0.109 | 0.400 | 0.086 | 0.450 |
| HDL total lipids | 183 | 3.722 | 0.621 | 3.660 | 0.707 | 3.731 | 0.643 | 3.685 | 0.879 | 0.010 | 0.347 | -0.038 | 0.464 |
| ***Particles*** | | | | | | | | | | | | | |
| Total particles | 183 | 0.020 | 0.002 | 0.020 | 0.003 | 0.021 | 0.002 | 0.020 | 0.003 | 0.000 | 0.002 | 0.000 | 0.002 |
| VLDL particles | 183 | 1.40e-04 | 4.30e-05 | 1.33e-04 | 5.60e-05 | 1.49e-04 | 4.59e-05 | 1.44e-04 | 4.90e-05 | 9.12e-06 | 2.86e-05 | 7.51e-06 | 2.94e-05 |
| LDL particles | 183 | 1.35e-03 | 2.83e-04 | 1.29e-03 | 3.40e-04 | 1.40e-03 | 2.92e-04 | 1.38e-03 | 3.50e-04 | 5.06e-05 | 1.57e-04 | 2.69e-05 | 1.89e-04 |
| HDL particles | 183 | 0.018 | 0.002 | 0.018 | 0.003 | 0.019 | 0.002 | 0.018 | 0.003 | 0.000 | 0.002 | 0.000 | 0.002 |

| | | First measurement | | | | Last measurement | | | | Change (last -first measurement) | | | |
|---|---|---|---|---|---|---|---|---|---|---|---|---|---|
| **Metabolite** | ***n*** | **Mean** | **SD** | **Median** | **IQR** | **Mean** | **SD** | **Median** | **IQR** | **Mean** | **SD** | **Median** | **IQR** |
| ***Particle diameter*** | | | | | | | | | | | | | |
| VLDL size | 183 | 38.106 | 1.107 | 38.036 | 1.418 | 38.331 | 1.109 | 38.258 | 1.526 | 0.225 | 0.855 | 0.257 | 0.864 |
| LDL size | 183 | 23.911 | 0.077 | 23.915 | 0.118 | 23.922 | 0.076 | 23.926 | 0.113 | 0.010 | 0.078 | 0.013 | 0.103 |
| HDL size | 183 | 9.774 | 0.196 | 9.768 | 0.262 | 9.753 | 0.195 | 9.724 | 0.296 | -0.020 | 0.093 | -0.020 | 0.121 |
| ***Phosphoglycerides*** | | | | | | | | | | | | | |
| Phosphoglycerides | 183 | 2.812 | 0.330 | 2.793 | 0.390 | 2.816 | 0.360 | 2.777 | 0.431 | 0.003 | 0.279 | -0.008 | 0.315 |
| Tri-/phosphoglyceride ratio | 183 | 0.398 | 0.160 | 0.359 | 0.193 | 0.436 | 0.177 | 0.395 | 0.218 | 0.038 | 0.133 | 0.038 | 0.123 |
| Total cholines | 183 | 3.169 | 0.337 | 3.154 | 0.432 | 3.165 | 0.373 | 3.118 | 0.441 | -0.004 | 0.282 | -0.005 | 0.319 |
| Phosphatidylcholines | 183 | 2.666 | 0.319 | 2.648 | 0.392 | 2.672 | 0.351 | 2.619 | 0.417 | 0.007 | 0.273 | 0.005 | 0.330 |
| Sphingomyelins | 183 | 0.545 | 0.061 | 0.544 | 0.080 | 0.549 | 0.063 | 0.542 | 0.079 | 0.004 | 0.045 | -0.004 | 0.051 |
| ***Apolipoproteins*** | | | | | | | | | | | | | |
| Apolipoprotein B | 183 | 0.937 | 0.191 | 0.912 | 0.235 | 0.973 | 0.199 | 0.964 | 0.241 | 0.036 | 0.107 | 0.019 | 0.131 |
| Apolipoprotein A-I | 183 | 1.736 | 0.225 | 1.736 | 0.290 | 1.748 | 0.233 | 1.752 | 0.312 | 0.011 | 0.142 | -0.006 | 0.192 |
| Apo B/A-I ratio | 183 | 0.551 | 0.143 | 0.527 | 0.184 | 0.569 | 0.150 | 0.546 | 0.184 | 0.018 | 0.064 | 0.019 | 0.072 |
| ***Fatty acids*** | | | | | | | | | | | | | |
| Total fatty acids | 183 | 13.491 | 2.058 | 13.275 | 2.433 | 13.653 | 2.190 | 13.339 | 2.485 | 0.162 | 1.733 | -0.017 | 1.830 |
| Degree of unsaturation | 182 | 1.353 | 0.057 | 1.351 | 0.070 | 1.344 | 0.063 | 1.343 | 0.089 | -0.008 | 0.048 | -0.005 | 0.057 |
| Omega-3 fatty acids | 183 | 0.624 | 0.181 | 0.603 | 0.218 | 0.611 | 0.206 | 0.576 | 0.234 | -0.013 | 0.150 | -0.011 | 0.181 |
| Omega-6 fatty acids | 183 | 5.241 | 0.577 | 5.231 | 0.716 | 5.223 | 0.669 | 5.150 | 0.799 | -0.019 | 0.514 | -0.040 | 0.607 |
| Polyunsaturated fatty acids | 183 | 5.865 | 0.682 | 5.834 | 0.822 | 5.834 | 0.799 | 5.693 | 1.011 | -0.031 | 0.573 | -0.105 | 0.677 |
| Monounsaturated fatty acids | 183 | 3.237 | 0.752 | 3.066 | 0.820 | 3.287 | 0.767 | 3.148 | 0.822 | 0.050 | 0.661 | 0.005 | 0.645 |
| Saturated fatty acids | 183 | 4.388 | 0.766 | 4.254 | 0.829 | 4.532 | 0.802 | 4.393 | 0.842 | 0.143 | 0.681 | 0.070 | 0.612 |
| Linoleic acid | 183 | 4.701 | 0.581 | 4.666 | 0.733 | 4.533 | 0.781 | 4.539 | 1.003 | -0.168 | 0.658 | -0.216 | 0.847 |
| Docosahexaenoic acid | 183 | 0.322 | 0.067 | 0.316 | 0.074 | 0.311 | 0.076 | 0.296 | 0.087 | -0.012 | 0.057 | -0.011 | 0.069 |
| ***Fatty acid ratios (%)*** | | | | | | | | | | | | | |
| Omega-3 ratio | 183 | 4.616 | 1.117 | 4.523 | 1.470 | 4.451 | 1.178 | 4.303 | 1.396 | -0.165 | 1.006 | -0.117 | 1.187 |
| Omega-6 ratio | 183 | 39.111 | 2.412 | 39.301 | 2.739 | 38.496 | 2.536 | 38.937 | 2.642 | -0.615 | 2.213 | -0.399 | 2.471 |
| Polyunsaturated ratio | 183 | 43.727 | 2.557 | 44.038 | 3.027 | 42.947 | 2.696 | 43.127 | 3.243 | -0.781 | 2.401 | -0.521 | 2.768 |
| Monounsaturated ratio | 183 | 23.806 | 2.201 | 23.536 | 2.719 | 23.884 | 2.200 | 23.731 | 3.018 | 0.078 | 1.752 | -0.026 | 2.039 |
| Saturated ratio | 183 | 32.467 | 1.274 | 32.379 | 1.523 | 33.170 | 1.743 | 33.122 | 2.222 | 0.703 | 1.685 | 0.503 | 2.275 |
| Linoleic acid ratio | 183 | 35.053 | 2.538 | 35.123 | 3.313 | 33.286 | 3.255 | 33.416 | 4.413 | -1.766 | 3.060 | -1.401 | 4.429 |
| Docosahexaenoic acid ratio | 181 | 2.414 | 0.471 | 2.377 | 0.646 | 2.292 | 0.477 | 2.224 | 0.588 | -0.122 | 0.387 | -0.107 | 0.448 |
| Polyu/monounsaturated ratio | 183 | 1.860 | 0.262 | 1.860 | 0.314 | 1.821 | 0.258 | 1.810 | 0.359 | -0.039 | 0.208 | -0.044 | 0.249 |
| Omega-6 to omega-3 ratio | 183 | 9.010 | 2.535 | 8.540 | 3.061 | 9.279 | 2.681 | 8.952 | 3.194 | 0.269 | 2.013 | 0.006 | 2.773 |
| ***Amino acids*** | | | | | | | | | | | | | |
| Alanine | 183 | 0.383 | 0.067 | 0.379 | 0.091 | 0.392 | 0.070 | 0.375 | 0.090 | 0.009 | 0.069 | 0.009 | 0.074 |
| Glutamine | 183 | 0.762 | 0.059 | 0.759 | 0.073 | 0.730 | 0.080 | 0.737 | 0.122 | -0.032 | 0.075 | -0.025 | 0.110 |
| Glycine | 183 | 0.316 | 0.079 | 0.295 | 0.099 | 0.325 | 0.080 | 0.304 | 0.094 | 0.009 | 0.044 | 0.008 | 0.047 |
| Histidine | 183 | 0.079 | 0.009 | 0.079 | 0.012 | 0.080 | 0.010 | 0.080 | 0.011 | 0.001 | 0.009 | 0.001 | 0.011 |
| Total BCAA | 183 | 0.383 | 0.051 | 0.378 | 0.054 | 0.395 | 0.056 | 0.395 | 0.081 | 0.012 | 0.051 | 0.012 | 0.070 |
| Isoleucine | 183 | 0.048 | 0.009 | 0.047 | 0.011 | 0.050 | 0.009 | 0.049 | 0.012 | 0.002 | 0.009 | 0.001 | 0.012 |
| Leucine | 183 | 0.108 | 0.015 | 0.107 | 0.019 | 0.113 | 0.018 | 0.111 | 0.025 | 0.004 | 0.015 | 0.005 | 0.022 |
| Valine | 183 | 0.226 | 0.030 | 0.222 | 0.039 | 0.233 | 0.032 | 0.233 | 0.044 | 0.006 | 0.030 | 0.007 | 0.043 |

| | | First measurement | | | | Last measurement | | | | Change (last -first measurement) | | | |
|---|---|---|---|---|---|---|---|---|---|---|---|---|---|
| **Metabolite** | ***n*** | **Mean** | **SD** | **Median** | **IQR** | **Mean** | **SD** | **Median** | **IQR** | **Mean** | **SD** | **Median** | **IQR** |
| ***Amino acids continue*** | | | | | | | | | | | | | |
| Phenylalanine | 183 | 0.066 | 0.008 | 0.065 | 0.011 | 0.067 | 0.009 | 0.067 | 0.011 | 0.001 | 0.009 | 0.001 | 0.012 |
| Tyrosine | 183 | 0.064 | 0.010 | 0.063 | 0.014 | 0.066 | 0.010 | 0.064 | 0.014 | 0.002 | 0.009 | 0.003 | 0.011 |
| ***Glycolysis-related*** | | | | | | | | | | | | | |
| Glucose | 183 | 5.279 | 0.426 | 5.248 | 0.517 | 5.248 | 0.502 | 5.222 | 0.647 | -0.030 | 0.370 | -0.014 | 0.536 |
| Lactate | 183 | 1.394 | 0.454 | 1.318 | 0.411 | 1.401 | 0.468 | 1.323 | 0.487 | 0.007 | 0.573 | -0.019 | 0.456 |
| Pyruvate | 181 | 0.057 | 0.017 | 0.054 | 0.020 | 0.059 | 0.022 | 0.057 | 0.025 | 0.002 | 0.022 | 0.002 | 0.022 |
| Citrate | 183 | 0.063 | 0.010 | 0.062 | 0.012 | 0.060 | 0.010 | 0.059 | 0.013 | -0.003 | 0.008 | -0.003 | 0.010 |
| Glycerol | 183 | 0.124 | 0.036 | 0.117 | 0.040 | 0.138 | 0.042 | 0.131 | 0.050 | 0.014 | 0.041 | 0.011 | 0.044 |
| ***Ketone bodies*** | | | | | | | | | | | | | |
| 3-Hydroxybutyrate | 183 | 0.069 | 0.097 | 0.032 | 0.064 | 0.034 | 0.043 | 0.019 | 0.032 | -0.035 | 0.085 | -0.013 | 0.054 |
| Acetate | 183 | 0.029 | 0.015 | 0.028 | 0.015 | 0.036 | 0.052 | 0.028 | 0.019 | 0.007 | 0.051 | 0.001 | 0.018 |
| Acetoacetate | 183 | 0.035 | 0.030 | 0.025 | 0.024 | 0.024 | 0.017 | 0.019 | 0.016 | -0.011 | 0.029 | -0.004 | 0.020 |
| Acetone | 183 | 0.018 | 0.008 | 0.015 | 0.006 | 0.016 | 0.005 | 0.014 | 0.004 | -0.002 | 0.008 | -0.001 | 0.006 |
| ***Miscellaneous*** | | | | | | | | | | | | | |
| Creatinine | 183 | 70.319 | 9.997 | 70.416 | 13.046 | 70.860 | 9.612 | 70.013 | 12.231 | 0.541 | 6.401 | 0.621 | 7.818 |
| Albumin | 183 | 43.022 | 2.583 | 43.312 | 3.395 | 43.197 | 2.668 | 43.237 | 3.528 | 0.175 | 2.349 | 0.202 | 3.277 |
| Glycoprotein acetyls | 183 | 0.855 | 0.100 | 0.848 | 0.143 | 0.849 | 0.110 | 0.838 | 0.151 | -0.006 | 0.081 | -0.007 | 0.095 |
| ***Chylomicrons and extremely large VLDL*** | | | | | | | | | | | | | |
| Particles | 183 | 4.75e-07 | 1.03e-06 | 3.89e-08 | 5.32e-07 | 6.43e-07 | 1.10e-06 | 2.09e-07 | 8.65e-07 | 1.68e-07 | 1.17e-06 | 7.20e-09 | 3.41e-07 |
| Total lipids | 183 | 0.066 | 0.140 | 0.017 | 0.060 | 0.087 | 0.153 | 0.025 | 0.107 | 0.021 | 0.166 | 0.002 | 0.050 |
| Phospholipids | 183 | 0.007 | 0.020 | 0.000 | 0.007 | 0.010 | 0.020 | 0.000 | 0.013 | 0.003 | 0.023 | 0.000 | 0.005 |
| Cholesterol | 183 | 0.023 | 0.029 | 0.015 | 0.024 | 0.027 | 0.030 | 0.018 | 0.032 | 0.004 | 0.030 | 0.001 | 0.015 |
| Cholesteryl esters | 183 | 0.017 | 0.017 | 0.014 | 0.019 | 0.019 | 0.018 | 0.015 | 0.023 | 0.002 | 0.017 | 0.000 | 0.011 |
| Free cholesterol | 183 | 0.006 | 0.013 | 0.001 | 0.006 | 0.008 | 0.013 | 0.003 | 0.010 | 0.002 | 0.014 | 0.000 | 0.005 |
| Triglycerides | 183 | 0.035 | 0.092 | 0.001 | 0.034 | 0.050 | 0.104 | 0.007 | 0.061 | 0.014 | 0.114 | 0.000 | 0.028 |
| ***Very large VLDL*** | | | | | | | | | | | | | |
| Particles | 183 | 2.49e-06 | 2.12e-06 | 1.98e-06 | 2.38e-06 | 2.93e-06 | 2.35e-06 | 2.45e-06 | 2.54e-06 | 4.43e-07 | 1.86e-06 | 4.16e-07 | 1.35e-06 |
| Total lipids | 183 | 0.145 | 0.126 | 0.111 | 0.143 | 0.171 | 0.142 | 0.138 | 0.156 | 0.026 | 0.113 | 0.024 | 0.082 |
| Phospholipids | 183 | 0.024 | 0.023 | 0.019 | 0.026 | 0.029 | 0.026 | 0.023 | 0.028 | 0.005 | 0.021 | 0.004 | 0.014 |
| Cholesterol | 183 | 0.043 | 0.027 | 0.038 | 0.034 | 0.048 | 0.030 | 0.043 | 0.034 | 0.005 | 0.020 | 0.004 | 0.019 |
| Cholesteryl esters | 183 | 0.027 | 0.014 | 0.025 | 0.019 | 0.030 | 0.016 | 0.028 | 0.019 | 0.002 | 0.010 | 0.002 | 0.010 |
| Free cholesterol | 183 | 0.015 | 0.013 | 0.013 | 0.016 | 0.018 | 0.014 | 0.016 | 0.015 | 0.003 | 0.011 | 0.002 | 0.009 |
| Triglycerides | 183 | 0.078 | 0.077 | 0.057 | 0.084 | 0.094 | 0.087 | 0.073 | 0.090 | 0.016 | 0.073 | 0.015 | 0.049 |
| ***Large VLDL*** | | | | | | | | | | | | | |
| Particles | 183 | 8.82e-06 | 5.61e-06 | 7.34e-06 | 6.57e-06 | 9.94e-06 | 6.19e-06 | 8.70e-06 | 6.82e-06 | 1.12e-06 | 4.68e-06 | 1.22e-06 | 3.97e-06 |
| Total lipids | 183 | 0.290 | 0.186 | 0.250 | 0.224 | 0.329 | 0.206 | 0.293 | 0.235 | 0.039 | 0.155 | 0.041 | 0.132 |
| Phospholipids | 183 | 0.054 | 0.040 | 0.045 | 0.049 | 0.061 | 0.044 | 0.053 | 0.050 | 0.008 | 0.032 | 0.008 | 0.028 |
| Cholesterol | 183 | 0.081 | 0.049 | 0.073 | 0.063 | 0.092 | 0.053 | 0.083 | 0.062 | 0.011 | 0.036 | 0.010 | 0.037 |
| Cholesteryl esters | 183 | 0.043 | 0.024 | 0.040 | 0.030 | 0.048 | 0.026 | 0.045 | 0.030 | 0.005 | 0.017 | 0.004 | 0.017 |
| Free cholesterol | 183 | 0.038 | 0.025 | 0.033 | 0.031 | 0.043 | 0.028 | 0.039 | 0.032 | 0.005 | 0.019 | 0.005 | 0.019 |
| Triglycerides | 182 | 0.156 | 0.099 | 0.128 | 0.122 | 0.177 | 0.111 | 0.158 | 0.120 | 0.021 | 0.088 | 0.021 | 0.074 |

| | | First measurement | | | | Last measurement | | | | Change (last -first measurement) | | | |
|---|---|---|---|---|---|---|---|---|---|---|---|---|---|
| **Metabolite** | ***n*** | **Mean** | **SD** | **Median** | **IQR** | **Mean** | **SD** | **Median** | **IQR** | **Mean** | **SD** | **Median** | **IQR** |
| ***Medium VLDL*** | | | | | | | | | | | | | |
| Particles | 183 | 3.65e-05 | 1.27e-05 | 3.40e-05 | 1.65e-05 | 3.91e-05 | 1.36e-05 | 3.78e-05 | 1.54e-05 | 2.69e-06 | 8.04e-06 | 2.26e-06 | 9.60e-06 |
| Total lipids | 183 | 0.607 | 0.221 | 0.578 | 0.287 | 0.654 | 0.236 | 0.617 | 0.275 | 0.047 | 0.152 | 0.041 | 0.178 |
| Phospholipids | 183 | 0.137 | 0.048 | 0.128 | 0.062 | 0.147 | 0.051 | 0.144 | 0.060 | 0.010 | 0.030 | 0.008 | 0.037 |
| Cholesterol | 183 | 0.189 | 0.060 | 0.181 | 0.072 | 0.201 | 0.064 | 0.196 | 0.075 | 0.012 | 0.035 | 0.009 | 0.044 |
| Cholesteryl esters | 183 | 0.105 | 0.033 | 0.100 | 0.041 | 0.111 | 0.035 | 0.108 | 0.045 | 0.006 | 0.022 | 0.002 | 0.025 |
| Free cholesterol | 183 | 0.084 | 0.029 | 0.079 | 0.035 | 0.090 | 0.031 | 0.087 | 0.036 | 0.006 | 0.017 | 0.004 | 0.022 |
| Triglycerides | 183 | 0.280 | 0.128 | 0.249 | 0.163 | 0.305 | 0.137 | 0.280 | 0.162 | 0.025 | 0.106 | 0.026 | 0.100 |
| ***Small VLDL*** | | | | | | | | | | | | | |
| Particles | 183 | 3.80e-05 | 1.32e-05 | 3.59e-05 | 1.73e-05 | 4.05e-05 | 1.38e-05 | 3.93e-05 | 1.54e-05 | 2.47e-06 | 9.02e-06 | 2.54e-06 | 9.38e-06 |
| Total lipids | 183 | 0.400 | 0.133 | 0.382 | 0.178 | 0.426 | 0.138 | 0.416 | 0.156 | 0.026 | 0.089 | 0.025 | 0.094 |
| Phospholipids | 183 | 0.101 | 0.030 | 0.097 | 0.038 | 0.107 | 0.032 | 0.104 | 0.036 | 0.006 | 0.018 | 0.005 | 0.022 |
| Cholesterol | 183 | 0.156 | 0.050 | 0.149 | 0.064 | 0.165 | 0.053 | 0.160 | 0.066 | 0.010 | 0.028 | 0.008 | 0.038 |
| Cholesteryl esters | 183 | 0.092 | 0.032 | 0.089 | 0.039 | 0.098 | 0.033 | 0.094 | 0.042 | 0.006 | 0.018 | 0.006 | 0.024 |
| Free cholesterol | 183 | 0.064 | 0.019 | 0.061 | 0.023 | 0.067 | 0.020 | 0.066 | 0.024 | 0.004 | 0.011 | 0.002 | 0.015 |
| Triglycerides | 183 | 0.143 | 0.061 | 0.132 | 0.081 | 0.153 | 0.061 | 0.145 | 0.069 | 0.010 | 0.052 | 0.012 | 0.056 |
| ***Very small VLDL*** | | | | | | | | | | | | | |
| Particles | 183 | 5.38e-05 | 1.14e-05 | 5.31e-05 | 1.42e-05 | 5.61e-05 | 1.23e-05 | 5.43e-05 | 1.62e-05 | 2.23e-06 | 7.32e-06 | 1.43e-06 | 8.91e-06 |
| Total lipids | 183 | 0.344 | 0.078 | 0.337 | 0.103 | 0.360 | 0.084 | 0.350 | 0.102 | 0.016 | 0.051 | 0.013 | 0.061 |
| Phospholipids | 183 | 0.097 | 0.025 | 0.094 | 0.033 | 0.101 | 0.026 | 0.097 | 0.034 | 0.005 | 0.017 | 0.004 | 0.020 |
| Cholesterol | 183 | 0.183 | 0.039 | 0.182 | 0.055 | 0.191 | 0.043 | 0.187 | 0.054 | 0.008 | 0.025 | 0.006 | 0.030 |
| Cholesteryl esters | 183 | 0.128 | 0.027 | 0.128 | 0.038 | 0.133 | 0.030 | 0.132 | 0.038 | 0.005 | 0.018 | 0.004 | 0.020 |
| Free cholesterol | 183 | 0.055 | 0.013 | 0.055 | 0.017 | 0.058 | 0.014 | 0.056 | 0.016 | 0.003 | 0.008 | 0.002 | 0.009 |
| Triglycerides | 183 | 0.064 | 0.020 | 0.062 | 0.023 | 0.067 | 0.020 | 0.063 | 0.022 | 0.003 | 0.016 | 0.003 | 0.018 |
| ***IDL*** | | | | | | | | | | | | | |
| Particles | 183 | 3.38e-04 | 6.06e-05 | 3.38e-04 | 7.60e-05 | 3.49e-04 | 6.41e-05 | 3.45e-04 | 8.60e-05 | 1.05e-05 | 4.28e-05 | 7.69e-06 | 4.78e-05 |
| Total lipids | 183 | 1.431 | 0.245 | 1.416 | 0.312 | 1.474 | 0.257 | 1.473 | 0.340 | 0.043 | 0.173 | 0.024 | 0.205 |
| Phospholipids | 183 | 0.327 | 0.058 | 0.327 | 0.079 | 0.338 | 0.061 | 0.338 | 0.083 | 0.011 | 0.039 | 0.008 | 0.047 |
| Cholesterol | 183 | 1.003 | 0.179 | 0.988 | 0.218 | 1.032 | 0.185 | 1.034 | 0.240 | 0.030 | 0.135 | 0.019 | 0.147 |
| Cholesteryl esters | 183 | 0.751 | 0.135 | 0.748 | 0.159 | 0.773 | 0.139 | 0.775 | 0.181 | 0.022 | 0.104 | 0.017 | 0.118 |
| Free cholesterol | 183 | 0.252 | 0.045 | 0.250 | 0.056 | 0.260 | 0.047 | 0.259 | 0.060 | 0.007 | 0.032 | 0.006 | 0.036 |
| Triglycerides | 183 | 0.101 | 0.023 | 0.098 | 0.025 | 0.104 | 0.023 | 0.098 | 0.027 | 0.003 | 0.018 | 0.003 | 0.019 |
| ***Large LDL*** | | | | | | | | | | | | | |
| Particles | 183 | 8.23e-04 | 1.73e-04 | 8.02e-04 | 2.12e-04 | 8.55e-04 | 1.75e-04 | 8.41e-04 | 2.40e-04 | 3.23e-05 | 1.00e-04 | 2.02e-05 | 1.15e-04 |
| Total lipids | 183 | 1.942 | 0.380 | 1.891 | 0.437 | 2.011 | 0.381 | 1.962 | 0.482 | 0.069 | 0.262 | 0.055 | 0.298 |
| Phospholipids | 183 | 0.427 | 0.079 | 0.419 | 0.090 | 0.440 | 0.079 | 0.427 | 0.102 | 0.013 | 0.055 | 0.009 | 0.065 |
| Cholesterol | 183 | 1.412 | 0.291 | 1.367 | 0.347 | 1.464 | 0.290 | 1.426 | 0.360 | 0.052 | 0.206 | 0.052 | 0.230 |
| Cholesteryl esters | 183 | 1.038 | 0.221 | 1.001 | 0.260 | 1.079 | 0.220 | 1.057 | 0.276 | 0.041 | 0.153 | 0.040 | 0.173 |
| Free cholesterol | 183 | 0.374 | 0.072 | 0.368 | 0.092 | 0.386 | 0.072 | 0.374 | 0.089 | 0.011 | 0.055 | 0.012 | 0.059 |
| Triglycerides | 183 | 0.103 | 0.021 | 0.100 | 0.024 | 0.106 | 0.021 | 0.101 | 0.027 | 0.003 | 0.016 | 0.003 | 0.017 |

| Metabolite | *n* | First measurement Mean | SD | Median | IQR | Last measurement Mean | SD | Median | IQR | Change (last -first measurement) Mean | SD | Median | IQR |
|---|---|---|---|---|---|---|---|---|---|---|---|---|---|
| ***Medium LDL*** | | | | | | | | | | | | | |
| Particles | 183 | 3.34e-04 | 8.07e-05 | 3.21e-04 | 9.50e-05 | 3.48e-04 | 8.65e-05 | 3.44e-04 | 1.02e-04 | 1.32e-05 | 4.75e-05 | 6.90e-06 | 5.74e-05 |
| Total lipids | 183 | 0.757 | 0.172 | 0.732 | 0.204 | 0.787 | 0.179 | 0.778 | 0.203 | 0.030 | 0.108 | 0.022 | 0.135 |
| Phospholipids | 183 | 0.199 | 0.043 | 0.192 | 0.047 | 0.206 | 0.044 | 0.203 | 0.052 | 0.007 | 0.027 | 0.006 | 0.032 |
| Cholesterol | 183 | 0.524 | 0.124 | 0.507 | 0.155 | 0.545 | 0.130 | 0.542 | 0.148 | 0.022 | 0.080 | 0.016 | 0.103 |
| Cholesteryl esters | 183 | 0.371 | 0.094 | 0.357 | 0.116 | 0.388 | 0.100 | 0.384 | 0.111 | 0.017 | 0.059 | 0.015 | 0.073 |
| Free cholesterol | 183 | 0.153 | 0.033 | 0.148 | 0.038 | 0.158 | 0.033 | 0.153 | 0.043 | 0.005 | 0.024 | 0.004 | 0.025 |
| Triglycerides | 183 | 0.034 | 0.008 | 0.033 | 0.009 | 0.036 | 0.008 | 0.034 | 0.008 | 0.001 | 0.006 | 0.002 | 0.006 |
| ***Small LDL*** | | | | | | | | | | | | | |
| Particles | 183 | 1.92e-04 | 3.61e-05 | 1.85e-04 | 3.90e-05 | 1.97e-04 | 3.79e-05 | 1.92e-04 | 4.50e-05 | 5.18e-06 | 2.26e-05 | 3.37e-06 | 2.60e-05 |
| Total lipids | 183 | 0.335 | 0.064 | 0.325 | 0.071 | 0.345 | 0.067 | 0.339 | 0.075 | 0.010 | 0.038 | 0.006 | 0.046 |
| Phospholipids | 183 | 0.104 | 0.019 | 0.102 | 0.021 | 0.106 | 0.019 | 0.104 | 0.020 | 0.002 | 0.010 | 0.002 | 0.012 |
| Cholesterol | 183 | 0.217 | 0.044 | 0.208 | 0.050 | 0.224 | 0.045 | 0.221 | 0.052 | 0.007 | 0.028 | 0.004 | 0.031 |
| Cholesteryl esters | 183 | 0.154 | 0.032 | 0.147 | 0.036 | 0.159 | 0.033 | 0.156 | 0.037 | 0.006 | 0.020 | 0.004 | 0.024 |
| Free cholesterol | 182 | 0.064 | 0.013 | 0.062 | 0.015 | 0.065 | 0.012 | 0.063 | 0.015 | 0.001 | 0.008 | 0.001 | 0.010 |
| Triglycerides | 183 | 0.014 | 0.004 | 0.013 | 0.005 | 0.015 | 0.004 | 0.014 | 0.004 | 0.001 | 0.004 | 0.001 | 0.003 |
| ***Very large HDL*** | | | | | | | | | | | | | |
| Particles | 183 | 3.12e-04 | 1.11e-04 | 2.95e-04 | 1.35e-04 | 3.05e-04 | 1.10e-04 | 2.81e-04 | 1.54e-04 | -6.87e-06 | 5.60e-05 | -8.89e-06 | 6.32e-05 |
| Total lipids | 183 | 0.223 | 0.095 | 0.212 | 0.115 | 0.216 | 0.094 | 0.198 | 0.135 | -0.008 | 0.048 | -0.009 | 0.056 |
| Phospholipids | 182 | 0.107 | 0.053 | 0.101 | 0.067 | 0.102 | 0.053 | 0.092 | 0.077 | -0.005 | 0.027 | -0.006 | 0.033 |
| Cholesterol | 183 | 0.110 | 0.041 | 0.104 | 0.046 | 0.107 | 0.040 | 0.099 | 0.054 | -0.003 | 0.021 | -0.003 | 0.023 |
| Cholesteryl esters | 183 | 0.085 | 0.033 | 0.080 | 0.037 | 0.082 | 0.032 | 0.075 | 0.044 | -0.003 | 0.016 | -0.002 | 0.017 |
| Free cholesterol | 183 | 0.026 | 0.009 | 0.025 | 0.010 | 0.025 | 0.009 | 0.024 | 0.011 | -0.001 | 0.005 | -0.001 | 0.006 |
| Triglycerides | 183 | 0.007 | 0.002 | 0.006 | 0.003 | 0.007 | 0.002 | 0.007 | 0.002 | 0.000 | 0.002 | 0.000 | 0.002 |
| ***Large HDL*** | | | | | | | | | | | | | |
| Particles | 183 | 2.21e-03 | 8.51e-04 | 2.10e-03 | 1.10e-03 | 2.16e-03 | 8.65e-04 | 2.01e-03 | 1.28e-03 | -5.22e-05 | 3.87e-04 | -6.89e-05 | 4.45e-04 |
| Total lipids | 183 | 0.970 | 0.355 | 0.931 | 0.459 | 0.947 | 0.363 | 0.893 | 0.535 | -0.023 | 0.163 | -0.030 | 0.195 |
| Phospholipids | 183 | 0.467 | 0.167 | 0.448 | 0.225 | 0.456 | 0.171 | 0.436 | 0.250 | -0.011 | 0.077 | -0.014 | 0.097 |
| Cholesterol | 183 | 0.475 | 0.186 | 0.457 | 0.248 | 0.462 | 0.188 | 0.434 | 0.279 | -0.014 | 0.086 | -0.016 | 0.103 |
| Cholesteryl esters | 183 | 0.372 | 0.144 | 0.358 | 0.192 | 0.361 | 0.145 | 0.343 | 0.222 | -0.011 | 0.066 | -0.013 | 0.078 |
| Free cholesterol | 183 | 0.103 | 0.042 | 0.099 | 0.051 | 0.100 | 0.043 | 0.093 | 0.063 | -0.003 | 0.020 | -0.003 | 0.024 |
| Triglycerides | 183 | 0.028 | 0.011 | 0.026 | 0.012 | 0.029 | 0.012 | 0.027 | 0.012 | 0.002 | 0.010 | 0.001 | 0.011 |
| ***Medium HDL*** | | | | | | | | | | | | | |
| Particles | 183 | 4.86e-03 | 8.49e-04 | 4.87e-03 | 1.07e-03 | 4.90e-03 | 8.80e-04 | 4.90e-03 | 1.14e-03 | 3.92e-05 | 5.43e-04 | -3.32e-05 | 7.03e-04 |
| Total lipids | 183 | 1.251 | 0.202 | 1.250 | 0.261 | 1.262 | 0.208 | 1.259 | 0.255 | 0.012 | 0.135 | 0.006 | 0.172 |
| Phospholipids | 183 | 0.565 | 0.087 | 0.562 | 0.112 | 0.572 | 0.090 | 0.571 | 0.115 | 0.007 | 0.063 | 0.007 | 0.081 |
| Cholesterol | 183 | 0.644 | 0.117 | 0.642 | 0.148 | 0.644 | 0.120 | 0.639 | 0.164 | 0.001 | 0.070 | -0.009 | 0.091 |
| Cholesteryl esters | 183 | 0.528 | 0.095 | 0.527 | 0.120 | 0.528 | 0.096 | 0.526 | 0.132 | 0.000 | 0.057 | -0.008 | 0.071 |
| Free cholesterol | 183 | 0.116 | 0.023 | 0.115 | 0.030 | 0.117 | 0.024 | 0.116 | 0.032 | 0.001 | 0.014 | -0.002 | 0.019 |
| Triglycerides | 183 | 0.042 | 0.018 | 0.040 | 0.024 | 0.046 | 0.018 | 0.044 | 0.020 | 0.004 | 0.017 | 0.003 | 0.019 |

| Metabolite | *n* | First measurement Mean | SD | Median | IQR | Last measurement Mean | SD | Median | IQR | Change (last -first measurement) Mean | SD | Median | IQR |
|---|---|---|---|---|---|---|---|---|---|---|---|---|---|
| ***Small HDL*** | | | | | | | | | | | | | |
| Particles | 183 | 1.11e-02 | 1.31e-03 | 1.10e-02 | 1.90e-03 | 1.13e-02 | 1.29e-03 | 1.13e-02 | 1.70e-03 | 0.000 | 0.001 | 0.000 | 0.001 |
| Total lipids | 183 | 1.277 | 0.149 | 1.275 | 0.216 | 1.306 | 0.145 | 1.318 | 0.178 | 0.028 | 0.127 | 0.035 | 0.158 |
| Phospholipids | 183 | 0.716 | 0.086 | 0.707 | 0.122 | 0.732 | 0.083 | 0.736 | 0.101 | 0.017 | 0.074 | 0.018 | 0.091 |
| Cholesterol | 183 | 0.520 | 0.059 | 0.517 | 0.084 | 0.528 | 0.058 | 0.528 | 0.077 | 0.009 | 0.051 | 0.011 | 0.067 |
| Cholesteryl esters | 183 | 0.384 | 0.046 | 0.383 | 0.069 | 0.390 | 0.046 | 0.391 | 0.059 | 0.006 | 0.041 | 0.009 | 0.051 |
| Free cholesterol | 183 | 0.136 | 0.014 | 0.135 | 0.019 | 0.138 | 0.015 | 0.138 | 0.019 | 0.003 | 0.012 | 0.003 | 0.016 |
| Triglycerides | 183 | 0.042 | 0.017 | 0.042 | 0.020 | 0.046 | 0.017 | 0.044 | 0.020 | 0.003 | 0.013 | 0.003 | 0.015 |

**Result Supplement Table 2.** Descriptive statistics for metabolite measures among women starting menopausal hormone therapy (*n* = 35) during the study.

| | | First measurement | | | | Last measurement | | | | Change (last – first measurement) | | | |
|---|---|---|---|---|---|---|---|---|---|---|---|---|---|
| **Metabolite** | ***n*** | **Mean** | **SD** | **Median** | **IQR** | **Mean** | **SD** | **Median** | **IQR** | **Mean** | **SD** | **Median** | **IQR** |
| ***Cholesterols*** | | | | | | | | | | | | | |
| Total cholesterol | 35 | 5.509 | 0.786 | 5.488 | 0.829 | 5.534 | 0.807 | 5.556 | 0.788 | 0.025 | 0.537 | 0.067 | 0.575 |
| Non-HDL cholesterol | 35 | 3.853 | 0.779 | 3.863 | 0.862 | 3.773 | 0.756 | 3.745 | 0.786 | -0.080 | 0.562 | -0.085 | 0.435 |
| Remnant cholesterol | 35 | 1.675 | 0.356 | 1.627 | 0.335 | 1.665 | 0.339 | 1.644 | 0.323 | -0.009 | 0.279 | 0.015 | 0.234 |
| VLDL cholesterol | 35 | 0.671 | 0.184 | 0.637 | 0.231 | 0.664 | 0.194 | 0.646 | 0.242 | -0.006 | 0.143 | 0.009 | 0.155 |
| Clinical LDL cholesterol | 35 | 3.217 | 0.715 | 3.213 | 0.781 | 3.075 | 0.687 | 3.047 | 0.656 | -0.142 | 0.519 | -0.071 | 0.541 |
| LDL cholesterol | 35 | 2.178 | 0.439 | 2.170 | 0.582 | 2.108 | 0.433 | 2.136 | 0.481 | -0.071 | 0.296 | -0.034 | 0.345 |
| HDL cholesterol | 35 | 1.656 | 0.274 | 1.644 | 0.308 | 1.761 | 0.280 | 1.689 | 0.419 | 0.105 | 0.182 | 0.133 | 0.201 |
| ***Triglycerides*** | | | | | | | | | | | | | |
| Total triglycerides | 35 | 1.046 | 0.341 | 0.993 | 0.420 | 1.194 | 0.493 | 1.064 | 0.546 | 0.147 | 0.340 | 0.090 | 0.277 |
| VLDL triglycerides | 35 | 0.697 | 0.312 | 0.623 | 0.367 | 0.789 | 0.424 | 0.657 | 0.500 | 0.092 | 0.276 | 0.055 | 0.211 |
| LDL triglycerides | 35 | 0.145 | 0.019 | 0.139 | 0.022 | 0.158 | 0.032 | 0.150 | 0.037 | 0.014 | 0.027 | 0.007 | 0.035 |
| HDL triglycerides | 35 | 0.108 | 0.027 | 0.103 | 0.033 | 0.139 | 0.045 | 0.134 | 0.054 | 0.031 | 0.045 | 0.022 | 0.061 |
| ***Phospholipids*** | | | | | | | | | | | | | |
| Total phospholipids | 35 | 3.222 | 0.300 | 3.202 | 0.376 | 3.389 | 0.402 | 3.457 | 0.530 | 0.167 | 0.306 | 0.177 | 0.408 |
| VLDL phospholipids | 35 | 0.410 | 0.121 | 0.408 | 0.148 | 0.424 | 0.145 | 0.411 | 0.176 | 0.014 | 0.095 | -0.003 | 0.096 |
| LDL phospholipids | 35 | 0.736 | 0.136 | 0.739 | 0.162 | 0.712 | 0.134 | 0.711 | 0.148 | -0.025 | 0.097 | -0.020 | 0.098 |
| HDL phospholipids | 35 | 1.746 | 0.234 | 1.704 | 0.302 | 1.925 | 0.283 | 1.893 | 0.458 | 0.179 | 0.262 | 0.166 | 0.282 |
| ***Cholesteryl esters*** | | | | | | | | | | | | | |
| Total cholesteryl esters | 35 | 4.049 | 0.563 | 4.052 | 0.659 | 4.070 | 0.586 | 4.095 | 0.556 | 0.021 | 0.379 | 0.036 | 0.454 |
| VLDL cholesteryl esters | 35 | 0.413 | 0.112 | 0.393 | 0.133 | 0.403 | 0.113 | 0.387 | 0.142 | -0.011 | 0.089 | 0.005 | 0.097 |
| LDL cholesteryl esters | 35 | 1.578 | 0.322 | 1.581 | 0.445 | 1.533 | 0.322 | 1.555 | 0.392 | -0.046 | 0.211 | -0.026 | 0.240 |
| HDL cholesteryl esters | 35 | 1.298 | 0.214 | 1.301 | 0.251 | 1.372 | 0.217 | 1.315 | 0.315 | 0.075 | 0.141 | 0.087 | 0.156 |
| ***Free cholesterol*** | | | | | | | | | | | | | |
| Total free cholesterol | 35 | 1.460 | 0.227 | 1.448 | 0.220 | 1.464 | 0.224 | 1.469 | 0.209 | 0.004 | 0.162 | 0.022 | 0.148 |
| VLDL free cholesterol | 35 | 0.257 | 0.076 | 0.251 | 0.099 | 0.262 | 0.085 | 0.249 | 0.106 | 0.004 | 0.057 | 0.000 | 0.054 |
| LDL free cholesterol | 35 | 0.600 | 0.119 | 0.603 | 0.132 | 0.575 | 0.115 | 0.580 | 0.108 | -0.025 | 0.087 | -0.008 | 0.097 |
| HDL free cholesterol | 35 | 0.359 | 0.061 | 0.354 | 0.067 | 0.389 | 0.065 | 0.389 | 0.108 | 0.030 | 0.044 | 0.035 | 0.058 |
| ***Total lipids*** | | | | | | | | | | | | | |
| Total lipids | 35 | 9.777 | 1.150 | 9.681 | 1.211 | 10.116 | 1.387 | 10.190 | 1.406 | 0.339 | 0.986 | 0.255 | 1.379 |
| VLDL total lipids | 35 | 1.778 | 0.557 | 1.714 | 0.699 | 1.878 | 0.715 | 1.779 | 0.832 | 0.100 | 0.447 | 0.072 | 0.435 |
| LDL total lipids | 35 | 3.059 | 0.586 | 3.067 | 0.767 | 2.978 | 0.581 | 2.983 | 0.681 | -0.082 | 0.402 | -0.067 | 0.414 |
| HDL total lipids | 35 | 3.511 | 0.494 | 3.412 | 0.582 | 3.825 | 0.558 | 3.844 | 0.888 | 0.314 | 0.463 | 0.321 | 0.533 |
| ***Particles*** | | | | | | | | | | | | | |
| Total particles | 35 | 0.020 | 0.002 | 0.020 | 0.002 | 0.021 | 0.002 | 0.021 | 0.003 | 0.001 | 0.002 | 0.001 | 0.002 |
| VLDL particles | 35 | 1.37e-04 | 3.05e-05 | 1.33e-04 | 3.70e-05 | 1.41e-04 | 3.66e-05 | 1.35e-04 | 4.20e-05 | 4.54e-06 | 2.58e-05 | 1.33e-06 | 2.25e-05 |
| LDL particles | 35 | 1.37e-03 | 2.74e-04 | 1.35e-03 | 3.70e-04 | 1.31e-03 | 2.63e-04 | 1.34e-03 | 3.20e-04 | -6.06e-05 | 1.96e-04 | -6.16e-05 | 1.75e-04 |
| HDL particles | 35 | 0.018 | 0.002 | 0.018 | 0.002 | 0.019 | 0.002 | 0.019 | 0.003 | 0.001 | 0.002 | 0.001 | 0.002 |

| Metabolite | *n* | First measurement Mean | SD | Median | IQR | Last measurement Mean | SD | Median | IQR | Change (last -first measurement) Mean | SD | Median | IQR |
|---|---|---|---|---|---|---|---|---|---|---|---|---|---|
| ***Particle diameter*** | | | | | | | | | | | | | |
| VLDL size | 35 | 38.140 | 1.016 | 38.167 | 1.135 | 38.146 | 1.162 | 37.949 | 1.547 | 0.005 | 0.614 | -0.054 | 0.880 |
| LDL size | 35 | 23.919 | 0.074 | 23.907 | 0.083 | 23.938 | 0.055 | 23.933 | 0.083 | 0.019 | 0.089 | 0.020 | 0.087 |
| HDL size | 35 | 9.736 | 0.189 | 9.712 | 0.230 | 9.780 | 0.175 | 9.715 | 0.198 | 0.044 | 0.093 | 0.040 | 0.125 |
| ***Phosphoglycerides*** | | | | | | | | | | | | | |
| Phosphoglycerides | 35 | 2.693 | 0.215 | 2.671 | 0.273 | 2.819 | 0.329 | 2.852 | 0.447 | 0.126 | 0.275 | 0.112 | 0.348 |
| Tri-/phosphoglyceride ratio | 35 | 0.390 | 0.127 | 0.378 | 0.159 | 0.420 | 0.147 | 0.380 | 0.157 | 0.030 | 0.088 | 0.013 | 0.112 |
| Total cholines | 35 | 3.050 | 0.241 | 3.001 | 0.319 | 3.145 | 0.325 | 3.186 | 0.445 | 0.095 | 0.255 | 0.099 | 0.353 |
| Phosphatidylcholines | 35 | 2.549 | 0.207 | 2.525 | 0.273 | 2.691 | 0.323 | 2.710 | 0.428 | 0.142 | 0.282 | 0.126 | 0.379 |
| Sphingomyelins | 35 | 0.537 | 0.058 | 0.538 | 0.085 | 0.538 | 0.060 | 0.542 | 0.072 | 0.002 | 0.031 | 0.007 | 0.030 |
| ***Apolipoproteins*** | | | | | | | | | | | | | |
| Apolipoprotein B | 35 | 0.946 | 0.185 | 0.925 | 0.222 | 0.915 | 0.177 | 0.930 | 0.212 | -0.031 | 0.136 | -0.027 | 0.116 |
| Apolipoprotein A-I | 35 | 1.655 | 0.170 | 1.645 | 0.240 | 1.780 | 0.207 | 1.777 | 0.309 | 0.125 | 0.189 | 0.132 | 0.210 |
| Apo B/A-I ratio | 35 | 0.581 | 0.145 | 0.522 | 0.246 | 0.521 | 0.120 | 0.516 | 0.177 | -0.060 | 0.124 | -0.033 | 0.151 |
| ***Fatty acids*** | | | | | | | | | | | | | |
| Total fatty acids | 35 | 12.941 | 1.297 | 12.743 | 1.235 | 13.337 | 1.957 | 13.017 | 2.318 | 0.396 | 1.327 | 0.187 | 1.515 |
| Degree of unsaturation | 35 | 1.356 | 0.059 | 1.358 | 0.056 | 1.356 | 0.062 | 1.352 | 0.072 | 0.000 | 0.043 | 0.005 | 0.061 |
| Omega-3 fatty acids | 35 | 0.611 | 0.146 | 0.609 | 0.224 | 0.650 | 0.188 | 0.623 | 0.171 | 0.039 | 0.143 | 0.041 | 0.164 |
| Omega-6 fatty acids | 35 | 5.083 | 0.439 | 5.100 | 0.571 | 5.057 | 0.515 | 5.049 | 0.633 | -0.027 | 0.376 | 0.053 | 0.426 |
| Polyunsaturated fatty acids | 35 | 5.694 | 0.539 | 5.723 | 0.835 | 5.707 | 0.626 | 5.658 | 0.796 | 0.013 | 0.422 | 0.096 | 0.531 |
| Monounsaturated fatty acids | 35 | 3.048 | 0.461 | 2.940 | 0.474 | 3.140 | 0.695 | 2.947 | 0.710 | 0.093 | 0.443 | -0.007 | 0.507 |
| Saturated fatty acids | 35 | 4.199 | 0.467 | 4.152 | 0.552 | 4.490 | 0.827 | 4.323 | 0.895 | 0.291 | 0.644 | 0.178 | 0.672 |
| Linoleic acid | 35 | 4.524 | 0.457 | 4.516 | 0.461 | 4.296 | 0.578 | 4.167 | 0.890 | -0.228 | 0.545 | -0.133 | 0.651 |
| Docosahexaenoic acid | 35 | 0.318 | 0.056 | 0.310 | 0.072 | 0.329 | 0.069 | 0.323 | 0.058 | 0.012 | 0.058 | 0.011 | 0.062 |
| ***Fatty acid ratios (%)*** | | | | | | | | | | | | | |
| Omega-3 ratio | 35 | 4.703 | 0.958 | 4.717 | 1.036 | 4.859 | 1.080 | 4.701 | 1.161 | 0.156 | 0.928 | 0.157 | 0.797 |
| Omega-6 ratio | 35 | 39.380 | 2.013 | 39.740 | 2.641 | 38.230 | 3.011 | 39.123 | 3.479 | -1.149 | 2.314 | -1.097 | 2.377 |
| Polyunsaturated ratio | 35 | 44.083 | 2.324 | 44.381 | 2.930 | 43.089 | 3.171 | 43.398 | 3.145 | -0.993 | 2.350 | -0.688 | 2.195 |
| Monounsaturated ratio | 35 | 23.487 | 1.837 | 23.510 | 1.946 | 23.354 | 2.220 | 23.118 | 2.934 | -0.133 | 1.399 | -0.205 | 1.744 |
| Saturated ratio | 35 | 32.430 | 1.136 | 32.488 | 1.110 | 33.557 | 1.996 | 33.221 | 2.757 | 1.127 | 2.032 | 0.713 | 2.623 |
| Linoleic acid ratio | 35 | 35.038 | 2.410 | 35.489 | 3.018 | 32.482 | 3.758 | 32.657 | 4.272 | -2.555 | 3.764 | -2.013 | 5.244 |
| Docosahexaenoic acid ratio | 35 | 2.461 | 0.405 | 2.468 | 0.523 | 2.491 | 0.478 | 2.411 | 0.451 | 0.030 | 0.436 | 0.027 | 0.435 |
| Polyu/monounsaturated ratio | 35 | 1.894 | 0.229 | 1.878 | 0.276 | 1.871 | 0.289 | 1.885 | 0.336 | -0.023 | 0.183 | -0.018 | 0.247 |
| Omega-6 to omega-3 ratio | 35 | 8.722 | 1.880 | 8.320 | 2.380 | 8.229 | 1.845 | 8.100 | 1.862 | -0.493 | 1.777 | -0.787 | 1.691 |
| ***Amino acids*** | | | | | | | | | | | | | |
| Alanine | 35 | 0.371 | 0.072 | 0.359 | 0.067 | 0.377 | 0.063 | 0.370 | 0.071 | 0.006 | 0.062 | 0.014 | 0.061 |
| Glutamine | 35 | 0.759 | 0.076 | 0.732 | 0.090 | 0.683 | 0.078 | 0.693 | 0.121 | -0.076 | 0.095 | -0.074 | 0.123 |
| Glycine | 35 | 0.317 | 0.071 | 0.302 | 0.108 | 0.296 | 0.073 | 0.277 | 0.084 | -0.021 | 0.045 | -0.017 | 0.054 |
| Histidine | 35 | 0.077 | 0.012 | 0.075 | 0.014 | 0.081 | 0.009 | 0.079 | 0.013 | 0.003 | 0.011 | 0.004 | 0.011 |
| Total BCAA | 35 | 0.394 | 0.062 | 0.374 | 0.062 | 0.396 | 0.063 | 0.388 | 0.054 | 0.003 | 0.064 | 0.005 | 0.070 |
| Isoleucine | 35 | 0.052 | 0.011 | 0.050 | 0.012 | 0.051 | 0.011 | 0.048 | 0.011 | -0.001 | 0.011 | 0.000 | 0.010 |
| Leucine | 35 | 0.112 | 0.019 | 0.109 | 0.020 | 0.113 | 0.019 | 0.114 | 0.021 | 0.001 | 0.020 | 0.001 | 0.022 |
| Valine | 35 | 0.230 | 0.035 | 0.220 | 0.034 | 0.233 | 0.035 | 0.228 | 0.030 | 0.003 | 0.037 | 0.004 | 0.044 |

| | | First measurement | | | | Last measurement | | | | Change (last -first measurement) | | | |
|---|---|---|---|---|---|---|---|---|---|---|---|---|---|
| **Metabolite** | ***n*** | **Mean** | **SD** | **Median** | **IQR** | **Mean** | **SD** | **Median** | **IQR** | **Mean** | **SD** | **Median** | **IQR** |
| ***Amino acids continue*** | | | | | | | | | | | | | |
| Phenylalanine | 35 | 0.064 | 0.008 | 0.064 | 0.009 | 0.066 | 0.008 | 0.065 | 0.011 | 0.002 | 0.009 | 0.004 | 0.012 |
| Tyrosine | 35 | 0.063 | 0.010 | 0.061 | 0.013 | 0.063 | 0.009 | 0.062 | 0.010 | 0.000 | 0.011 | -0.001 | 0.015 |
| ***Glycolysis-related*** | | | | | | | | | | | | | |
| Glucose | 35 | 5.221 | 0.490 | 5.180 | 0.484 | 5.122 | 0.536 | 5.105 | 0.644 | -0.099 | 0.452 | -0.129 | 0.607 |
| Lactate | 35 | 1.219 | 0.220 | 1.232 | 0.263 | 1.406 | 0.471 | 1.299 | 0.492 | 0.188 | 0.465 | 0.113 | 0.441 |
| Pyruvate | 35 | 0.050 | 0.014 | 0.048 | 0.018 | 0.059 | 0.025 | 0.059 | 0.032 | 0.009 | 0.019 | 0.002 | 0.022 |
| Citrate | 35 | 0.062 | 0.009 | 0.061 | 0.009 | 0.059 | 0.008 | 0.059 | 0.008 | -0.003 | 0.008 | -0.003 | 0.008 |
| Glycerol | 35 | 0.113 | 0.047 | 0.107 | 0.038 | 0.132 | 0.064 | 0.125 | 0.059 | 0.019 | 0.044 | 0.007 | 0.054 |
| ***Ketone bodies*** | | | | | | | | | | | | | |
| 3-Hydroxybutyrate | 35 | 0.056 | 0.053 | 0.033 | 0.074 | 0.023 | 0.025 | 0.015 | 0.020 | -0.033 | 0.055 | -0.020 | 0.071 |
| Acetate | 35 | 0.027 | 0.013 | 0.025 | 0.011 | 0.029 | 0.019 | 0.026 | 0.018 | 0.002 | 0.020 | 0.000 | 0.015 |
| Acetoacetate | 35 | 0.031 | 0.018 | 0.025 | 0.023 | 0.022 | 0.011 | 0.017 | 0.014 | -0.009 | 0.019 | -0.007 | 0.023 |
| Acetone | 35 | 0.018 | 0.006 | 0.015 | 0.007 | 0.016 | 0.004 | 0.015 | 0.004 | -0.002 | 0.005 | -0.001 | 0.006 |
| ***Miscellaneous*** | | | | | | | | | | | | | |
| Creatinine | 35 | 72.348 | 8.354 | 72.368 | 12.359 | 73.196 | 10.579 | 71.884 | 14.107 | 0.848 | 6.970 | 1.963 | 9.742 |
| Albumin | 35 | 42.263 | 2.821 | 42.945 | 3.608 | 41.293 | 2.734 | 41.748 | 4.456 | -0.969 | 2.152 | -0.546 | 3.195 |
| Glycoprotein acetyls | 35 | 0.822 | 0.074 | 0.823 | 0.108 | 0.816 | 0.086 | 0.802 | 0.120 | -0.006 | 0.060 | -0.006 | 0.070 |
| ***Chylomicrons and extremely large VLDL*** | | | | | | | | | | | | | |
| Particles | 35 | 3.69e-07 | 6.92e-07 | 2.91e-08 | 4.93e-07 | 5.92e-07 | 1.02e-06 | 5.90e-08 | 6.58e-07 | 2.23e-07 | 7.30e-07 | 3.38e-09 | 1.33e-07 |
| Total lipids | 35 | 0.051 | 0.097 | 0.014 | 0.049 | 0.080 | 0.138 | 0.015 | 0.085 | 0.028 | 0.097 | 0.000 | 0.022 |
| Phospholipids | 35 | 0.006 | 0.012 | 0.000 | 0.006 | 0.010 | 0.019 | 0.000 | 0.010 | 0.004 | 0.013 | 0.000 | 0.001 |
| Cholesterol | 35 | 0.019 | 0.019 | 0.013 | 0.020 | 0.023 | 0.026 | 0.013 | 0.025 | 0.004 | 0.015 | 0.000 | 0.012 |
| Cholesteryl esters | 35 | 0.014 | 0.011 | 0.012 | 0.013 | 0.015 | 0.015 | 0.012 | 0.016 | 0.001 | 0.008 | -0.001 | 0.008 |
| Free cholesterol | 35 | 0.005 | 0.008 | 0.001 | 0.006 | 0.008 | 0.012 | 0.002 | 0.008 | 0.003 | 0.008 | 0.001 | 0.001 |
| Triglycerides | 35 | 0.026 | 0.067 | 0.001 | 0.014 | 0.047 | 0.095 | 0.001 | 0.054 | 0.021 | 0.071 | 0.000 | 0.009 |
| ***Very large VLDL*** | | | | | | | | | | | | | |
| Particles | 35 | 2.30e-06 | 1.55e-06 | 2.16e-06 | 1.97e-06 | 2.64e-06 | 1.99e-06 | 2.16e-06 | 2.15e-06 | 3.42e-07 | 1.21e-06 | 1.44e-07 | 1.07e-06 |
| Total lipids | 35 | 0.134 | 0.094 | 0.118 | 0.112 | 0.150 | 0.120 | 0.130 | 0.130 | 0.017 | 0.073 | -0.002 | 0.071 |
| Phospholipids | 35 | 0.022 | 0.017 | 0.019 | 0.023 | 0.025 | 0.022 | 0.021 | 0.023 | 0.003 | 0.013 | 0.002 | 0.013 |
| Cholesterol | 35 | 0.041 | 0.019 | 0.041 | 0.027 | 0.041 | 0.023 | 0.041 | 0.030 | 0.000 | 0.013 | 0.000 | 0.016 |
| Cholesteryl esters | 35 | 0.027 | 0.010 | 0.027 | 0.015 | 0.025 | 0.012 | 0.025 | 0.017 | -0.001 | 0.007 | -0.001 | 0.009 |
| Free cholesterol | 35 | 0.014 | 0.010 | 0.013 | 0.013 | 0.016 | 0.012 | 0.014 | 0.012 | 0.001 | 0.007 | 0.001 | 0.007 |
| Triglycerides | 35 | 0.070 | 0.059 | 0.058 | 0.064 | 0.084 | 0.076 | 0.056 | 0.085 | 0.014 | 0.049 | 0.001 | 0.047 |
| ***Large VLDL*** | | | | | | | | | | | | | |
| Particles | 35 | 8.28e-06 | 3.91e-06 | 7.85e-06 | 5.80e-06 | 9.02e-06 | 5.00e-06 | 8.09e-06 | 6.21e-06 | 7.38e-07 | 2.96e-06 | 5.52e-07 | 3.33e-06 |
| Total lipids | 35 | 0.274 | 0.135 | 0.259 | 0.197 | 0.294 | 0.170 | 0.275 | 0.205 | 0.020 | 0.101 | 0.001 | 0.112 |
| Phospholipids | 35 | 0.051 | 0.028 | 0.049 | 0.042 | 0.055 | 0.035 | 0.050 | 0.044 | 0.005 | 0.020 | 0.002 | 0.024 |
| Cholesterol | 35 | 0.078 | 0.035 | 0.079 | 0.049 | 0.081 | 0.041 | 0.079 | 0.054 | 0.003 | 0.024 | 0.003 | 0.031 |
| Cholesteryl esters | 35 | 0.042 | 0.017 | 0.043 | 0.024 | 0.043 | 0.020 | 0.041 | 0.028 | 0.000 | 0.012 | 0.000 | 0.014 |
| Free cholesterol | 35 | 0.036 | 0.018 | 0.036 | 0.025 | 0.039 | 0.022 | 0.035 | 0.026 | 0.003 | 0.012 | 0.001 | 0.015 |
| Triglycerides | 35 | 0.145 | 0.074 | 0.126 | 0.091 | 0.158 | 0.096 | 0.138 | 0.112 | 0.013 | 0.061 | -0.005 | 0.065 |

| | | First measurement | | | | Last measurement | | | | Change (last -first measurement) | | | |
|---|---|---|---|---|---|---|---|---|---|---|---|---|---|
| **Metabolite** | ***n*** | **Mean** | **SD** | **Median** | **IQR** | **Mean** | **SD** | **Median** | **IQR** | **Mean** | **SD** | **Median** | **IQR** |
| ***Medium VLDL*** | | | | | | | | | | | | | |
| Particles | 35 | 3.58e-05 | 9.60e-06 | 3.54e-05 | 1.32e-05 | 3.54e-05 | 1.05e-05 | 3.41e-05 | 1.68e-05 | -4.05e-07 | 6.68e-06 | 5.61e-07 | 7.87e-06 |
| Total lipids | 35 | 0.593 | 0.160 | 0.597 | 0.190 | 0.595 | 0.187 | 0.563 | 0.287 | 0.002 | 0.113 | 0.005 | 0.138 |
| Phospholipids | 35 | 0.136 | 0.038 | 0.134 | 0.050 | 0.134 | 0.041 | 0.130 | 0.063 | -0.003 | 0.028 | 0.001 | 0.030 |
| Cholesterol | 35 | 0.192 | 0.058 | 0.188 | 0.064 | 0.180 | 0.056 | 0.176 | 0.065 | -0.012 | 0.046 | -0.009 | 0.041 |
| Cholesteryl esters | 35 | 0.108 | 0.035 | 0.108 | 0.037 | 0.098 | 0.033 | 0.097 | 0.028 | -0.010 | 0.029 | -0.006 | 0.025 |
| Free cholesterol | 35 | 0.084 | 0.024 | 0.080 | 0.027 | 0.081 | 0.025 | 0.081 | 0.036 | -0.002 | 0.018 | 0.000 | 0.016 |
| Triglycerides | 35 | 0.265 | 0.088 | 0.245 | 0.124 | 0.281 | 0.114 | 0.263 | 0.145 | 0.016 | 0.069 | 0.010 | 0.077 |
| ***Small VLDL*** | | | | | | | | | | | | | |
| Particles | 35 | 3.65e-05 | 8.22e-06 | 3.67e-05 | 1.20e-05 | 3.81e-05 | 1.04e-05 | 3.66e-05 | 1.24e-05 | 1.55e-06 | 7.16e-06 | 1.70e-06 | 7.71e-06 |
| Total lipids | 35 | 0.387 | 0.086 | 0.378 | 0.120 | 0.401 | 0.108 | 0.388 | 0.132 | 0.014 | 0.074 | 0.018 | 0.091 |
| Phospholipids | 35 | 0.100 | 0.023 | 0.099 | 0.032 | 0.099 | 0.026 | 0.099 | 0.035 | -0.001 | 0.018 | 0.001 | 0.020 |
| Cholesterol | 35 | 0.156 | 0.041 | 0.142 | 0.051 | 0.152 | 0.043 | 0.148 | 0.057 | -0.004 | 0.034 | 0.003 | 0.035 |
| Cholesteryl esters | 35 | 0.092 | 0.025 | 0.086 | 0.034 | 0.090 | 0.027 | 0.088 | 0.037 | -0.002 | 0.022 | 0.001 | 0.023 |
| Free cholesterol | 35 | 0.064 | 0.016 | 0.061 | 0.021 | 0.061 | 0.016 | 0.060 | 0.022 | -0.002 | 0.012 | 0.000 | 0.012 |
| Triglycerides | 35 | 0.131 | 0.036 | 0.120 | 0.043 | 0.150 | 0.051 | 0.136 | 0.057 | 0.019 | 0.037 | 0.011 | 0.041 |
| ***Very small VLDL*** | | | | | | | | | | | | | |
| Particles | 35 | 5.32e-05 | 1.11e-05 | 5.06e-05 | 1.12e-05 | 5.53e-05 | 1.18e-05 | 5.30e-05 | 1.03e-05 | 2.10e-06 | 1.01e-05 | 2.28e-06 | 1.03e-05 |
| Total lipids | 35 | 0.339 | 0.074 | 0.324 | 0.085 | 0.357 | 0.079 | 0.343 | 0.075 | 0.019 | 0.068 | 0.020 | 0.075 |
| Phospholipids | 35 | 0.095 | 0.023 | 0.088 | 0.026 | 0.101 | 0.025 | 0.098 | 0.023 | 0.006 | 0.021 | 0.008 | 0.019 |
| Cholesterol | 35 | 0.184 | 0.045 | 0.173 | 0.041 | 0.187 | 0.041 | 0.187 | 0.029 | 0.003 | 0.036 | 0.004 | 0.034 |
| Cholesteryl esters | 35 | 0.129 | 0.032 | 0.124 | 0.029 | 0.131 | 0.029 | 0.132 | 0.021 | 0.001 | 0.025 | 0.001 | 0.022 |
| Free cholesterol | 35 | 0.055 | 0.013 | 0.052 | 0.015 | 0.057 | 0.013 | 0.055 | 0.010 | 0.002 | 0.011 | 0.003 | 0.013 |
| Triglycerides | 35 | 0.060 | 0.011 | 0.058 | 0.012 | 0.069 | 0.019 | 0.066 | 0.017 | 0.009 | 0.016 | 0.006 | 0.023 |
| ***IDL*** | | | | | | | | | | | | | |
| Particles | 35 | 3.38e-04 | 6.88e-05 | 3.29e-04 | 6.50e-05 | 3.33e-04 | 6.12e-05 | 3.32e-04 | 5.50e-05 | -4.94e-06 | 5.45e-05 | -5.84e-06 | 4.71e-05 |
| Total lipids | 35 | 1.429 | 0.270 | 1.420 | 0.274 | 1.436 | 0.254 | 1.462 | 0.246 | 0.007 | 0.203 | 0.011 | 0.181 |
| Phospholipids | 35 | 0.330 | 0.065 | 0.323 | 0.061 | 0.328 | 0.060 | 0.330 | 0.043 | -0.001 | 0.047 | 0.003 | 0.037 |
| Cholesterol | 35 | 1.004 | 0.199 | 1.005 | 0.207 | 1.001 | 0.185 | 1.040 | 0.195 | -0.003 | 0.150 | 0.000 | 0.144 |
| Cholesteryl esters | 35 | 0.751 | 0.148 | 0.751 | 0.150 | 0.752 | 0.140 | 0.773 | 0.158 | 0.002 | 0.117 | 0.006 | 0.107 |
| Free cholesterol | 35 | 0.253 | 0.052 | 0.250 | 0.053 | 0.249 | 0.046 | 0.258 | 0.040 | -0.004 | 0.035 | -0.001 | 0.032 |
| Triglycerides | 35 | 0.096 | 0.014 | 0.090 | 0.016 | 0.107 | 0.023 | 0.104 | 0.024 | 0.011 | 0.021 | 0.008 | 0.025 |
| ***Large LDL*** | | | | | | | | | | | | | |
| Particles | 35 | 8.39e-04 | 1.79e-04 | 8.07e-04 | 2.17e-04 | 8.06e-04 | 1.62e-04 | 8.22e-04 | 1.69e-04 | -3.28e-05 | 1.40e-04 | -1.89e-05 | 9.50e-05 |
| Total lipids | 35 | 1.955 | 0.380 | 1.929 | 0.421 | 1.922 | 0.369 | 1.908 | 0.396 | -0.034 | 0.271 | -0.005 | 0.286 |
| Phospholipids | 35 | 0.431 | 0.079 | 0.431 | 0.078 | 0.419 | 0.077 | 0.417 | 0.085 | -0.012 | 0.058 | -0.010 | 0.058 |
| Cholesterol | 35 | 1.425 | 0.294 | 1.401 | 0.343 | 1.394 | 0.282 | 1.401 | 0.286 | -0.031 | 0.207 | 0.005 | 0.236 |
| Cholesteryl esters | 35 | 1.047 | 0.220 | 1.044 | 0.288 | 1.027 | 0.213 | 1.032 | 0.205 | -0.020 | 0.154 | 0.005 | 0.176 |
| Free cholesterol | 35 | 0.379 | 0.075 | 0.376 | 0.076 | 0.367 | 0.071 | 0.368 | 0.068 | -0.011 | 0.055 | 0.001 | 0.058 |
| Triglycerides | 35 | 0.099 | 0.013 | 0.094 | 0.016 | 0.108 | 0.021 | 0.105 | 0.029 | 0.009 | 0.019 | 0.006 | 0.025 |

| | | First measurement | | | | Last measurement | | | | Change (last -first measurement) | | | |
|---|---|---|---|---|---|---|---|---|---|---|---|---|---|
| **Metabolite** | ***n*** | **Mean** | **SD** | **Median** | **IQR** | **Mean** | **SD** | **Median** | **IQR** | **Mean** | **SD** | **Median** | **IQR** |
| ***Medium LDL*** | | | | | | | | | | | | | |
| Particles | 35 | 3.37e-04 | 6.94e-05 | 3.36e-04 | 9.50e-05 | 3.20e-04 | 7.13e-05 | 3.33e-04 | 8.50e-05 | -1.71e-05 | 4.28e-05 | -8.03e-06 | 5.25e-05 |
| Total lipids | 35 | 0.766 | 0.152 | 0.768 | 0.209 | 0.734 | 0.158 | 0.748 | 0.208 | -0.032 | 0.097 | -0.030 | 0.115 |
| Phospholipids | 35 | 0.200 | 0.040 | 0.199 | 0.062 | 0.193 | 0.040 | 0.196 | 0.049 | -0.007 | 0.028 | -0.003 | 0.029 |
| Cholesterol | 35 | 0.533 | 0.110 | 0.536 | 0.144 | 0.506 | 0.115 | 0.521 | 0.152 | -0.027 | 0.068 | -0.028 | 0.084 |
| Cholesteryl esters | 35 | 0.377 | 0.079 | 0.383 | 0.104 | 0.358 | 0.085 | 0.366 | 0.121 | -0.019 | 0.046 | -0.022 | 0.052 |
| Free cholesterol | 35 | 0.156 | 0.033 | 0.156 | 0.045 | 0.147 | 0.032 | 0.149 | 0.030 | -0.009 | 0.024 | -0.003 | 0.028 |
| Triglycerides | 35 | 0.033 | 0.004 | 0.032 | 0.006 | 0.036 | 0.007 | 0.035 | 0.008 | 0.003 | 0.006 | 0.001 | 0.009 |
| ***Small LDL*** | | | | | | | | | | | | | |
| Particles | 35 | 1.93e-04 | 3.31e-05 | 1.96e-04 | 4.60e-05 | 1.83e-04 | 3.42e-05 | 1.83e-04 | 4.50e-05 | -1.06e-05 | 2.29e-05 | -5.24e-06 | 2.20e-05 |
| Total lipids | 35 | 0.338 | 0.060 | 0.341 | 0.079 | 0.322 | 0.060 | 0.330 | 0.069 | -0.016 | 0.040 | -0.008 | 0.042 |
| Phospholipids | 35 | 0.105 | 0.019 | 0.107 | 0.024 | 0.099 | 0.018 | 0.101 | 0.018 | -0.005 | 0.013 | -0.002 | 0.013 |
| Cholesterol | 35 | 0.220 | 0.041 | 0.223 | 0.053 | 0.208 | 0.041 | 0.216 | 0.047 | -0.013 | 0.027 | -0.007 | 0.028 |
| Cholesteryl esters | 35 | 0.155 | 0.028 | 0.154 | 0.037 | 0.147 | 0.029 | 0.152 | 0.038 | -0.008 | 0.018 | -0.005 | 0.016 |
| Free cholesterol | 35 | 0.066 | 0.013 | 0.067 | 0.015 | 0.061 | 0.013 | 0.060 | 0.014 | -0.005 | 0.010 | -0.002 | 0.011 |
| Triglycerides | 35 | 0.013 | 0.002 | 0.013 | 0.002 | 0.015 | 0.004 | 0.014 | 0.004 | 0.001 | 0.003 | 0.001 | 0.003 |
| ***Very large HDL*** | | | | | | | | | | | | | |
| Particles | 35 | 2.91e-04 | 1.07e-04 | 2.72e-04 | 1.14e-04 | 3.06e-04 | 1.02e-04 | 2.66e-04 | 1.15e-04 | 1.48e-05 | 4.54e-05 | 2.75e-05 | 3.45e-05 |
| Total lipids | 35 | 0.208 | 0.090 | 0.191 | 0.092 | 0.219 | 0.087 | 0.190 | 0.095 | 0.011 | 0.039 | 0.019 | 0.039 |
| Phospholipids | 35 | 0.097 | 0.051 | 0.087 | 0.054 | 0.105 | 0.049 | 0.087 | 0.052 | 0.008 | 0.023 | 0.013 | 0.026 |
| Cholesterol | 35 | 0.105 | 0.039 | 0.099 | 0.044 | 0.107 | 0.038 | 0.098 | 0.042 | 0.002 | 0.017 | 0.005 | 0.016 |
| Cholesteryl esters | 35 | 0.079 | 0.031 | 0.076 | 0.035 | 0.082 | 0.030 | 0.074 | 0.035 | 0.002 | 0.013 | 0.004 | 0.011 |
| Free cholesterol | 35 | 0.025 | 0.008 | 0.025 | 0.010 | 0.025 | 0.008 | 0.023 | 0.008 | 0.000 | 0.004 | 0.001 | 0.005 |
| Triglycerides | 35 | 0.006 | 0.001 | 0.006 | 0.002 | 0.007 | 0.002 | 0.007 | 0.003 | 0.001 | 0.002 | 0.001 | 0.002 |
| ***Large HDL*** | | | | | | | | | | | | | |
| Particles | 35 | 2.00e-03 | 7.73e-04 | 1.91e-03 | 8.50e-04 | 2.21e-03 | 7.51e-04 | 1.98e-03 | 9.90e-04 | 2.12e-04 | 3.60e-04 | 2.74e-04 | 5.81e-04 |
| Total lipids | 35 | 0.883 | 0.317 | 0.853 | 0.370 | 0.982 | 0.312 | 0.888 | 0.436 | 0.099 | 0.160 | 0.130 | 0.263 |
| Phospholipids | 35 | 0.425 | 0.146 | 0.406 | 0.168 | 0.479 | 0.145 | 0.433 | 0.209 | 0.055 | 0.085 | 0.057 | 0.131 |
| Cholesterol | 35 | 0.434 | 0.167 | 0.417 | 0.190 | 0.470 | 0.164 | 0.416 | 0.201 | 0.035 | 0.073 | 0.044 | 0.109 |
| Cholesteryl esters | 35 | 0.340 | 0.129 | 0.322 | 0.147 | 0.366 | 0.128 | 0.328 | 0.160 | 0.025 | 0.056 | 0.027 | 0.080 |
| Free cholesterol | 35 | 0.094 | 0.038 | 0.091 | 0.043 | 0.104 | 0.037 | 0.089 | 0.046 | 0.010 | 0.018 | 0.015 | 0.028 |
| Triglycerides | 35 | 0.024 | 0.008 | 0.022 | 0.012 | 0.033 | 0.012 | 0.032 | 0.016 | 0.009 | 0.012 | 0.007 | 0.017 |
| ***Medium HDL*** | | | | | | | | | | | | | |
| Particles | 35 | 4.55e-03 | 6.37e-04 | 4.57e-03 | 7.80e-04 | 5.05e-03 | 7.86e-04 | 4.94e-03 | 1.22e-03 | 4.97e-04 | 7.79e-04 | 4.62e-04 | 8.48e-04 |
| Total lipids | 35 | 1.176 | 0.147 | 1.197 | 0.179 | 1.307 | 0.195 | 1.268 | 0.300 | 0.131 | 0.204 | 0.128 | 0.197 |
| Phospholipids | 35 | 0.532 | 0.061 | 0.542 | 0.091 | 0.596 | 0.090 | 0.583 | 0.139 | 0.065 | 0.099 | 0.060 | 0.087 |
| Cholesterol | 35 | 0.607 | 0.090 | 0.602 | 0.109 | 0.660 | 0.100 | 0.639 | 0.148 | 0.053 | 0.094 | 0.058 | 0.096 |
| Cholesteryl esters | 35 | 0.499 | 0.074 | 0.497 | 0.086 | 0.540 | 0.079 | 0.523 | 0.118 | 0.041 | 0.075 | 0.043 | 0.075 |
| Free cholesterol | 35 | 0.108 | 0.017 | 0.107 | 0.025 | 0.121 | 0.021 | 0.121 | 0.032 | 0.013 | 0.019 | 0.013 | 0.021 |
| Triglycerides | 35 | 0.038 | 0.012 | 0.034 | 0.014 | 0.051 | 0.020 | 0.047 | 0.025 | 0.013 | 0.019 | 0.009 | 0.023 |

| | | First measurement | | | | Last measurement | | | | Change (last -first measurement) | | | |
|---|---|---|---|---|---|---|---|---|---|---|---|---|---|
| **Metabolite** | ***n*** | **Mean** | **SD** | **Median** | **IQR** | **Mean** | **SD** | **Median** | **IQR** | **Mean** | **SD** | **Median** | **IQR** |
| ***Small HDL*** | | | | | | | | | | | | | |
| Particles | 35 | 1.09e-02 | 1.05e-03 | 1.10e-02 | 1.71e-03 | 1.12e-02 | 1.29e-03 | 1.11e-02 | 1.60e-03 | 3.39e-04 | 1.02e-03 | 3.07e-04 | 1.26e-03 |
| Total lipids | 35 | 1.243 | 0.111 | 1.241 | 0.184 | 1.316 | 0.160 | 1.321 | 0.241 | 0.073 | 0.146 | 0.077 | 0.171 |
| Phospholipids | 35 | 0.692 | 0.063 | 0.691 | 0.102 | 0.744 | 0.096 | 0.748 | 0.138 | 0.052 | 0.094 | 0.045 | 0.098 |
| Cholesterol | 35 | 0.510 | 0.046 | 0.518 | 0.077 | 0.524 | 0.057 | 0.523 | 0.067 | 0.014 | 0.047 | 0.016 | 0.058 |
| Cholesteryl esters | 35 | 0.379 | 0.039 | 0.388 | 0.057 | 0.385 | 0.045 | 0.383 | 0.051 | 0.006 | 0.035 | 0.005 | 0.041 |
| Free cholesterol | 35 | 0.131 | 0.009 | 0.129 | 0.014 | 0.139 | 0.015 | 0.138 | 0.020 | 0.007 | 0.013 | 0.006 | 0.012 |
| Triglycerides | 35 | 0.040 | 0.012 | 0.040 | 0.017 | 0.047 | 0.016 | 0.045 | 0.014 | 0.007 | 0.013 | 0.004 | 0.017 |

**Result Supplement Table 3. Menopause-associated metabolite changes.** Skew ($g_1$) and kurtosis ($g_2$) for distribution of residuals for linear mixed model analysis of longitudinal change, lambda parameter used in Box-Cox transformation of metabolite distribution, the estimate of change parameter, $K_{eff}$-Šidák-corrected confidence intervals and *P*-values in crude, covariate-adjusted model (excluding body fat percentage) and full covariate and mediator -adjusted linear mixed model. *P*-values < 0.05 are bolded.

| | | | | **Crude** | | | | **Adjusted** | | | | **Adjusted + body fat %** | | | |
|---|---|---|---|---|---|---|---|---|---|---|---|---|---|---|---|
| | | | | | **99.5% CI** | | | | **99.5% CI** | | | | **99.5% CI** | | |
| **Metabolite** | $g_1$ | $g_2$ | $\lambda$ | **Change** | **Lower** | **Upper** | ***P*** | **Change** | **Lower** | **Upper** | ***P*** | **Change** | **Lower** | **Upper** | ***P*** |
| ***Cholesterols*** | | | | | | | | | | | | | | | |
| Total cholesterol | 0.00 | 0.60 | 0.00 | **0.18** | **0.00** | **0.35** | **0.037** | 0.17 | 0.00 | 0.34 | 0.057 | 0.15 | -0.03 | 0.32 | 0.245 |
| Non-HDL cholesterol | -0.08 | 0.87 | -0.01 | **0.19** | **0.04** | **0.35** | **0.002** | **0.19** | **0.03** | **0.34** | **0.005** | 0.15 | -0.01 | 0.31 | 0.086 |
| Remnant cholesterol | -0.06 | 0.41 | 0.03 | **0.20** | **0.05** | **0.35** | **0.001** | **0.19** | **0.04** | **0.34** | **0.002** | **0.16** | **0.01** | **0.31** | **0.032** |
| VLDL cholesterol | -0.12 | 0.89 | -0.05 | **0.21** | **0.06** | **0.35** | **0.000** | **0.20** | **0.05** | **0.34** | **0.001** | **0.15** | **0.00** | **0.30** | **0.035** |
| Clinical LDL cholesterol | -0.04 | 1.11 | 0.11 | **0.17** | **0.00** | **0.34** | **0.043** | 0.17 | 0.00 | 0.34 | 0.068 | 0.13 | -0.04 | 0.31 | 0.417 |
| LDL cholesterol | -0.08 | 1.11 | -0.04 | **0.18** | **0.01** | **0.34** | **0.020** | **0.17** | **0.01** | **0.34** | **0.034** | **0.14** | -0.03 | 0.30 | 0.319 |
| HDL cholesterol | -0.21 | 0.44 | 0.01 | -0.03 | -0.16 | 0.10 | 1.000 | -0.02 | -0.15 | 0.11 | 1.000 | 0.00 | -0.14 | 0.14 | 1.000 |
| ***Triglycerides*** | | | | | | | | | | | | | | | |
| Total triglycerides | -0.01 | 0.16 | -0.51 | **0.26** | **0.07** | **0.45** | **0.001** | **0.26** | **0.06** | **0.45** | **0.001** | **0.21** | **0.02** | **0.40** | **0.020** |
| VLDL triglycerides | -0.07 | 0.39 | -0.30 | **0.25** | **0.06** | **0.45** | **0.001** | **0.25** | **0.05** | **0.45** | **0.002** | **0.20** | **0.00** | **0.39** | **0.038** |
| LDL triglycerides | -0.06 | 0.53 | -0.94 | 0.16 | -0.01 | 0.33 | 0.089 | 0.15 | -0.02 | 0.32 | 0.140 | 0.12 | -0.05 | 0.29 | 0.553 |
| HDL triglycerides | -0.05 | 1.07 | 0.10 | **0.25** | **0.02** | **0.47** | **0.018** | **0.24** | **0.02** | **0.46** | **0.021** | 0.22 | 0.00 | 0.44 | 0.063 |
| ***Phospholipids*** | | | | | | | | | | | | | | | |
| Total phospholipids | -0.07 | 0.72 | -0.04 | **0.19** | **0.01** | **0.38** | **0.033** | **0.19** | **0.00** | **0.37** | **0.041** | 0.17 | -0.02 | 0.35 | 0.144 |
| VLDL phospholipids | -0.07 | 0.95 | -0.18 | **0.22** | **0.07** | **0.38** | **0.000** | **0.22** | **0.06** | **0.38** | **0.000** | **0.17** | **0.01** | **0.33** | **0.022** |
| LDL phospholipids | -0.05 | 0.75 | -0.08 | **0.17** | **0.00** | **0.33** | **0.041** | 0.16 | 0.00 | 0.32 | 0.060 | 0.13 | -0.04 | 0.29 | 0.433 |
| HDL phospholipids | -0.32 | 0.54 | 0.02 | 0.03 | -0.12 | 0.18 | 1.000 | 0.03 | -0.12 | 0.18 | 1.000 | 0.05 | -0.11 | 0.20 | 1.000 |
| ***Cholesteryl esters*** | | | | | | | | | | | | | | | |
| Total cholesteryl esters | 0.01 | 0.66 | 0.02 | 0.17 | -0.01 | 0.35 | 0.082 | 0.17 | -0.01 | 0.34 | 0.111 | 0.14 | -0.04 | 0.32 | 0.354 |
| VLDL cholesteryl esters | -0.15 | 0.73 | 0.04 | **0.20** | **0.05** | **0.34** | **0.000** | **0.18** | **0.04** | **0.33** | **0.002** | 0.14 | 0.00 | 0.29 | 0.068 |
| LDL cholesteryl esters | -0.10 | 1.09 | -0.05 | **0.19** | **0.03** | **0.35** | **0.008** | **0.18** | **0.02** | **0.34** | **0.013** | 0.14 | -0.02 | 0.31 | 0.199 |
| HDL cholesteryl esters | -0.19 | 0.53 | 0.11 | -0.03 | -0.17 | 0.10 | 1.000 | -0.03 | -0.16 | 0.11 | 1.000 | 0.00 | -0.14 | 0.13 | 1.000 |
| ***Free cholesterol*** | | | | | | | | | | | | | | | |
| Total free cholesterol | -0.02 | 0.54 | -0.07 | **0.19** | **0.03** | **0.35** | **0.005** | **0.18** | **0.02** | **0.34** | **0.011** | 0.15 | -0.01 | 0.32 | 0.099 |
| VLDL free cholesterol | -0.10 | 0.98 | -0.16 | **0.23** | **0.07** | **0.38** | **0.000** | **0.22** | **0.06** | **0.37** | **0.000** | **0.17** | **0.01** | **0.32** | **0.017** |
| LDL free cholesterol | -0.04 | 1.84 | 0.01 | 0.15 | -0.03 | 0.33 | 0.291 | 0.14 | -0.04 | 0.33 | 0.402 | 0.12 | -0.07 | 0.30 | 0.863 |
| HDL free cholesterol | -0.26 | 0.54 | -0.12 | -0.01 | -0.14 | 0.13 | 1.000 | 0.00 | -0.14 | 0.13 | 1.000 | 0.02 | -0.12 | 0.16 | 1.000 |
| ***Total lipids*** | | | | | | | | | | | | | | | |
| Total lipids | -0.01 | 1.06 | -0.53 | **0.22** | **0.05** | **0.40** | **0.002** | **0.22** | **0.04** | **0.39** | **0.003** | **0.18** | **0.00** | **0.35** | **0.042** |
| VLDL total lipids | -0.05 | 0.64 | -0.33 | **0.24** | **0.07** | **0.41** | **0.000** | **0.23** | **0.06** | **0.41** | **0.001** | **0.18** | **0.01** | **0.35** | **0.023** |
| LDL total lipids | -0.06 | 0.86 | -0.08 | **0.18** | **0.02** | **0.34** | **0.015** | **0.17** | **0.01** | **0.33** | **0.025** | 0.14 | -0.03 | 0.30 | 0.272 |
| HDL total lipids | -0.30 | 0.65 | -0.04 | 0.01 | -0.13 | 0.15 | 1.000 | 0.02 | -0.12 | 0.16 | 1.000 | 0.03 | -0.11 | 0.18 | 1.000 |

| | | | | Crude | | | | Adjusted | | | | Adjusted + body fat % | | | |
|---|---|---|---|---|---|---|---|---|---|---|---|---|---|---|---|
| | | | | | 99.95% CI | | | | 99.95% CI | | | | 99.95% CI | | |
| Metabolite | $g_1$ | $g_2$ | $\lambda$ | Change | Lower | Upper | *P* | Change | Lower | Upper | *P* | Change | Lower | Upper | *P* |
| ***Particles*** | | | | | | | | | | | | | | | |
| Total particles | -0.20 | 0.58 | 0.17 | 0.12 | -0.07 | 0.32 | 0.835 | 0.13 | -0.06 | 0.32 | 0.710 | 0.13 | -0.07 | 0.32 | 0.768 |
| VLDL particles | -0.10 | 0.84 | **-0.32** | **0.22** | **0.06** | **0.37** | **0.000** | **0.21** | **0.05** | **0.36** | **0.001** | **0.16** | **0.01** | **0.32** | **0.033** |
| LDL particles | -0.09 | 0.83 | **-0.21** | **0.18** | **0.04** | **0.31** | **0.002** | **0.17** | **0.03** | **0.31** | **0.005** | 0.13 | -0.01 | 0.27 | 0.128 |
| HDL particles | -0.25 | 0.57 | 0.21 | 0.09 | -0.09 | 0.28 | 0.993 | 0.10 | -0.08 | 0.29 | 0.961 | 0.11 | -0.08 | 0.29 | 0.948 |
| ***Particle diameter*** | | | | | | | | | | | | | | | |
| VLDL size | 0.01 | 0.41 | -5.81 | **0.21** | **0.02** | **0.40** | **0.011** | **0.22** | **0.03** | **0.41** | **0.009** | 0.16 | -0.02 | 0.35 | 0.187 |
| LDL size | 0.02 | 0.06 | 6.00 | 0.13 | -0.12 | 0.39 | 0.986 | 0.12 | -0.13 | 0.37 | 0.997 | 0.12 | -0.14 | 0.37 | 0.999 |
| HDL size | -0.07 | -0.02 | -6.00 | -0.10 | -0.22 | 0.01 | 0.166 | -0.11 | -0.23 | 0.01 | 0.129 | -0.07 | -0.20 | 0.05 | 0.920 |
| ***Phosphoglycerides*** | | | | | | | | | | | | | | | |
| Phosphoglycerides | -0.06 | 0.65 | -0.15 | 0.00 | -0.20 | 0.20 | 1.000 | 0.00 | -0.20 | 0.19 | 1.000 | -0.02 | -0.22 | 0.18 | 1.000 |
| Tri-/phosphoglyceride ratio | -0.02 | -0.03 | -0.30 | **0.27** | **0.09** | **0.45** | **0.000** | **0.27** | **0.08** | **0.45** | **0.000** | **0.22** | **0.04** | **0.40** | **0.004** |
| Total cholines | -0.04 | 0.68 | -0.03 | -0.02 | -0.22 | 0.18 | 1.000 | -0.03 | -0.22 | 0.17 | 1.000 | -0.04 | -0.23 | 0.16 | 1.000 |
| Phosphatidylcholines | -0.05 | 0.70 | -0.08 | 0.01 | -0.19 | 0.21 | 1.000 | 0.01 | -0.19 | 0.20 | 1.000 | -0.01 | -0.21 | 0.19 | 1.000 |
| Sphingomyelins | 0.02 | 0.39 | -0.31 | 0.06 | -0.12 | 0.23 | 1.000 | 0.05 | -0.13 | 0.22 | 1.000 | 0.03 | -0.15 | 0.21 | 1.000 |
| ***Apolipoproteins*** | | | | | | | | | | | | | | | |
| Apolipoprotein B | -0.07 | 0.79 | -0.18 | **0.18** | **0.05** | **0.32** | **0.001** | **0.17** | **0.03** | **0.31** | **0.002** | 0.14 | 0.00 | 0.28 | 0.069 |
| Apolipoprotein A-I | -0.32 | 0.67 | 0.00 | 0.05 | -0.11 | 0.21 | 1.000 | 0.05 | -0.10 | 0.21 | 1.000 | 0.07 | -0.09 | 0.23 | 1.000 |
| Apo B/A-I ratio | -0.12 | 1.15 | -0.16 | **0.12** | **0.01** | **0.23** | **0.023** | 0.11 | 0.00 | 0.22 | 0.066 | 0.08 | -0.04 | 0.20 | 0.743 |
| ***Fatty acids*** | | | | | | | | | | | | | | | |
| Total fatty acids | 0.04 | 0.70 | -1.37 | 0.06 | -0.13 | 0.25 | 1.000 | 0.05 | -0.13 | 0.24 | 1.000 | 0.01 | -0.18 | 0.19 | 1.000 |
| Degree of unsaturation | 0.05 | 0.72 | -0.23 | -0.15 | -0.34 | 0.05 | 0.500 | -0.15 | -0.35 | 0.05 | 0.484 | -0.13 | -0.33 | 0.07 | 0.750 |
| Omega-3 fatty acids | 0.15 | -0.17 | 0.09 | -0.10 | -0.28 | 0.08 | 0.974 | -0.09 | -0.28 | 0.09 | 0.994 | -0.12 | -0.31 | 0.07 | 0.814 |
| Omega-6 fatty acids | 0.11 | 1.56 | -0.71 | -0.05 | -0.26 | 0.15 | 1.000 | -0.07 | -0.27 | 0.13 | 1.000 | -0.10 | -0.30 | 0.10 | 0.997 |
| Polyunsaturated fatty acids | 0.08 | 0.99 | -0.73 | -0.07 | -0.26 | 0.12 | 1.000 | -0.08 | -0.27 | 0.11 | 1.000 | -0.11 | -0.30 | 0.08 | 0.918 |
| Monounsaturated fatty acids | 0.02 | 0.68 | -1.06 | 0.06 | -0.14 | 0.25 | 1.000 | 0.05 | -0.14 | 0.24 | 1.000 | 0.00 | -0.19 | 0.19 | 1.000 |
| Saturated fatty acids | -0.03 | 0.98 | -1.29 | 0.20 | 0.00 | 0.39 | 0.056 | 0.19 | 0.00 | 0.38 | 0.066 | 0.14 | -0.05 | 0.34 | 0.506 |
| Linoleic acid | 0.09 | 0.72 | -0.27 | **-0.29** | **-0.53** | **-0.05** | **0.003** | **-0.32** | **-0.55** | **-0.08** | **0.001** | **-0.34** | **-0.57** | **-0.10** | **0.000** |
| Docosahexaenoic acid | 0.08 | 0.09 | -0.41 | **-0.21** | **-0.40** | **-0.02** | **0.012** | **-0.21** | **-0.40** | **-0.01** | **0.023** | **-0.22** | **-0.41** | **-0.02** | **0.014** |
| ***Fatty acid ratios (%)*** | | | | | | | | | | | | | | | |
| Omega-3 ratio | 0.31 | 0.40 | 0.25 | -0.16 | -0.36 | 0.04 | 0.374 | -0.15 | -0.35 | 0.06 | 0.573 | -0.16 | -0.37 | 0.05 | 0.483 |
| Omega-6 ratio | 0.18 | 0.44 | 5.58 | **-0.27** | **-0.48** | **-0.06** | **0.002** | **-0.27** | **-0.48** | **-0.06** | **0.002** | **-0.22** | **-0.44** | **-0.01** | **0.033** |
| Polyunsaturated ratio | 0.17 | 0.89 | 4.96 | **-0.31** | **-0.52** | **-0.09** | **0.000** | **-0.31** | **-0.52** | **-0.09** | **0.000** | **-0.26** | **-0.48** | **-0.05** | **0.003** |
| Monounsaturated ratio | 0.04 | 1.36 | -0.34 | 0.04 | -0.16 | 0.23 | 1.000 | 0.04 | -0.16 | 0.23 | 1.000 | -0.01 | -0.20 | 0.19 | 1.000 |
| Saturated ratio | -0.22 | 1.79 | -0.76 | **0.43** | **0.16** | **0.70** | **0.000** | **0.43** | **0.16** | **0.70** | **0.000** | **0.42** | **0.15** | **0.69** | **0.000** |
| Linoleic acid ratio | 0.18 | 0.09 | 2.98 | **-0.57** | **-0.81** | **-0.32** | **0.000** | **-0.58** | **-0.83** | **-0.33** | **0.000** | **-0.54** | **-0.79** | **-0.29** | **0.000** |
| Docosahexaenoic acid ratio | 0.10 | 0.08 | -0.04 | **-0.27** | **-0.47** | **-0.08** | **0.000** | **-0.27** | **-0.47** | **-0.07** | **0.001** | **-0.25** | **-0.45** | **-0.05** | **0.002** |
| Polyu/monounsaturated ratio | -0.01 | 1.32 | 1.17 | -0.15 | -0.35 | 0.04 | 0.414 | -0.15 | -0.35 | 0.05 | 0.437 | -0.11 | -0.30 | 0.09 | 0.983 |
| Omega-6 to omega-3 ratio | -0.25 | 0.26 | -0.21 | 0.09 | -0.11 | 0.29 | 0.999 | 0.08 | -0.12 | 0.28 | 1.000 | 0.10 | -0.11 | 0.31 | 0.997 |

| | | | | Crude | | | | Adjusted | | | | Adjusted + body fat % | | | |
|---|---|---|---|---|---|---|---|---|---|---|---|---|---|---|---|
| | | | | | 99.95% CI | | | | 99.95% CI | | | | 99.95% CI | | |
| **Metabolite** | $g_1$ | $g_2$ | $\lambda$ | **Change** | **Lower** | **Upper** | ***P*** | **Change** | **Lower** | **Upper** | ***P*** | **Change** | **Lower** | **Upper** | ***P*** |
| ***Amino acids*** | | | | | | | | | | | | | | | |
| Alanine | 0.04 | 0.00 | -0.16 | 0.13 | -0.11 | 0.38 | 0.976 | 0.13 | -0.11 | 0.38 | 0.981 | 0.09 | -0.16 | 0.33 | 1.000 |
| Glutamine | 0.07 | -0.19 | 0.76 | **-0.44** | **-0.70** | **-0.19** | **0.000** | **-0.46** | **-0.72** | **-0.20** | **0.000** | **-0.45** | **-0.71** | **-0.19** | **0.000** |
| Glycine | 0.05 | 0.05 | -0.55 | 0.12 | -0.01 | 0.25 | 0.129 | 0.10 | -0.03 | 0.23 | 0.459 | 0.11 | -0.03 | 0.25 | 0.329 |
| Histidine | 0.06 | 0.65 | 0.52 | 0.07 | -0.17 | 0.30 | 1.000 | 0.07 | -0.16 | 0.31 | 1.000 | 0.07 | -0.17 | 0.30 | 1.000 |
| Total BCAA | -0.14 | -0.20 | -0.16 | 0.22 | -0.02 | 0.45 | 0.122 | 0.22 | -0.03 | 0.46 | 0.152 | 0.18 | -0.06 | 0.42 | 0.527 |
| Isoleucine | -0.15 | 1.04 | 0.45 | 0.19 | -0.06 | 0.44 | 0.516 | 0.18 | -0.08 | 0.43 | 0.652 | 0.15 | -0.11 | 0.40 | 0.950 |
| Leucine | -0.14 | -0.27 | 0.15 | **0.25** | **0.02** | **0.48** | **0.023** | **0.25** | **0.02** | **0.49** | **0.023** | 0.22 | -0.01 | 0.46 | 0.088 |
| Valine | -0.03 | -0.20 | -0.08 | 0.19 | -0.05 | 0.43 | 0.364 | 0.19 | -0.06 | 0.43 | 0.445 | 0.15 | -0.10 | 0.40 | 0.913 |
| Phenylalanine | -0.04 | -0.36 | 0.00 | 0.12 | -0.13 | 0.37 | 0.997 | 0.12 | -0.13 | 0.38 | 0.996 | 0.08 | -0.17 | 0.33 | 1.000 |
| Tyrosine | -0.04 | 0.41 | 0.03 | 0.20 | -0.04 | 0.43 | 0.236 | 0.21 | -0.02 | 0.44 | 0.120 | 0.16 | -0.07 | 0.40 | 0.636 |
| ***Glycolysis-related*** | | | | | | | | | | | | | | | |
| Glucose | -0.43 | 0.53 | -1.26 | -0.10 | -0.32 | 0.11 | 0.998 | -0.09 | -0.31 | 0.12 | 1.000 | -0.13 | -0.34 | 0.09 | 0.936 |
| Lactate | 0.11 | 1.26 | -0.71 | -0.03 | -0.31 | 0.25 | 1.000 | -0.03 | -0.31 | 0.25 | 1.000 | -0.07 | -0.35 | 0.21 | 1.000 |
| Pyruvate | 0.10 | 0.54 | -0.10 | 0.06 | -0.20 | 0.32 | 1.000 | 0.05 | -0.21 | 0.32 | 1.000 | 0.03 | -0.24 | 0.29 | 1.000 |
| Citrate | -0.25 | 0.82 | -0.96 | **-0.33** | **-0.54** | **-0.12** | **0.000** | **-0.36** | **-0.57** | **-0.14** | **0.000** | **-0.38** | **-0.60** | **-0.16** | **0.000** |
| Glycerol | -0.10 | 1.25 | -0.14 | **0.33** | **0.07** | **0.58** | **0.001** | **0.32** | **0.07** | **0.58** | **0.002** | **0.28** | **0.02** | **0.54** | **0.025** |
| ***Ketone bodies*** | | | | | | | | | | | | | | | |
| 3-Hydroxybutyrate | -0.10 | 0.05 | 0.13 | **-0.47** | **-0.75** | **-0.18** | **0.000** | **-0.46** | **-0.75** | **-0.17** | **0.000** | **-0.45** | **-0.74** | **-0.16** | **0.000** |
| Acetate | -0.01 | -0.01 | -0.36 | 0.14 | -0.15 | 0.42 | 0.997 | 0.16 | -0.13 | 0.44 | 0.978 | 0.18 | -0.11 | 0.46 | 0.880 |
| Acetoacetate | -0.06 | -0.24 | -0.36 | **-0.44** | **-0.73** | **-0.16** | **0.000** | **-0.44** | **-0.72** | **-0.15** | **0.000** | **-0.44** | **-0.73** | **-0.15** | **0.000** |
| Acetone | -0.04 | 0.22 | -1.21 | -0.28 | -0.58 | 0.02 | 0.093 | -0.27 | -0.57 | 0.02 | 0.115 | -0.25 | -0.55 | 0.05 | 0.230 |
| ***Miscellaneous*** | | | | | | | | | | | | | | | |
| Creatinine | 0.03 | 0.66 | -0.06 | 0.06 | -0.10 | 0.22 | 1.000 | 0.06 | -0.10 | 0.22 | 1.000 | 0.05 | -0.11 | 0.22 | 1.000 |
| Albumin | 0.15 | 0.28 | 2.06 | 0.07 | -0.15 | 0.29 | 1.000 | 0.08 | -0.15 | 0.31 | 1.000 | 0.10 | -0.13 | 0.33 | 0.999 |
| Glycoprotein acetyls | 0.20 | 0.38 | -0.37 | -0.07 | -0.26 | 0.12 | 1.000 | -0.09 | -0.28 | 0.10 | 0.999 | -0.15 | -0.34 | 0.04 | 0.348 |
| ***Chylomicrons and extremely large VLDL*** | | | | | | | | | | | | | | | |
| Particles | 0.03 | -0.19 | -0.13 | **0.26** | **0.05** | **0.48** | **0.002** | **0.26** | **0.05** | **0.48** | **0.003** | 0.21 | 0.00 | 0.42 | 0.056 |
| Total lipids | -0.05 | -0.19 | -0.12 | 0.17 | -0.04 | 0.39 | 0.338 | 0.17 | -0.04 | 0.39 | 0.334 | 0.12 | -0.09 | 0.33 | 0.956 |
| Phospholipids | -0.10 | -0.02 | -0.10 | **0.26** | **0.04** | **0.49** | **0.007** | **0.26** | **0.03** | **0.49** | **0.009** | 0.21 | -0.02 | 0.43 | 0.106 |
| Cholesterol | 0.01 | 0.11 | 0.05 | 0.13 | -0.06 | 0.33 | 0.699 | 0.13 | -0.07 | 0.32 | 0.774 | 0.08 | -0.11 | 0.27 | 1.000 |
| Cholesteryl esters | 0.02 | 0.55 | 0.21 | 0.09 | -0.11 | 0.29 | 0.999 | 0.08 | -0.11 | 0.28 | 1.000 | 0.03 | -0.16 | 0.23 | 1.000 |
| Free cholesterol | -0.21 | 0.09 | -0.15 | **0.29** | **0.06** | **0.52** | **0.002** | **0.29** | **0.06** | **0.52** | **0.003** | **0.24** | **0.01** | **0.47** | **0.034** |
| Triglycerides | -0.30 | -0.47 | 0.09 | 0.25 | -0.01 | 0.50 | 0.068 | 0.26 | 0.00 | 0.51 | 0.047 | 0.21 | -0.04 | 0.46 | 0.290 |
| ***Very large VLDL*** | | | | | | | | | | | | | | | |
| Particles | 0.03 | 0.98 | 0.34 | **0.25** | **0.07** | **0.43** | **0.000** | **0.25** | **0.06** | **0.43** | **0.001** | **0.19** | **0.01** | **0.38** | **0.021** |
| Total lipids | 0.00 | 0.84 | 0.26 | **0.24** | **0.06** | **0.43** | **0.001** | **0.24** | **0.05** | **0.43** | **0.001** | **0.19** | **0.00** | **0.37** | **0.039** |
| Phospholipids | -0.04 | 1.15 | 0.40 | **0.25** | **0.08** | **0.43** | **0.000** | **0.25** | **0.07** | **0.43** | **0.000** | **0.20** | **0.02** | **0.38** | **0.014** |
| Cholesterol | -0.01 | 1.06 | 0.28 | **0.20** | **0.04** | **0.36** | **0.003** | **0.19** | **0.03** | **0.36** | **0.005** | 0.14 | -0.02 | 0.30 | 0.180 |
| Cholesteryl esters | -0.05 | 1.13 | 0.32 | **0.16** | **0.01** | **0.32** | **0.023** | 0.16 | 0.00 | 0.31 | 0.051 | 0.11 | -0.05 | 0.26 | 0.711 |

| | | | | Crude | | | | Adjusted | | | | Adjusted + body fat % | | | |
|---|---|---|---|---|---|---|---|---|---|---|---|---|---|---|---|
| | | | | | 99.95% CI | | | | 99.95% CI | | | | 99.95% CI | | |
| **Metabolite** | $g_1$ | $g_2$ | $\lambda$ | **Change** | **Lower** | **Upper** | $P$ | **Change** | **Lower** | **Upper** | $P$ | **Change** | **Lower** | **Upper** | $P$ |
| ***Very large VLDL continue*** | | | | | | | | | | | | | | | |
| Free cholesterol | 0.01 | 1.05 | 0.34 | **0.25** | **0.07** | **0.42** | **0.000** | **0.24** | **0.07** | **0.42** | **0.000** | **0.19** | **0.02** | **0.37** | **0.017** |
| Triglycerides | -0.13 | 1.70 | 0.35 | **0.25** | **0.05** | **0.46** | **0.002** | **0.25** | **0.05** | **0.46** | **0.003** | 0.20 | 0.00 | 0.40 | 0.058 |
| ***Large VLDL*** | | | | | | | | | | | | | | | |
| Particles | -0.03 | 0.44 | 0.09 | **0.23** | **0.05** | **0.42** | **0.001** | **0.23** | **0.05** | **0.41** | **0.002** | 0.18 | 0.00 | 0.36 | 0.052 |
| Total lipids | -0.11 | 0.85 | 0.12 | **0.24** | **0.05** | **0.42** | **0.001** | **0.23** | **0.05** | **0.42** | **0.003** | 0.18 | 0.00 | 0.36 | 0.065 |
| Phospholipids | 0.00 | 1.38 | 0.43 | **0.23** | **0.05** | **0.41** | **0.002** | **0.22** | **0.04** | **0.41** | **0.002** | 0.18 | 0.00 | 0.35 | 0.060 |
| Cholesterol | -0.05 | 0.90 | 0.31 | **0.23** | **0.06** | **0.39** | **0.000** | **0.22** | **0.05** | **0.39** | **0.001** | **0.17** | **0.01** | **0.34** | **0.031** |
| Cholesteryl esters | -0.07 | 0.89 | 0.34 | **0.22** | **0.06** | **0.38** | **0.000** | **0.21** | **0.05** | **0.38** | **0.001** | **0.17** | **0.00** | **0.33** | **0.041** |
| Free cholesterol | -0.02 | 0.92 | 0.31 | **0.24** | **0.07** | **0.41** | **0.000** | **0.23** | **0.06** | **0.41** | **0.001** | **0.18** | **0.01** | **0.35** | **0.026** |
| Triglycerides | -0.12 | 0.94 | -0.01 | **0.26** | **0.06** | **0.46** | **0.001** | **0.26** | **0.06** | **0.47** | **0.001** | **0.21** | **0.01** | **0.40** | **0.029** |
| ***Medium VLDL*** | | | | | | | | | | | | | | | |
| Particles | -0.13 | 0.92 | -0.10 | **0.21** | **0.06** | **0.36** | **0.000** | **0.20** | **0.05** | **0.36** | **0.001** | **0.15** | **0.00** | **0.31** | **0.049** |
| Total lipids | -0.12 | 0.74 | -0.18 | **0.22** | **0.06** | **0.39** | **0.001** | **0.21** | **0.05** | **0.38** | **0.002** | 0.16 | 0.00 | 0.33 | 0.058 |
| Phospholipids | -0.13 | 0.94 | -0.01 | **0.21** | **0.06** | **0.36** | **0.000** | **0.20** | **0.04** | **0.35** | **0.001** | 0.15 | 0.00 | 0.30 | 0.055 |
| Cholesterol | -0.13 | 0.50 | 0.22 | **0.19** | **0.04** | **0.33** | **0.001** | **0.18** | **0.03** | **0.32** | **0.004** | 0.14 | -0.01 | 0.29 | 0.082 |
| Cholesteryl esters | -0.07 | 0.28 | 0.33 | **0.17** | **0.01** | **0.33** | **0.023** | **0.16** | **0.00** | **0.32** | **0.061** | 0.13 | -0.03 | 0.29 | 0.348 |
| Free cholesterol | -0.14 | 0.92 | 0.06 | **0.20** | **0.06** | **0.35** | **0.000** | **0.19** | **0.04** | **0.34** | **0.001** | **0.15** | **0.00** | **0.30** | **0.048** |
| Triglycerides | -0.13 | 0.69 | -0.21 | **0.23** | **0.04** | **0.42** | **0.005** | **0.22** | **0.03** | **0.42** | **0.009** | 0.17 | -0.02 | 0.36 | 0.143 |
| ***Small VLDL*** | | | | | | | | | | | | | | | |
| Particles | -0.08 | 0.47 | -0.22 | **0.21** | **0.05** | **0.37** | **0.002** | **0.20** | **0.03** | **0.36** | **0.004** | 0.16 | -0.01 | 0.32 | 0.080 |
| Total lipids | -0.08 | 0.55 | -0.20 | **0.21** | **0.05** | **0.37** | **0.001** | **0.20** | **0.04** | **0.36** | **0.002** | 0.16 | 0.00 | 0.32 | 0.054 |
| Phospholipids | -0.13 | 0.68 | -0.08 | **0.19** | **0.04** | **0.33** | **0.001** | **0.18** | **0.03** | **0.32** | **0.004** | 0.14 | -0.01 | 0.28 | 0.101 |
| Cholesterol | -0.18 | 0.72 | 0.07 | **0.19** | **0.05** | **0.33** | **0.001** | **0.18** | **0.03** | **0.32** | **0.003** | 0.14 | 0.00 | 0.28 | 0.073 |
| Cholesteryl esters | -0.20 | 0.82 | 0.14 | **0.19** | **0.05** | **0.33** | **0.001** | **0.18** | **0.03** | **0.32** | **0.003** | 0.14 | 0.00 | 0.29 | 0.066 |
| Free cholesterol | -0.15 | 0.57 | 0.01 | **0.19** | **0.04** | **0.33** | **0.001** | **0.17** | **0.03** | **0.32** | **0.004** | 0.13 | -0.01 | 0.28 | 0.106 |
| Triglycerides | 0.02 | 0.07 | -0.21 | **0.23** | **0.03** | **0.43** | **0.009** | **0.23** | **0.03** | **0.43** | **0.011** | 0.19 | -0.01 | 0.39 | 0.092 |
| ***Very small VLDL*** | | | | | | | | | | | | | | | |
| Particles | -0.04 | 0.34 | -0.21 | **0.18** | **0.03** | **0.33** | **0.005** | **0.16** | **0.01** | **0.31** | **0.021** | 0.14 | -0.02 | 0.29 | 0.140 |
| Total lipids | -0.10 | 0.58 | -0.22 | **0.20** | **0.04** | **0.35** | **0.002** | **0.18** | **0.02** | **0.33** | **0.011** | 0.15 | -0.01 | 0.31 | 0.086 |
| Phospholipids | -0.16 | 0.75 | -0.08 | **0.19** | **0.03** | **0.35** | **0.008** | **0.17** | **0.01** | **0.33** | **0.035** | 0.14 | -0.02 | 0.31 | 0.205 |
| Cholesterol | 0.01 | 0.13 | 0.17 | **0.19** | **0.04** | **0.34** | **0.003** | **0.17** | **0.02** | **0.32** | **0.014** | 0.15 | -0.01 | 0.30 | 0.081 |
| Cholesteryl esters | 0.04 | 0.21 | 0.28 | **0.18** | **0.03** | **0.34** | **0.007** | **0.17** | **0.01** | **0.33** | **0.027** | 0.15 | -0.01 | 0.31 | 0.111 |
| Free cholesterol | -0.08 | 0.40 | -0.07 | **0.19** | **0.04** | **0.34** | **0.002** | **0.17** | **0.02** | **0.32** | **0.011** | 0.14 | -0.01 | 0.30 | 0.095 |
| Triglycerides | -0.07 | 0.71 | -0.19 | **0.20** | **0.00** | **0.39** | **0.041** | 0.19 | 0.00 | 0.38 | 0.062 | 0.16 | -0.03 | 0.35 | 0.246 |
| ***IDL*** | | | | | | | | | | | | | | | |
| Particles | 0.11 | 0.25 | 0.27 | 0.16 | 0.00 | 0.33 | 0.062 | 0.16 | -0.01 | 0.32 | 0.106 | 0.14 | -0.03 | 0.31 | 0.351 |
| Total lipids | 0.07 | 0.40 | 0.24 | 0.17 | 0.00 | 0.34 | 0.050 | 0.16 | -0.01 | 0.33 | 0.093 | 0.14 | -0.03 | 0.31 | 0.277 |
| Phospholipids | -0.01 | 0.19 | 0.08 | **0.17** | **0.01** | **0.34** | **0.023** | 0.16 | 0.00 | 0.32 | 0.063 | 0.14 | -0.03 | 0.30 | 0.279 |
| Cholesterol | 0.08 | 1.45 | 0.33 | 0.16 | -0.02 | 0.34 | 0.164 | 0.15 | -0.03 | 0.33 | 0.245 | 0.14 | -0.05 | 0.32 | 0.482 |
| Cholesteryl esters | 0.09 | 1.35 | 0.37 | 0.16 | -0.03 | 0.34 | 0.203 | 0.15 | -0.03 | 0.34 | 0.283 | 0.14 | -0.05 | 0.33 | 0.523 |
| Free cholesterol | 0.09 | 1.51 | 0.24 | 0.16 | -0.01 | 0.33 | 0.121 | 0.15 | -0.03 | 0.32 | 0.221 | 0.13 | -0.04 | 0.31 | 0.472 |

| Metabolite | $g_1$ | $g_2$ | $\lambda$ | Crude Change | Crude 99.95% CI Lower | Crude 99.95% CI Upper | Crude *P* | Adjusted Change | Adjusted 99.95% CI Lower | Adjusted 99.95% CI Upper | Adjusted *P* | Adjusted + body fat % Change | Adjusted + body fat % 99.95% CI Lower | Adjusted + body fat % 99.95% CI Upper | Adjusted + body fat % *P* |
|---|---|---|---|---|---|---|---|---|---|---|---|---|---|---|---|
| ***IDL continue*** | | | | | | | | | | | | | | | |
| Triglycerides | -0.11 | 0.50 | -0.74 | 0.16 | -0.03 | 0.34 | 0.229 | 0.14 | -0.04 | 0.32 | 0.374 | 0.12 | -0.06 | 0.31 | 0.757 |
| ***Large LDL*** | | | | | | | | | | | | | | | |
| Particles | -0.07 | 0.49 | -0.28 | **0.19** | **0.04** | **0.33** | **0.001** | **0.18** | **0.03** | **0.32** | **0.004** | 0.14 | -0.01 | 0.29 | 0.097 |
| Total lipids | -0.06 | 0.75 | -0.04 | **0.18** | **0.01** | **0.35** | **0.023** | **0.17** | **0.00** | **0.34** | **0.043** | 0.14 | -0.03 | 0.31 | 0.303 |
| Phospholipids | -0.05 | 0.69 | -0.01 | 0.17 | 0.00 | 0.34 | 0.059 | 0.16 | -0.01 | 0.34 | 0.089 | 0.13 | -0.04 | 0.31 | 0.455 |
| Cholesterol | -0.07 | 1.05 | -0.02 | **0.18** | **0.01** | **0.36** | **0.031** | 0.17 | 0.00 | 0.35 | 0.056 | 0.14 | -0.04 | 0.32 | 0.353 |
| Cholesteryl esters | -0.09 | 0.92 | -0.02 | **0.19** | **0.02** | **0.36** | **0.017** | **0.18** | **0.01** | **0.35** | **0.031** | 0.14 | -0.03 | 0.32 | 0.267 |
| Free cholesterol | -0.05 | 1.88 | 0.03 | 0.16 | -0.03 | 0.35 | 0.215 | 0.15 | -0.04 | 0.34 | 0.325 | 0.13 | -0.06 | 0.32 | 0.746 |
| Triglycerides | -0.08 | 0.39 | -0.83 | 0.14 | -0.03 | 0.31 | 0.330 | 0.13 | -0.04 | 0.29 | 0.504 | 0.10 | -0.07 | 0.28 | 0.899 |
| ***Medium LDL*** | | | | | | | | | | | | | | | |
| Particles | -0.12 | 0.94 | -0.09 | **0.15** | **0.01** | **0.29** | **0.023** | **0.15** | **0.00** | **0.29** | **0.036** | 0.11 | -0.04 | 0.26 | 0.481 |
| Total lipids | -0.10 | 1.00 | -0.07 | **0.17** | **0.02** | **0.32** | **0.014** | **0.17** | **0.01** | **0.32** | **0.019** | 0.13 | -0.03 | 0.28 | 0.318 |
| Phospholipids | -0.09 | 0.78 | -0.08 | **0.16** | **0.01** | **0.32** | **0.034** | **0.16** | **0.00** | **0.32** | **0.048** | 0.12 | -0.04 | 0.28 | 0.465 |
| Cholesterol | -0.11 | 1.17 | -0.01 | **0.17** | **0.01** | **0.32** | **0.018** | **0.17** | **0.01** | **0.32** | **0.023** | 0.12 | -0.03 | 0.28 | 0.361 |
| Cholesteryl esters | -0.13 | 1.16 | -0.05 | **0.17** | **0.02** | **0.32** | **0.008** | **0.17** | **0.02** | **0.32** | **0.009** | 0.13 | -0.02 | 0.28 | 0.243 |
| Free cholesterol | -0.06 | 1.39 | 0.07 | 0.14 | -0.04 | 0.32 | 0.405 | 0.14 | -0.05 | 0.32 | 0.497 | 0.11 | -0.08 | 0.29 | 0.950 |
| Triglycerides | -0.03 | 0.56 | -0.96 | **0.17** | **0.01** | **0.34** | **0.029** | **0.16** | **0.00** | **0.33** | **0.042** | 0.13 | -0.03 | 0.30 | 0.294 |
| ***Small LDL*** | | | | | | | | | | | | | | | |
| Particles | -0.04 | 0.82 | -0.13 | 0.14 | -0.02 | 0.29 | 0.154 | 0.13 | -0.02 | 0.29 | 0.215 | 0.10 | -0.06 | 0.25 | 0.877 |
| Total lipids | -0.02 | 1.06 | -0.18 | **0.15** | **0.01** | **0.29** | **0.030** | **0.15** | **0.00** | **0.29** | **0.043** | 0.11 | -0.04 | 0.26 | 0.500 |
| Phospholipids | 0.01 | 0.96 | -0.03 | 0.13 | -0.01 | 0.27 | 0.105 | 0.13 | -0.01 | 0.27 | 0.141 | 0.10 | -0.05 | 0.24 | 0.755 |
| Cholesterol | -0.05 | 1.04 | -0.12 | 0.15 | 0.00 | 0.30 | 0.066 | 0.15 | -0.01 | 0.30 | 0.090 | 0.11 | -0.05 | 0.26 | 0.688 |
| Cholesteryl esters | -0.06 | 1.05 | -0.23 | **0.17** | **0.02** | **0.31** | **0.014** | **0.16** | **0.01** | **0.31** | **0.021** | 0.12 | -0.03 | 0.27 | 0.364 |
| Free cholesterol | -0.04 | 1.08 | 0.22 | 0.08 | -0.09 | 0.25 | 0.997 | 0.08 | -0.09 | 0.25 | 0.999 | 0.05 | -0.13 | 0.22 | 1.000 |
| Triglycerides | -0.01 | 0.62 | -1.02 | **0.24** | **0.06** | **0.42** | **0.001** | **0.24** | **0.06** | **0.42** | **0.001** | **0.20** | **0.02** | **0.37** | **0.017** |
| ***Very large HDL*** | | | | | | | | | | | | | | | |
| Particles | -0.27 | 1.26 | 0.13 | -0.06 | -0.19 | 0.07 | 1.000 | -0.07 | -0.20 | 0.06 | 0.986 | -0.04 | -0.18 | 0.10 | 1.000 |
| Total lipids | -0.29 | 1.49 | 0.34 | -0.08 | -0.21 | 0.05 | 0.913 | -0.09 | -0.23 | 0.04 | 0.745 | -0.06 | -0.19 | 0.08 | 1.000 |
| Phospholipids | -0.23 | 0.73 | 0.49 | -0.09 | -0.22 | 0.03 | 0.534 | -0.10 | -0.23 | 0.03 | 0.374 | -0.07 | -0.20 | 0.07 | 0.995 |
| Cholesterol | -0.25 | 1.00 | 0.23 | -0.08 | -0.22 | 0.05 | 0.877 | -0.09 | -0.23 | 0.04 | 0.714 | -0.06 | -0.20 | 0.08 | 1.000 |
| Cholesteryl esters | -0.23 | 0.89 | 0.24 | -0.08 | -0.21 | 0.05 | 0.806 | -0.09 | -0.22 | 0.04 | 0.640 | -0.06 | -0.19 | 0.08 | 1.000 |
| Free cholesterol | -0.33 | 1.21 | 0.25 | -0.08 | -0.23 | 0.08 | 0.995 | -0.09 | -0.24 | 0.07 | 0.949 | -0.05 | -0.21 | 0.10 | 1.000 |
| Triglycerides | 0.07 | 0.65 | -0.35 | 0.16 | -0.04 | 0.36 | 0.398 | 0.14 | -0.06 | 0.34 | 0.592 | 0.14 | -0.06 | 0.34 | 0.665 |
| ***Large HDL*** | | | | | | | | | | | | | | | |
| Particles | -0.16 | 0.50 | 0.30 | -0.07 | -0.18 | 0.05 | 0.943 | -0.07 | -0.18 | 0.05 | 0.924 | -0.04 | -0.16 | 0.08 | 1.000 |
| Total lipids | -0.14 | 0.35 | 0.33 | -0.07 | -0.18 | 0.04 | 0.895 | -0.07 | -0.19 | 0.04 | 0.884 | -0.04 | -0.16 | 0.08 | 1.000 |
| Phospholipids | -0.14 | 0.35 | 0.35 | -0.07 | -0.18 | 0.05 | 0.920 | -0.07 | -0.19 | 0.05 | 0.926 | -0.04 | -0.16 | 0.08 | 1.000 |
| Cholesterol | -0.15 | 0.24 | 0.37 | -0.08 | -0.19 | 0.04 | 0.717 | -0.08 | -0.20 | 0.04 | 0.704 | -0.05 | -0.17 | 0.07 | 1.000 |
| Cholesteryl esters | -0.14 | 0.17 | 0.39 | -0.08 | -0.20 | 0.03 | 0.645 | -0.08 | -0.20 | 0.03 | 0.640 | -0.05 | -0.17 | 0.07 | 1.000 |
| Free cholesterol | -0.19 | 0.71 | 0.39 | -0.07 | -0.19 | 0.05 | 0.947 | -0.07 | -0.19 | 0.05 | 0.928 | -0.04 | -0.17 | 0.08 | 1.000 |
| Triglycerides | -0.20 | 0.67 | -0.02 | 0.16 | -0.05 | 0.37 | 0.458 | 0.15 | -0.06 | 0.36 | 0.648 | 0.16 | -0.05 | 0.38 | 0.480 |

| | | | | Crude | | | | Adjusted | | | | Adjusted + body fat % | | | |
|---|---|---|---|---|---|---|---|---|---|---|---|---|---|---|---|
| | | | | | 99.95% CI | | | | 99.95% CI | | | | 99.95% CI | | |
| **Metabolite** | $g_1$ | $g_2$ | $\lambda$ | **Change** | **Lower** | **Upper** | ***P*** | **Change** | **Lower** | **Upper** | ***P*** | **Change** | **Lower** | **Upper** | ***P*** |
| ***Medium HDL*** | | | | | | | | | | | | | | | |
| Particles | -0.29 | 0.50 | 0.40 | 0.04 | -0.12 | 0.20 | 1.000 | 0.05 | -0.10 | 0.21 | 1.000 | 0.06 | -0.10 | 0.22 | 1.000 |
| Total lipids | -0.29 | 0.41 | 0.39 | 0.06 | -0.11 | 0.22 | 1.000 | 0.06 | -0.10 | 0.23 | 1.000 | 0.08 | -0.09 | 0.24 | 0.999 |
| Phospholipids | -0.32 | 0.43 | 0.32 | 0.08 | -0.10 | 0.26 | 0.999 | 0.09 | -0.09 | 0.27 | 0.991 | 0.10 | -0.08 | 0.28 | 0.975 |
| Cholesterol | -0.20 | 0.41 | 0.47 | 0.00 | -0.15 | 0.15 | 1.000 | 0.01 | -0.13 | 0.16 | 1.000 | 0.03 | -0.12 | 0.18 | 1.000 |
| Cholesteryl esters | -0.17 | 0.45 | 0.53 | 0.00 | -0.15 | 0.14 | 1.000 | 0.01 | -0.14 | 0.15 | 1.000 | 0.02 | -0.13 | 0.17 | 1.000 |
| Free cholesterol | -0.31 | 0.60 | 0.33 | 0.03 | -0.13 | 0.19 | 1.000 | 0.04 | -0.12 | 0.19 | 1.000 | 0.05 | -0.11 | 0.21 | 1.000 |
| Triglycerides | 0.06 | 1.63 | 0.47 | **0.24** | **0.02** | **0.47** | **0.021** | **0.24** | **0.02** | **0.46** | **0.023** | 0.22 | -0.01 | 0.44 | 0.073 |
| ***Small HDL*** | | | | | | | | | | | | | | | |
| Particles | -0.02 | 0.22 | -0.07 | 0.18 | -0.03 | 0.39 | 0.204 | 0.19 | -0.01 | 0.40 | 0.107 | 0.16 | -0.04 | 0.37 | 0.390 |
| Total lipids | -0.01 | 0.36 | 0.07 | 0.20 | -0.01 | 0.41 | 0.102 | **0.21** | **0.00** | **0.41** | **0.049** | 0.18 | -0.03 | 0.39 | 0.214 |
| Phospholipids | -0.05 | 0.44 | 0.04 | 0.20 | -0.02 | 0.42 | 0.107 | 0.21 | 0.00 | 0.42 | 0.052 | 0.18 | -0.03 | 0.40 | 0.197 |
| Cholesterol | -0.03 | 0.26 | -0.07 | 0.15 | -0.06 | 0.37 | 0.648 | 0.16 | -0.05 | 0.38 | 0.444 | 0.14 | -0.08 | 0.35 | 0.818 |
| Cholesteryl esters | 0.00 | 0.40 | 0.17 | 0.13 | -0.08 | 0.35 | 0.872 | 0.15 | -0.07 | 0.36 | 0.672 | 0.12 | -0.09 | 0.34 | 0.949 |
| Free cholesterol | -0.06 | 0.57 | -0.07 | 0.18 | -0.03 | 0.38 | 0.233 | 0.18 | -0.02 | 0.38 | 0.175 | 0.16 | -0.05 | 0.36 | 0.423 |
| Triglycerides | 0.30 | 2.02 | 0.55 | **0.22** | **0.03** | **0.40** | **0.009** | **0.21** | **0.03** | **0.40** | **0.010** | 0.18 | -0.01 | 0.37 | 0.093 |

Note. K-effective test number: 51.881.

**Result Supplement Table 4. Direct and indirect associations (via body composition change) of menopausal hormonal shift and metabolite changes.** The effect sizes are the main results. They can be interpreted similarly to squared semi-partial correlations ($R^2$) as they describe how much of the metabolite change the menopausal hormonal shift (E2 decline and FSH increase) explains. E2 and FSH were included in the model simultaneously to better estimate the effect of menopausal hormonal shift. Therefore, their estimates provide standardised associations between the specific hormone level change and metabolite changes. However, they should be interpreted with caution due to the correlated nature of hormones. No statistically significant indirect associations between menopausal hormonal shift and metabolite changes were observed (light grey background), mainly because the menopausal hormonal shift did not associate with body fat percentage change. **Bolded metabolites** are outcomes of which direct effect *Q*-values (false discovery rate corrected *P*-values) were < 0.05. **Bolded metabolites with red font** are outcomes of which direct effect *Q*-values were < 0.05 and were statistically significantly associated with menopause in the primary analysis (**Result Supplement Table 3**).

| | **Direct effect** | | | | | | | | **Indirect effect via body fat percentage** | | | | | | | |
|---|---|---|---|---|---|---|---|---|---|---|---|---|---|---|---|---|
| | | | **E2** | | | **FSH** | | | | | **E2** | | | **FSH** | | |
| **Metabolite** | **Effect size** | ***Q*** | **Est.** | **S.E.** | ***Q*** | **Est.** | **S.E.** | ***Q*** | **Effect size** | ***Q*** | **Est.** | **S.E.** | ***Q*** | **Est.** | **S.E.** | ***Q*** |
| ***Cholesterols*** | | | | | | | | | | | | | | | | |
| **Total cholesterol** | **0.086** | **0.030** | **-0.108** | **0.065** | **0.366** | **0.038** | **0.035** | **0.552** | 0.00019 | 0.197 | -0.040 | 0.024 | 0.366 | 0.002 | 0.012 | 0.937 |
| **Non-HDL cholesterol** | **0.089** | **0.006** | **-0.135** | **0.058** | **0.217** | **0.030** | **0.031** | **0.616** | 0.00034 | 0.197 | -0.043 | 0.025 | 0.366 | 0.002 | 0.013 | 0.937 |
| **Remnant cholesterol** | **0.064** | **0.045** | **-0.134** | **0.059** | **0.235** | **0.004** | **0.032** | **0.938** | 0.00032 | 0.197 | -0.043 | 0.025 | 0.366 | 0.002 | 0.013 | 0.937 |
| **VLDL cholesterol** | **0.050** | **0.024** | **-0.158** | **0.057** | **0.102** | **-0.011** | **0.031** | **0.937** | 0.00042 | 0.197 | -0.041 | 0.024 | 0.366 | 0.002 | 0.012 | 0.937 |
| **Clinical LDL cholesterol** | **0.098** | **0.009** | **-0.118** | **0.061** | **0.366** | **0.042** | **0.033** | **0.460** | 0.00026 | 0.197 | -0.038 | 0.023 | 0.366 | 0.002 | 0.012 | 0.937 |
| **LDL cholesterol** | **0.106** | **0.004** | **-0.133** | **0.059** | **0.241** | **0.048** | **0.032** | **0.378** | 0.00032 | 0.197 | -0.041 | 0.024 | 0.366 | 0.002 | 0.012 | 0.937 |
| HDL cholesterol | 0.010 | 0.606 | 0.043 | 0.063 | 0.817 | 0.037 | 0.034 | 0.539 | 0.00007 | 0.999 | 0.000 | 0.008 | 0.977 | 0.000 | 0.000 | 0.977 |
| ***Triglycerides*** | | | | | | | | | | | | | | | | |
| **Total triglycerides** | **0.022** | **0.024** | **-0.205** | **0.067** | **0.07** | **-0.035** | **0.036** | **0.614** | 0.00041 | 0.197 | -0.044 | 0.026 | 0.366 | 0.002 | 0.013 | 0.937 |
| **VLDL triglycerides** | **0.029** | **0.006** | **-0.245** | **0.068** | **0.055** | **-0.039** | **0.036** | **0.549** | 0.00042 | 0.197 | -0.044 | 0.027 | 0.366 | 0.002 | 0.013 | 0.937 |
| LDL triglycerides | 0.018 | 0.401 | -0.088 | 0.065 | 0.425 | -0.015 | 0.034 | 0.937 | 0.00021 | 0.211 | -0.034 | 0.021 | 0.366 | 0.002 | 0.010 | 0.937 |
| HDL triglycerides | 0.004 | 0.539 | -0.085 | 0.077 | 0.532 | -0.013 | 0.041 | 0.937 | 0.00010 | 0.278 | -0.027 | 0.018 | 0.378 | 0.001 | 0.008 | 0.937 |
| ***Phospholipids*** | | | | | | | | | | | | | | | | |
| Total phospholipids | 0.057 | 0.108 | -0.107 | 0.070 | 0.37 | 0.026 | 0.038 | 0.808 | 0.00014 | 0.211 | -0.037 | 0.023 | 0.366 | 0.002 | 0.011 | 0.937 |
| **VLDL phospholipids** | **0.042** | **0.022** | **-0.180** | **0.059** | **0.07** | **-0.025** | **0.032** | **0.735** | 0.00048 | 0.197 | -0.044 | 0.026 | 0.366 | 0.002 | 0.013 | 0.937 |
| **LDL phospholipids** | **0.095** | **0.008** | **-0.126** | **0.060** | **0.309** | **0.037** | **0.032** | **0.517** | 0.00029 | 0.197 | -0.040 | 0.024 | 0.366 | 0.002 | 0.012 | 0.937 |
| HDL phospholipids | 0.009 | 0.699 | 0.011 | 0.066 | 0.937 | 0.029 | 0.035 | 0.721 | 0.00003 | 0.912 | -0.005 | 0.009 | 0.937 | 0.000 | 0.001 | 0.937 |
| ***Cholesteryl esters*** | | | | | | | | | | | | | | | | |
| **Total cholesteryl esters** | **0.086** | **0.038** | **-0.104** | **0.067** | **0.37** | **0.040** | **0.036** | **0.538** | 0.00017 | 0.206 | -0.038 | 0.023 | 0.366 | 0.002 | 0.012 | 0.937 |
| **VLDL cholesteryl esters** | **0.052** | **0.040** | **-0.145** | **0.057** | **0.153** | **-0.009** | **0.030** | **0.937** | 0.00036 | 0.197 | -0.037 | 0.022 | 0.366 | 0.002 | 0.011 | 0.937 |
| **LDL cholesteryl esters** | **0.108** | **0.003** | **-0.142** | **0.057** | **0.185** | **0.047** | **0.031** | **0.370** | 0.00037 | 0.197 | -0.042 | 0.025 | 0.366 | 0.002 | 0.013 | 0.937 |
| HDL cholesteryl esters | 0.011 | 0.556 | 0.046 | 0.063 | 0.774 | 0.040 | 0.034 | 0.494 | 0.00008 | 0.997 | 0.002 | 0.008 | 0.937 | 0.000 | 0.001 | 0.943 |

| Metabolite | Direct Effect size | Q | E2 Est. | S.E. | Q | FSH Est. | S.E. | Q | Indirect Effect size | Q | E2 Est. | S.E. | Q | FSH Est. | S.E. | Q |
|---|---|---|---|---|---|---|---|---|---|---|---|---|---|---|---|---|
| ***Free cholesterol*** | | | | | | | | | | | | | | | | |
| **Total free cholesterol** | **0.081** | **0.022** | **-0.116** | **0.061** | **0.366** | **0.031** | **0.033** | **0.631** | 0.00026 | 0.197 | -0.043 | 0.026 | 0.366 | 0.002 | 0.013 | 0.937 |
| **VLDL free cholesterol** | **0.048** | **0.016** | **-0.176** | **0.059** | **0.07** | **-0.014** | **0.032** | **0.937** | 0.00048 | 0.197 | -0.045 | 0.026 | 0.366 | 0.002 | 0.014 | 0.937 |
| **LDL free cholesterol** | **0.095** | **0.024** | **-0.104** | **0.066** | **0.366** | **0.044** | **0.035** | **0.469** | 0.00019 | 0.211 | -0.035 | 0.021 | 0.366 | 0.002 | 0.011 | 0.937 |
| HDL free cholesterol | 0.006 | 0.799 | 0.033 | 0.063 | 0.937 | 0.025 | 0.033 | 0.773 | 0.00004 | 0.954 | -0.003 | 0.008 | 0.937 | 0.000 | 0.001 | 0.937 |
| ***Lipids*** | | | | | | | | | | | | | | | | |
| **Total lipids** | **0.071** | **0.017** | **-0.155** | **0.064** | **0.195** | **0.015** | **0.035** | **0.937** | 0.00035 | 0.197 | -0.050 | 0.029 | 0.366 | 0.002 | 0.015 | 0.937 |
| **VLDL total lipids** | **0.041** | **0.011** | **-0.212** | **0.063** | **0.07** | **-0.032** | **0.034** | **0.631** | 0.00049 | 0.197 | -0.047 | 0.028 | 0.366 | 0.002 | 0.014 | 0.937 |
| **LDL total lipids** | **0.103** | **0.004** | **-0.132** | **0.058** | **0.235** | **0.044** | **0.032** | **0.409** | 0.00033 | 0.197 | -0.041 | 0.024 | 0.366 | 0.002 | 0.013 | 0.937 |
| HDL total lipids | 0.01 | 0.673 | 0.021 | 0.064 | 0.937 | 0.032 | 0.034 | 0.644 | 0.00004 | 0.941 | -0.004 | 0.009 | 0.937 | 0.000 | 0.001 | 0.937 |
| ***Particles*** | | | | | | | | | | | | | | | | |
| Total particles | 0.065 | 0.073 | -0.063 | 0.075 | 0.701 | 0.062 | 0.040 | 0.370 | 0.00002 | 0.305 | -0.024 | 0.017 | 0.394 | 0.001 | 0.007 | 0.937 |
| **VLDL particles** | **0.040** | **0.046** | **-0.166** | **0.059** | **0.1** | **-0.028** | **0.032** | **0.663** | 0.00042 | 0.197 | -0.042 | 0.025 | 0.366 | 0.002 | 0.013 | 0.937 |
| **LDL particles** | **0.085** | **0.005** | **-0.129** | **0.053** | **0.191** | **0.030** | **0.029** | **0.563** | 0.00036 | 0.197 | -0.039 | 0.023 | 0.366 | 0.002 | 0.012 | 0.937 |
| HDL particles | 0.055 | 0.113 | -0.042 | 0.074 | 0.906 | 0.064 | 0.040 | 0.366 | 0.00001 | 0.393 | -0.018 | 0.014 | 0.448 | 0.001 | 0.006 | 0.937 |
| ***Particle diameter*** | | | | | | | | | | | | | | | | |
| **VLDL size** | **0.028** | **0.005** | **-0.236** | **0.066** | **0.055** | **-0.023** | **0.035** | **0.857** | 0.00048 | 0.197 | -0.045 | 0.027 | 0.366 | 0.002 | 0.014 | 0.937 |
| LDL size | 0.008 | 0.592 | -0.008 | 0.092 | 0.958 | 0.039 | 0.049 | 0.723 | 0.00000 | 0.821 | -0.009 | 0.013 | 0.823 | 0.000 | 0.003 | 0.937 |
| HDL size | 0.019 | 0.078 | 0.099 | 0.054 | 0.366 | -0.014 | 0.029 | 0.937 | 0.00026 | 0.366 | 0.015 | 0.011 | 0.432 | -0.001 | 0.005 | 0.937 |
| ***Phosphoglycerides*** | | | | | | | | | | | | | | | | |
| Phosphoglycerides | 0.025 | 0.310 | -0.133 | 0.077 | 0.366 | -0.043 | 0.041 | 0.567 | 0.00009 | 0.251 | -0.030 | 0.020 | 0.370 | 0.001 | 0.009 | 0.937 |
| Tri/phosphoglyceride ratio | 0.008 | 0.080 | -0.167 | 0.068 | 0.187 | -0.021 | 0.036 | 0.906 | 0.00033 | 0.211 | -0.036 | 0.022 | 0.366 | 0.002 | 0.011 | 0.937 |
| Total cholines | 0.025 | 0.391 | -0.118 | 0.078 | 0.37 | -0.038 | 0.042 | 0.660 | 0.00007 | 0.264 | -0.029 | 0.019 | 0.370 | 0.001 | 0.009 | 0.937 |
| Phosphatidylcholines | 0.020 | 0.368 | -0.123 | 0.078 | 0.366 | -0.047 | 0.042 | 0.527 | 0.00008 | 0.249 | -0.031 | 0.020 | 0.370 | 0.001 | 0.010 | 0.937 |
| Sphingomyelins | 0.053 | 0.267 | -0.089 | 0.071 | 0.463 | 0.009 | 0.038 | 0.937 | 0.00006 | 0.263 | -0.026 | 0.017 | 0.370 | 0.001 | 0.008 | 0.937 |
| ***Apolipoproteins*** | | | | | | | | | | | | | | | | |
| **Apolipoprotein B** | **0.074** | **0.012** | **-0.130** | **0.054** | **0.195** | **0.017** | **0.029** | **0.896** | 0.00036 | 0.197 | -0.040 | 0.024 | 0.366 | 0.002 | 0.012 | 0.937 |
| Apolipoprotein A-I | 0.023 | 0.368 | -0.003 | 0.068 | 0.977 | 0.044 | 0.036 | 0.490 | 0.00001 | 0.699 | -0.009 | 0.010 | 0.671 | 0.000 | 0.003 | 0.937 |
| Apo B/A-I ratio | 0.026 | 0.197 | -0.098 | 0.053 | 0.366 | -0.009 | 0.028 | 0.937 | 0.00029 | 0.218 | -0.025 | 0.016 | 0.366 | 0.001 | 0.008 | 0.937 |
| ***Fatty acids*** | | | | | | | | | | | | | | | | |
| **Total fatty acids** | **0.048** | **0.028** | **-0.226** | **0.070** | **0.07** | **-0.068** | **0.037** | **0.366** | 0.00037 | 0.197 | -0.046 | 0.027 | 0.366 | 0.002 | 0.014 | 0.937 |
| Degree of unsaturation | 0.001 | 0.950 | 0.011 | 0.087 | 0.939 | -0.010 | 0.047 | 0.937 | 0.00005 | 0.506 | 0.017 | 0.015 | 0.516 | -0.001 | 0.005 | 0.937 |
| Omega-3 fatty acids | 0.022 | 0.075 | -0.206 | 0.083 | 0.18 | -0.115 | 0.044 | 0.149 | 0.00006 | 0.691 | -0.011 | 0.013 | 0.663 | 0.001 | 0.003 | 0.937 |

| | Direct | | E2 | | | FSH | | | Indirect | | E2 | | | FSH | | |
|---|---|---|---|---|---|---|---|---|---|---|---|---|---|---|---|---|
| **Metabolite** | **Effect size** | **Q** | **Est.** | **S.E.** | **Q** | **Est.** | **S.E.** | **Q** | **Effect size** | **Q** | **Est.** | **S.E.** | **Q** | **Est.** | **S.E.** | **Q** |
| ***Fatty acids continue*** | | | | | | | | | | | | | | | | |
| Omega-6 fatty acids | 0.031 | 0.226 | -0.159 | 0.082 | 0.366 | -0.068 | 0.044 | 0.370 | 0.00017 | 0.226 | -0.037 | 0.023 | 0.366 | 0.002 | 0.011 | 0.937 |
| Polyunsaturated fatty acids | 0.032 | 0.197 | -0.183 | 0.080 | 0.225 | -0.081 | 0.043 | 0.366 | 0.00017 | 0.227 | -0.035 | 0.022 | 0.366 | 0.002 | 0.011 | 0.937 |
| Monounsaturated fatty acids | 0.029 | 0.051 | -0.212 | 0.071 | 0.07 | -0.071 | 0.038 | 0.366 | 0.00040 | 0.2 | -0.042 | 0.026 | 0.366 | 0.002 | 0.013 | 0.937 |
| **Saturated fatty acids** | **0.049** | **0.032** | **-0.202** | **0.070** | **0.079** | **-0.030** | **0.037** | **0.727** | 0.00041 | 0.197 | -0.051 | 0.030 | 0.366 | 0.002 | 0.016 | 0.937 |
| **Linoleic acid** | **0.037** | **0.040** | **-0.220** | **0.116** | **0.366** | **-0.192** | **0.062** | **0.070** | 0.00004 | 0.483 | -0.024 | 0.020 | 0.499 | 0.001 | 0.007 | 0.937 |
| **Docosahexaenoic acid** | **0.030** | **0.033** | **-0.206** | **0.089** | **0.225** | **-0.150** | **0.048** | **0.070** | 0.00000 | 0.997 | 0.002 | 0.012 | 0.937 | 0.000 | 0.001 | 0.941 |
| ***Fatty acid ratios (%)*** | | | | | | | | | | | | | | | | |
| Omega-3 ratio | 0.017 | 0.197 | -0.135 | 0.088 | 0.37 | -0.105 | 0.047 | 0.242 | 0.00000 | 0.756 | 0.010 | 0.013 | 0.735 | 0.000 | 0.003 | 0.937 |
| Omega-6 ratio | 0.012 | 0.051 | 0.184 | 0.076 | 0.19 | 0.009 | 0.041 | 0.937 | 0.00028 | 0.211 | 0.040 | 0.025 | 0.366 | -0.002 | 0.012 | 0.937 |
| Polyunsaturated ratio | 0.010 | 0.123 | 0.112 | 0.080 | 0.408 | -0.033 | 0.043 | 0.745 | 0.00028 | 0.211 | 0.043 | 0.027 | 0.366 | -0.002 | 0.013 | 0.937 |
| Monounsaturated ratio | 0.003 | 0.379 | -0.113 | 0.073 | 0.37 | -0.041 | 0.039 | 0.562 | 0.00022 | 0.291 | -0.024 | 0.017 | 0.386 | 0.001 | 0.007 | 0.937 |
| Saturated ratio | 0.025 | 0.072 | -0.092 | 0.125 | 0.774 | 0.110 | 0.067 | 0.366 | 0.00004 | 0.391 | -0.031 | 0.024 | 0.447 | 0.001 | 0.010 | 0.937 |
| **Linoleic acid ratio** | **0.034** | **0.005** | **0.130** | **0.112** | **0.51** | **-0.135** | **0.060** | **0.235** | 0.00012 | 0.261 | 0.042 | 0.028 | 0.370 | -0.002 | 0.013 | 0.937 |
| Docosahexaenoic acid ratio | 0.026 | 0.074 | 0.002 | 0.083 | 0.987 | -0.094 | 0.044 | 0.270 | 0.00013 | 0.339 | 0.027 | 0.018 | 0.379 | 0.000 | 0.007 | 0.979 |
| Poly-/monounsaturated ratio | 0.003 | 0.280 | 0.114 | 0.072 | 0.366 | 0.013 | 0.039 | 0.937 | 0.00028 | 0.226 | 0.032 | 0.021 | 0.366 | -0.001 | 0.010 | 0.937 |
| Omega-6 to omega-3 ratio | 0.016 | 0.197 | 0.173 | 0.088 | 0.366 | 0.105 | 0.047 | 0.241 | 0.00001 | 0.999 | 0.000 | 0.011 | 0.979 | 0.000 | 0.001 | 0.979 |
| ***Amino acids*** | | | | | | | | | | | | | | | | |
| **Alanine** | **0.041** | **0.045** | **-0.260** | **0.086** | **0.07** | **-0.074** | **0.046** | **0.366** | 0.00018 | 0.261 | -0.032 | 0.021 | 0.370 | 0.001 | 0.010 | 0.937 |
| **Glutamine** | **0.024** | **0.041** | **-0.208** | **0.155** | **0.434** | **-0.247** | **0.083** | **0.070** | 0.00001 | 0.464 | 0.033 | 0.027 | 0.489 | -0.001 | 0.010 | 0.937 |
| Glycine | 0.026 | 0.108 | -0.091 | 0.056 | 0.366 | 0.017 | 0.030 | 0.896 | 0.00001 | 0.711 | -0.007 | 0.008 | 0.688 | 0.000 | 0.002 | 0.937 |
| Histidine | 0.022 | 0.249 | -0.176 | 0.093 | 0.366 | -0.071 | 0.050 | 0.397 | 0.00001 | 0.999 | -0.001 | 0.012 | 0.945 | 0.000 | 0.001 | 0.958 |
| Total BCAA | 0.016 | 0.197 | -0.211 | 0.098 | 0.27 | -0.053 | 0.052 | 0.570 | 0.00007 | 0.387 | -0.025 | 0.019 | 0.445 | 0.001 | 0.008 | 0.937 |
| Isoleucine | 0.023 | 0.079 | -0.243 | 0.091 | 0.126 | -0.053 | 0.049 | 0.539 | 0.00007 | 0.522 | -0.017 | 0.015 | 0.523 | 0.001 | 0.005 | 0.937 |
| Leucine | 0.026 | 0.072 | -0.243 | 0.096 | 0.159 | -0.031 | 0.051 | 0.871 | 0.00009 | 0.382 | -0.025 | 0.019 | 0.441 | 0.001 | 0.008 | 0.937 |
| Valine | 0.009 | 0.357 | -0.156 | 0.097 | 0.366 | -0.058 | 0.052 | 0.522 | 0.00004 | 0.4 | -0.024 | 0.018 | 0.451 | 0.001 | 0.007 | 0.937 |
| Phenylalanine | 0.052 | 0.116 | -0.220 | 0.093 | 0.215 | -0.032 | 0.050 | 0.861 | 0.00007 | 0.446 | -0.021 | 0.017 | 0.478 | 0.001 | 0.006 | 0.937 |
| **Tyrosine** | **0.078** | **0.022** | **-0.279** | **0.088** | **0.07** | **-0.051** | **0.047** | **0.541** | 0.00007 | 0.291 | -0.030 | 0.020 | 0.386 | 0.001 | 0.009 | 0.937 |
| ***Glycolysis-related*** | | | | | | | | | | | | | | | | |
| Glucose | 0.020 | 0.382 | -0.139 | 0.092 | 0.37 | -0.059 | 0.049 | 0.489 | 0.00010 | 0.313 | -0.029 | 0.020 | 0.400 | 0.001 | 0.009 | 0.937 |
| Lactate | 0.002 | 0.886 | -0.054 | 0.104 | 0.937 | -0.007 | 0.056 | 0.939 | 0.00010 | 0.329 | -0.031 | 0.022 | 0.409 | 0.001 | 0.010 | 0.937 |
| Pyruvate | 0.012 | 0.446 | -0.117 | 0.112 | 0.562 | -0.075 | 0.054 | 0.419 | 0.00009 | 0.506 | -0.025 | 0.021 | 0.490 | -0.001 | 0.006 | 0.937 |
| **Citrate** | **0.050** | **0.005** | **0.109** | **0.096** | **0.518** | **-0.117** | **0.051** | **0.225** | 0.00013 | 0.401 | -0.024 | 0.018 | 0.451 | 0.001 | 0.007 | 0.937 |
| Glycerol | 0.038 | 0.069 | -0.131 | 0.100 | 0.445 | 0.060 | 0.053 | 0.522 | 0.00017 | 0.249 | -0.040 | 0.026 | 0.370 | 0.002 | 0.012 | 0.937 |

| | Direct | | E2 | | | FSH | | | Indirect | | E2 | | | FSH | | |
|---|---|---|---|---|---|---|---|---|---|---|---|---|---|---|---|---|
| **Metabolite** | **Effect size** | ***Q*** | **Est.** | **S.E.** | ***Q*** | **Est.** | **S.E.** | ***Q*** | **Effect size** | ***Q*** | **Est.** | **S.E.** | ***Q*** | **Est.** | **S.E.** | ***Q*** |
| ***Ketone bodies*** | | | | | | | | | | | | | | | | |
| **3-hydroxybutyrate** | **0.044** | **0.029** | **0.071** | **0.066** | **0.552** | **-0.059** | **0.035** | **0.366** | 0.00000 | 0.919 | 0.004 | 0.009 | 0.937 | 0.000 | 0.001 | 0.937 |
| Acetate | 0.023 | 0.211 | 0.203 | 0.114 | 0.366 | 0.121 | 0.061 | 0.366 | 0.00000 | 0.999 | -0.001 | 0.015 | 0.979 | 0.000 | 0.001 | 0.979 |
| **Acetoacetate** | **0.034** | **0.041** | **0.130** | **0.098** | **0.438** | **-0.068** | **0.052** | **0.448** | 0.00000 | 0.999 | -0.001 | 0.013 | 0.968 | 0.000 | 0.001 | 0.971 |
| Acetone | 0.005 | 0.764 | 0.070 | 0.093 | 0.764 | 0.012 | 0.050 | 0.937 | 0.00000 | 0.778 | 0.010 | 0.013 | 0.767 | 0.000 | 0.003 | 0.937 |
| ***Miscellaneous*** | | | | | | | | | | | | | | | | |
| Creatinine | 0.029 | 0.112 | -0.115 | 0.066 | 0.366 | 0.014 | 0.035 | 0.937 | 0.00000 | 0.983 | -0.002 | 0.009 | 0.937 | 0.000 | 0.001 | 0.938 |
| **Albumin** | **0.090** | **0.006** | **-0.055** | **0.087** | **0.857** | **0.121** | **0.046** | **0.145** | 0.00003 | 0.368 | 0.024 | 0.017 | 0.433 | -0.001 | 0.007 | 0.937 |
| Glycoprotein acetyls | 0.032 | 0.069 | -0.208 | 0.076 | 0.116 | -0.041 | 0.041 | 0.577 | 0.00028 | 0.291 | -0.026 | 0.018 | 0.386 | 0.001 | 0.008 | 0.937 |
| ***Chylomicrons and extremely large VLDL*** | | | | | | | | | | | | | | | | |
| Particles | 0.012 | 0.197 | -0.146 | 0.078 | 0.366 | -0.005 | 0.042 | 0.938 | 0.00037 | 0.209 | -0.044 | 0.027 | 0.366 | 0.002 | 0.013 | 0.937 |
| Total lipids | 0.005 | 0.216 | -0.151 | 0.080 | 0.366 | -0.023 | 0.043 | 0.933 | 0.00024 | 0.254 | -0.031 | 0.020 | 0.370 | 0.001 | 0.010 | 0.937 |
| Phospholipids | 0.004 | 0.155 | -0.146 | 0.077 | 0.366 | 0.001 | 0.041 | 0.985 | 0.00041 | 0.206 | -0.044 | 0.027 | 0.366 | 0.002 | 0.013 | 0.937 |
| Cholesterol | 0.007 | 0.254 | -0.137 | 0.073 | 0.366 | -0.038 | 0.039 | 0.616 | 0.00028 | 0.228 | -0.032 | 0.021 | 0.366 | 0.001 | 0.010 | 0.937 |
| Cholesteryl esters | 0.010 | 0.217 | -0.151 | 0.074 | 0.352 | -0.053 | 0.039 | 0.430 | 0.00026 | 0.249 | -0.029 | 0.019 | 0.370 | 0.001 | 0.009 | 0.937 |
| Free cholesterol | 0.003 | 0.345 | -0.075 | 0.084 | 0.663 | 0.022 | 0.045 | 0.937 | 0.00028 | 0.218 | -0.040 | 0.025 | 0.366 | 0.002 | 0.012 | 0.937 |
| Triglycerides | 0.001 | 0.217 | -0.134 | 0.085 | 0.366 | 0.000 | 0.045 | 0.999 | 0.00025 | 0.249 | -0.034 | 0.022 | 0.370 | 0.002 | 0.010 | 0.937 |
| ***Very large VLDL*** | | | | | | | | | | | | | | | | |
| **Particles** | **0.023** | **0.017** | **-0.194** | **0.065** | **0.07** | **-0.017** | **0.035** | **0.937** | 0.00043 | 0.197 | -0.041 | 0.025 | 0.366 | 0.002 | 0.013 | 0.937 |
| **Total lipids** | **0.028** | **0.011** | **-0.210** | **0.066** | **0.07** | **-0.020** | **0.035** | **0.906** | 0.00043 | 0.197 | -0.041 | 0.025 | 0.366 | 0.002 | 0.013 | 0.937 |
| **Phospholipids** | **0.035** | **0.006** | **-0.215** | **0.065** | **0.07** | **-0.014** | **0.035** | **0.937** | 0.00040 | 0.197 | -0.040 | 0.024 | 0.366 | 0.002 | 0.012 | 0.937 |
| **Cholesterol** | **0.030** | **0.023** | **-0.185** | **0.061** | **0.07** | **-0.028** | **0.032** | **0.688** | 0.00047 | 0.197 | -0.040 | 0.024 | 0.366 | 0.002 | 0.012 | 0.937 |
| **Cholesteryl esters** | **0.031** | **0.028** | **-0.181** | **0.059** | **0.07** | **-0.037** | **0.031** | **0.490** | 0.00045 | 0.197 | -0.038 | 0.023 | 0.366 | 0.002 | 0.011 | 0.937 |
| **Free cholesterol** | **0.029** | **0.016** | **-0.191** | **0.066** | **0.076** | **-0.010** | **0.035** | **0.937** | 0.00043 | 0.197 | -0.041 | 0.025 | 0.366 | 0.002 | 0.013 | 0.937 |
| **Triglycerides** | **0.024** | **0.009** | **-0.229** | **0.072** | **0.07** | **-0.018** | **0.038** | **0.937** | 0.00033 | 0.211 | -0.038 | 0.023 | 0.366 | 0.002 | 0.012 | 0.937 |
| ***Large VLDL*** | | | | | | | | | | | | | | | | |
| **Particles** | **0.028** | **0.012** | **-0.213** | **0.064** | **0.07** | **-0.032** | **0.034** | **0.642** | 0.00044 | 0.197 | -0.042 | 0.025 | 0.366 | 0.002 | 0.013 | 0.937 |
| **Total lipids** | **0.034** | **0.005** | **-0.237** | **0.066** | **0.055** | **-0.031** | **0.035** | **0.671** | 0.00044 | 0.197 | -0.043 | 0.026 | 0.366 | 0.002 | 0.013 | 0.937 |
| **Phospholipids** | **0.021** | **0.028** | **-0.189** | **0.064** | **0.07** | **-0.029** | **0.034** | **0.702** | 0.00042 | 0.197 | -0.039 | 0.024 | 0.366 | 0.002 | 0.012 | 0.937 |
| **Cholesterol** | **0.032** | **0.017** | **-0.188** | **0.062** | **0.07** | **-0.021** | **0.033** | **0.868** | 0.00044 | 0.197 | -0.040 | 0.024 | 0.366 | 0.002 | 0.012 | 0.937 |
| **Cholesteryl esters** | **0.034** | **0.022** | **-0.182** | **0.061** | **0.07** | **-0.022** | **0.033** | **0.840** | 0.00041 | 0.197 | -0.039 | 0.023 | 0.366 | 0.002 | 0.012 | 0.937 |
| **Free cholesterol** | **0.028** | **0.017** | **-0.191** | **0.063** | **0.07** | **-0.019** | **0.034** | **0.906** | 0.00044 | 0.197 | -0.041 | 0.024 | 0.366 | 0.002 | 0.012 | 0.937 |
| **Triglycerides** | **0.016** | **0.017** | **-0.213** | **0.068** | **0.07** | **-0.028** | **0.036** | **0.733** | 0.00048 | 0.197 | -0.045 | 0.027 | 0.366 | 0.002 | 0.014 | 0.937 |

| | Direct | | E2 | | | FSH | | | Indirect | | E2 | | | FSH | | |
|---|---|---|---|---|---|---|---|---|---|---|---|---|---|---|---|---|
| **Metabolite** | **Effect size** | **Q** | **Est.** | **S.E.** | **Q** | **Est.** | **S.E.** | **Q** | **Effect size** | **Q** | **Est.** | **S.E.** | **Q** | **Est.** | **S.E.** | **Q** |
| ***Medium VLDL*** | | | | | | | | | | | | | | | | |
| **Particles** | **0.055** | **0.012** | **-0.187** | **0.059** | **0.07** | **-0.020** | **0.032** | **0.868** | 0.00047 | 0.197 | -0.046 | 0.027 | 0.366 | 0.002 | 0.014 | 0.937 |
| **Total lipids** | **0.057** | **0.005** | **-0.226** | **0.062** | **0.055** | **-0.029** | **0.033** | **0.663** | 0.00049 | 0.197 | -0.048 | 0.028 | 0.366 | 0.002 | 0.015 | 0.937 |
| **Phospholipids** | **0.059** | **0.009** | **-0.189** | **0.058** | **0.07** | **-0.018** | **0.031** | **0.906** | 0.00046 | 0.197 | -0.046 | 0.027 | 0.366 | 0.002 | 0.014 | 0.937 |
| **Cholesterol** | **0.069** | **0.028** | **-0.127** | **0.058** | **0.266** | **0.015** | **0.031** | **0.937** | 0.00030 | 0.197 | -0.037 | 0.022 | 0.366 | 0.002 | 0.011 | 0.937 |
| Cholesteryl esters | 0.067 | 0.123 | -0.094 | 0.064 | 0.38 | 0.024 | 0.034 | 0.807 | 0.00017 | 0.223 | -0.029 | 0.019 | 0.366 | 0.001 | 0.009 | 0.937 |
| **Free cholesterol** | **0.063** | **0.012** | **-0.167** | **0.057** | **0.076** | **-0.005** | **0.031** | **0.937** | 0.00043 | 0.197 | -0.044 | 0.026 | 0.366 | 0.002 | 0.014 | 0.937 |
| **Triglycerides** | **0.038** | **0.005** | **-0.260** | **0.067** | **0.055** | **-0.045** | **0.036** | **0.463** | 0.00042 | 0.197 | -0.044 | 0.026 | 0.366 | 0.002 | 0.013 | 0.937 |
| ***Small VLDL*** | | | | | | | | | | | | | | | | |
| Particles | 0.033 | 0.059 | -0.170 | 0.061 | 0.102 | -0.034 | 0.033 | 0.562 | 0.00037 | 0.197 | -0.038 | 0.023 | 0.366 | 0.002 | 0.012 | 0.937 |
| **Total lipids** | **0.040** | **0.033** | **-0.180** | **0.061** | **0.07** | **-0.034** | **0.032** | **0.562** | 0.00038 | 0.197 | -0.040 | 0.024 | 0.366 | 0.002 | 0.012 | 0.937 |
| **Phospholipids** | **0.057** | **0.022** | **-0.168** | **0.057** | **0.07** | **-0.019** | **0.030** | **0.868** | 0.00039 | 0.197 | -0.040 | 0.024 | 0.366 | 0.002 | 0.012 | 0.937 |
| **Cholesterol** | **0.054** | **0.045** | **-0.144** | **0.056** | **0.153** | **-0.011** | **0.030** | **0.937** | 0.00033 | 0.197 | -0.034 | 0.021 | 0.366 | 0.002 | 0.010 | 0.937 |
| Cholesteryl esters | 0.046 | 0.083 | -0.138 | 0.057 | 0.191 | -0.016 | 0.030 | 0.937 | 0.00030 | 0.21 | -0.031 | 0.019 | 0.366 | 0.001 | 0.010 | 0.937 |
| **Free cholesterol** | **0.066** | **0.020** | **-0.149** | **0.056** | **0.126** | **-0.001** | **0.030** | **0.979** | 0.00036 | 0.197 | -0.038 | 0.023 | 0.366 | 0.002 | 0.012 | 0.937 |
| Triglycerides | 0.014 | 0.086 | -0.183 | 0.068 | 0.126 | -0.046 | 0.036 | 0.460 | 0.00026 | 0.212 | -0.036 | 0.022 | 0.366 | 0.002 | 0.011 | 0.937 |
| ***Very small VLDL*** | | | | | | | | | | | | | | | | |
| Particles | 0.032 | 0.374 | -0.091 | 0.060 | 0.37 | -0.021 | 0.032 | 0.856 | 0.00019 | 0.217 | -0.030 | 0.019 | 0.366 | 0.001 | 0.009 | 0.937 |
| Total lipids | 0.031 | 0.379 | -0.090 | 0.061 | 0.384 | -0.017 | 0.033 | 0.937 | 0.00017 | 0.222 | -0.029 | 0.018 | 0.366 | 0.001 | 0.009 | 0.937 |
| Phospholipids | 0.023 | 0.539 | -0.075 | 0.062 | 0.488 | -0.021 | 0.033 | 0.865 | 0.00014 | 0.254 | -0.024 | 0.016 | 0.370 | 0.001 | 0.007 | 0.937 |
| Cholesterol | 0.045 | 0.298 | -0.090 | 0.063 | 0.4 | -0.004 | 0.034 | 0.943 | 0.00013 | 0.229 | -0.028 | 0.018 | 0.366 | 0.001 | 0.008 | 0.937 |
| Cholesteryl esters | 0.048 | 0.305 | -0.088 | 0.065 | 0.435 | 0.000 | 0.035 | 1.000 | 0.00010 | 0.244 | -0.027 | 0.017 | 0.370 | 0.001 | 0.008 | 0.937 |
| Free cholesterol | 0.033 | 0.357 | -0.090 | 0.060 | 0.378 | -0.014 | 0.032 | 0.937 | 0.00017 | 0.223 | -0.028 | 0.018 | 0.366 | 0.001 | 0.008 | 0.937 |
| Triglycerides | 0.008 | 0.499 | -0.087 | 0.068 | 0.454 | -0.035 | 0.036 | 0.610 | 0.00013 | 0.254 | -0.026 | 0.017 | 0.370 | 0.001 | 0.008 | 0.937 |
| ***IDL*** | | | | | | | | | | | | | | | | |
| Particles | 0.038 | 0.436 | -0.098 | 0.069 | 0.408 | -0.029 | 0.037 | 0.745 | 0.00016 | 0.217 | -0.034 | 0.021 | 0.366 | 0.002 | 0.010 | 0.937 |
| Total lipids | 0.064 | 0.217 | -0.088 | 0.066 | 0.441 | 0.013 | 0.036 | 0.937 | 0.00013 | 0.211 | -0.035 | 0.021 | 0.366 | 0.002 | 0.011 | 0.937 |
| Phospholipids | 0.074 | 0.133 | -0.080 | 0.063 | 0.461 | 0.031 | 0.034 | 0.660 | 0.00014 | 0.217 | -0.031 | 0.019 | 0.366 | 0.001 | 0.010 | 0.937 |
| Cholesterol | 0.061 | 0.271 | -0.089 | 0.070 | 0.46 | 0.007 | 0.037 | 0.937 | 0.00011 | 0.217 | -0.034 | 0.021 | 0.366 | 0.002 | 0.010 | 0.937 |
| Cholesteryl esters | 0.062 | 0.267 | -0.095 | 0.071 | 0.435 | 0.004 | 0.038 | 0.943 | 0.00011 | 0.217 | -0.034 | 0.021 | 0.366 | 0.002 | 0.010 | 0.937 |
| Free cholesterol | 0.054 | 0.326 | -0.068 | 0.068 | 0.582 | 0.015 | 0.036 | 0.937 | 0.00010 | 0.226 | -0.030 | 0.019 | 0.366 | 0.001 | 0.009 | 0.937 |
| Triglycerides | 0.008 | 0.698 | -0.060 | 0.068 | 0.671 | -0.030 | 0.036 | 0.717 | 0.00011 | 0.254 | -0.026 | 0.017 | 0.370 | 0.001 | 0.008 | 0.937 |

| | Direct | | E2 | | | FSH | | | Indirect | | E2 | | | FSH | | |
|---|---|---|---|---|---|---|---|---|---|---|---|---|---|---|---|---|
| **Metabolite** | **Effect size** | ***Q*** | **Est.** | **S.E.** | ***Q*** | **Est.** | **S.E.** | ***Q*** | **Effect size** | ***Q*** | **Est.** | **S.E.** | ***Q*** | **Est.** | **S.E.** | ***Q*** |
| ***Large LDL*** | | | | | | | | | | | | | | | | |
| **Particles** | **0.088** | **0.006** | **-0.121** | **0.056** | **0.270** | **0.035** | **0.030** | **0.516** | 0.00033 | 0.197 | -0.040 | 0.024 | 0.366 | 0.002 | 0.012 | 0.937 |
| **Total lipids** | **0.107** | **0.005** | **-0.124** | **0.061** | **0.366** | **0.048** | **0.033** | **0.388** | 0.00027 | 0.197 | -0.039 | 0.024 | 0.366 | 0.002 | 0.012 | 0.937 |
| Phospholipids | 0.099 | 0.012 | -0.120 | 0.063 | 0.366 | 0.041 | 0.034 | 0.489 | 0.00025 | 0.197 | -0.039 | 0.023 | 0.366 | 0.002 | 0.012 | 0.937 |
| **Cholesterol** | **0.109** | **0.005** | **-0.125** | **0.063** | **0.366** | **0.051** | **0.034** | **0.370** | 0.00026 | 0.197 | -0.039 | 0.024 | 0.366 | 0.002 | 0.012 | 0.937 |
| **Cholesteryl esters** | **0.112** | **0.004** | **-0.134** | **0.061** | **0.266** | **0.051** | **0.033** | **0.370** | 0.00029 | 0.197 | -0.040 | 0.024 | 0.366 | 0.002 | 0.012 | 0.937 |
| **Free cholesterol** | **0.094** | **0.029** | **-0.097** | **0.067** | **0.394** | **0.048** | **0.036** | **0.445** | 0.00016 | 0.216 | -0.034 | 0.021 | 0.366 | 0.002 | 0.010 | 0.937 |
| Triglycerides | 0.015 | 0.606 | -0.068 | 0.066 | 0.571 | -0.012 | 0.035 | 0.937 | 0.00014 | 0.226 | -0.030 | 0.019 | 0.366 | 0.001 | 0.009 | 0.937 |
| ***Medium LDL*** | | | | | | | | | | | | | | | | |
| **Particles** | **0.066** | **0.014** | **-0.133** | **0.057** | **0.210** | **0.018** | **0.030** | **0.890** | 0.00000 | 0.198 | -0.035 | 0.021 | 0.363 | 0.002 | 0.011 | 0.935 |
| **Total lipids** | **0.092** | **0.003** | **-0.144** | **0.055** | **0.143** | **0.037** | **0.030** | **0.473** | 0.00000 | 0.198 | -0.042 | 0.025 | 0.363 | 0.002 | 0.013 | 0.935 |
| **Phospholipids** | **0.093** | **0.004** | **-0.140** | **0.057** | **0.184** | **0.035** | **0.031** | **0.517** | 0.00000 | 0.198 | -0.040 | 0.024 | 0.363 | 0.002 | 0.012 | 0.935 |
| **Cholesterol** | **0.091** | **0.003** | **-0.142** | **0.056** | **0.147** | **0.039** | **0.030** | **0.450** | 0.00000 | 0.198 | -0.042 | 0.025 | 0.363 | 0.002 | 0.013 | 0.935 |
| **Cholesteryl esters** | **0.085** | **0.003** | **-0.153** | **0.054** | **0.092** | **0.031** | **0.029** | **0.552** | 0.00000 | 0.198 | -0.043 | 0.025 | 0.363 | 0.002 | 0.013 | 0.935 |
| Free cholesterol | **0.096** | **0.012** | **-0.109** | **0.064** | **0.363** | **0.048** | **0.035** | **0.423** | 0.00000 | 0.211 | -0.035 | 0.021 | 0.363 | 0.002 | 0.011 | 0.935 |
| **Triglycerides** | **0.023** | **0.222** | **-0.113** | **0.063** | **0.363** | **-0.014** | **0.034** | **0.935** | 0.00000 | 0.201 | -0.037 | 0.023 | 0.363 | 0.002 | 0.011 | 0.935 |
| ***Small LDL*** | | | | | | | | | | | | | | | | |
| **Particles** | **0.062** | **0.020** | **-0.148** | **0.061** | **0.19** | **0.010** | **0.033** | **0.937** | 0.00028 | 0.21 | -0.034 | 0.021 | 0.366 | 0.002 | 0.010 | 0.937 |
| **Total lipids** | **0.075** | **0.009** | **-0.130** | **0.056** | **0.223** | **0.026** | **0.030** | **0.693** | 0.00036 | 0.197 | -0.041 | 0.024 | 0.366 | 0.002 | 0.012 | 0.937 |
| Phospholipids | 0.059 | 0.080 | -0.103 | 0.057 | 0.366 | 0.014 | 0.031 | 0.937 | 0.00027 | 0.197 | -0.037 | 0.022 | 0.366 | 0.002 | 0.011 | 0.937 |
| **Cholesterol** | **0.080** | **0.006** | **-0.132** | **0.058** | **0.225** | **0.032** | **0.031** | **0.569** | 0.00035 | 0.197 | -0.040 | 0.024 | 0.366 | 0.002 | 0.012 | 0.937 |
| **Cholesteryl esters** | **0.083** | **0.004** | **-0.143** | **0.056** | **0.153** | **0.035** | **0.030** | **0.516** | 0.00040 | 0.197 | -0.042 | 0.025 | 0.366 | 0.002 | 0.013 | 0.937 |
| Free cholesterol | 0.066 | 0.073 | -0.105 | 0.063 | 0.366 | 0.024 | 0.034 | 0.795 | 0.00020 | 0.217 | -0.031 | 0.019 | 0.366 | 0.001 | 0.010 | 0.937 |
| Triglycerides | 0.018 | 0.155 | -0.151 | 0.066 | 0.225 | -0.026 | 0.035 | 0.768 | 0.00039 | 0.197 | -0.044 | 0.027 | 0.366 | 0.002 | 0.014 | 0.937 |
| ***Very large HDL*** | | | | | | | | | | | | | | | | |
| Particles | 0.006 | 0.315 | 0.077 | 0.060 | 0.451 | -0.002 | 0.032 | 0.978 | 0.00014 | 0.556 | 0.011 | 0.010 | 0.549 | 0.000 | 0.003 | 0.937 |
| Total lipids | 0.009 | 0.234 | 0.080 | 0.060 | 0.433 | -0.008 | 0.032 | 0.937 | 0.00018 | 0.391 | 0.015 | 0.012 | 0.447 | -0.001 | 0.005 | 0.937 |
| Phospholipids | 0.020 | 0.071 | 0.074 | 0.053 | 0.411 | -0.029 | 0.028 | 0.563 | 0.00022 | 0.379 | 0.014 | 0.010 | 0.441 | -0.001 | 0.004 | 0.937 |
| Cholesterol | 0.006 | 0.284 | 0.082 | 0.060 | 0.433 | -0.002 | 0.032 | 0.977 | 0.00017 | 0.423 | 0.014 | 0.011 | 0.463 | -0.001 | 0.004 | 0.937 |
| Cholesteryl esters | 0.005 | 0.291 | 0.082 | 0.060 | 0.425 | 0.000 | 0.032 | 0.997 | 0.00017 | 0.446 | 0.013 | 0.011 | 0.478 | -0.001 | 0.004 | 0.937 |
| Free cholesterol | 0.010 | 0.280 | 0.076 | 0.063 | 0.489 | -0.009 | 0.034 | 0.937 | 0.00015 | 0.375 | 0.017 | 0.013 | 0.435 | -0.001 | 0.005 | 0.937 |
| Triglycerides | 0.008 | 0.545 | -0.024 | 0.071 | 0.937 | -0.042 | 0.038 | 0.532 | 0.00005 | 0.313 | -0.022 | 0.016 | 0.400 | 0.001 | 0.007 | 0.937 |

| | Direct | | E2 | | | FSH | | | Indirect | | E2 | | | FSH | | |
|---|---|---|---|---|---|---|---|---|---|---|---|---|---|---|---|---|
| **Metabolite** | **Effect size** | ***Q*** | **Est.** | **S.E.** | ***Q*** | **Est.** | **S.E.** | ***Q*** | **Effect size** | ***Q*** | **Est.** | **S.E.** | ***Q*** | **Est.** | **S.E.** | ***Q*** |
| ***Large HDL*** | | | | | | | | | | | | | | | | |
| Particles | 0.001 | 0.450 | 0.069 | 0.057 | 0.478 | 0.008 | 0.030 | 0.937 | 0.00017 | 0.638 | 0.008 | 0.009 | 0.614 | 0.000 | 0.003 | 0.937 |
| Total lipids | 0.001 | 0.414 | 0.071 | 0.056 | 0.46 | 0.007 | 0.030 | 0.937 | 0.00018 | 0.59 | 0.009 | 0.009 | 0.567 | 0.000 | 0.003 | 0.937 |
| Phospholipids | 0.001 | 0.446 | 0.066 | 0.056 | 0.505 | 0.005 | 0.030 | 0.937 | 0.00017 | 0.612 | 0.009 | 0.009 | 0.588 | 0.000 | 0.003 | 0.937 |
| Cholesterol | 0.001 | 0.387 | 0.078 | 0.057 | 0.425 | 0.011 | 0.030 | 0.937 | 0.00019 | 0.531 | 0.011 | 0.010 | 0.531 | 0.000 | 0.003 | 0.937 |
| Cholesteryl esters | 0.001 | 0.383 | 0.080 | 0.057 | 0.408 | 0.013 | 0.030 | 0.937 | 0.00019 | 0.521 | 0.011 | 0.010 | 0.522 | -0.001 | 0.003 | 0.937 |
| Free cholesterol | 0.001 | 0.393 | 0.071 | 0.057 | 0.469 | 0.004 | 0.030 | 0.938 | 0.00017 | 0.574 | 0.010 | 0.009 | 0.562 | 0.000 | 0.003 | 0.937 |
| Triglycerides | 0.003 | 0.888 | 0.005 | 0.072 | 0.971 | -0.015 | 0.039 | 0.937 | 0.00000 | 0.58 | -0.012 | 0.012 | 0.562 | 0.001 | 0.004 | 0.937 |
| ***Medium HDL*** | | | | | | | | | | | | | | | | |
| Particles | 0.024 | 0.329 | -0.005 | 0.069 | 0.971 | 0.046 | 0.037 | 0.466 | 0.00001 | 0.772 | -0.008 | 0.010 | 0.759 | 0.000 | 0.002 | 0.937 |
| Total lipids | 0.026 | 0.282 | -0.016 | 0.070 | 0.937 | 0.046 | 0.038 | 0.475 | 0.00001 | 0.737 | -0.008 | 0.010 | 0.717 | 0.000 | 0.003 | 0.937 |
| Phospholipids | 0.026 | 0.242 | -0.028 | 0.073 | 0.937 | 0.048 | 0.039 | 0.480 | 0.00000 | 0.644 | -0.011 | 0.011 | 0.619 | 0.000 | 0.003 | 0.937 |
| Cholesterol | 0.023 | 0.375 | 0.011 | 0.067 | 0.937 | 0.047 | 0.036 | 0.445 | 0.00003 | 0.97 | -0.003 | 0.009 | 0.937 | 0.000 | 0.001 | 0.938 |
| Cholesteryl esters | 0.024 | 0.361 | 0.015 | 0.067 | 0.937 | 0.049 | 0.036 | 0.425 | 0.00004 | 0.997 | -0.002 | 0.009 | 0.937 | 0.000 | 0.001 | 0.943 |
| Free cholesterol | 0.020 | 0.446 | -0.002 | 0.069 | 0.985 | 0.039 | 0.037 | 0.557 | 0.00001 | 0.737 | -0.008 | 0.010 | 0.717 | 0.000 | 0.003 | 0.937 |
| Triglycerides | 0.005 | 0.423 | -0.091 | 0.077 | 0.499 | -0.004 | 0.041 | 0.949 | 0.00009 | 0.291 | -0.026 | 0.018 | 0.386 | 0.001 | 0.008 | 0.937 |
| ***Small HDL*** | | | | | | | | | | | | | | | | |
| **Particles** | **0.109** | **0.004** | **-0.135** | **0.070** | **0.366** | **0.065** | **0.038** | **0.366** | 0.00018 | 0.218 | -0.033 | 0.021 | 0.366 | 0.002 | 0.010 | 0.937 |
| **Total lipids** | **0.083** | **0.009** | **-0.142** | **0.072** | **0.366** | **0.049** | **0.039** | **0.460** | 0.00017 | 0.217 | -0.035 | 0.022 | 0.366 | 0.002 | 0.011 | 0.937 |
| **Phospholipids** | **0.066** | **0.017** | **-0.137** | **0.074** | **0.366** | **0.043** | **0.040** | **0.539** | 0.00014 | 0.225 | -0.034 | 0.021 | 0.366 | 0.002 | 0.010 | 0.937 |
| **Cholesterol** | **0.107** | **0.006** | **-0.124** | **0.073** | **0.366** | **0.068** | **0.039** | **0.366** | 0.00013 | 0.237 | -0.031 | 0.020 | 0.370 | 0.001 | 0.009 | 0.937 |
| **Cholesteryl esters** | **0.105** | **0.006** | **-0.108** | **0.074** | **0.38** | **0.077** | **0.039** | **0.366** | 0.00011 | 0.264 | -0.027 | 0.018 | 0.370 | 0.001 | 0.008 | 0.937 |
| **Free cholesterol** | **0.077** | **0.024** | **-0.130** | **0.074** | **0.366** | **0.041** | **0.039** | **0.563** | 0.00014 | 0.217 | -0.037 | 0.023 | 0.366 | 0.002 | 0.011 | 0.937 |
| Triglycerides | 0.007 | 0.235 | -0.118 | 0.069 | 0.366 | -0.013 | 0.037 | 0.937 | 0.00021 | 0.237 | -0.029 | 0.019 | 0.370 | 0.001 | 0.009 | 0.937 |

**Result Supplement Table 5. Associations between menopausal hormone therapy and metabolite measures**. Skew ($g_1$) and kurtosis ($g_2$) for distribution of residuals for linear mixed model analysis of interaction of time and group, lambda parameter used in Box-Cox transformation of metabolite distribution, estimate of change parameter, $K_{eff}$-Šidák-corrected confidence intervals and *P*-values in crude, covariate-adjusted model (excluding fat percentage) and full covariate adjusted linear mixed model. *P*-values < 0.05 are bolded.

| | | | | **Crude** | | | | **Adjusted** | | | | **Adjusted + body fat %** | | | |
|---|---|---|---|---|---|---|---|---|---|---|---|---|---|---|---|
| | | | | | **99.95% CI** | | | | **99.95% CI** | | | | **99.95% CI** | | |
| **Metabolite** | ***g*₁** | ***g*₂** | ***λ*** | **Est.** | **Lower** | **Upper** | ***P*** | **Est.** | **Lower** | **Upper** | ***P*** | **Est.** | **Lower** | **Upper** | ***P*** |
| ***Cholesterols*** | | | | | | | | | | | | | | | |
| Total cholesterol | 0.06 | 0.73 | 0.15 | -0.15 | -0.58 | 0.27 | 1.000 | -0.12 | -0.55 | 0.30 | 1.000 | -0.12 | -0.55 | 0.30 | 1.000 |
| Non-HDL cholesterol | 0.01 | 1.11 | 0.04 | -0.30 | -0.69 | 0.09 | 0.445 | -0.28 | -0.67 | 0.12 | 0.650 | -0.28 | -0.66 | 0.11 | 0.633 |
| Remnant cholesterol | 0.01 | 0.85 | 0.04 | -0.23 | -0.62 | 0.15 | 0.910 | -0.21 | -0.60 | 0.18 | 0.974 | -0.21 | -0.60 | 0.17 | 0.972 |
| VLDL cholesterol | -0.11 | 1.13 | -0.07 | -0.27 | -0.64 | 0.11 | 0.614 | -0.26 | -0.63 | 0.12 | 0.727 | -0.25 | -0.63 | 0.12 | 0.708 |
| Clinical LDL cholesterol | 0.06 | 1.16 | 0.18 | -0.38 | -0.80 | 0.04 | 0.149 | -0.35 | -0.78 | 0.07 | 0.267 | -0.35 | -0.78 | 0.07 | 0.259 |
| LDL cholesterol | 0.02 | 1.17 | 0.04 | -0.34 | -0.75 | 0.07 | 0.253 | -0.32 | -0.73 | 0.09 | 0.423 | -0.32 | -0.73 | 0.09 | 0.404 |
| HDL cholesterol | -0.22 | 0.32 | 0.06 | **0.36** | **0.02** | **0.70** | **0.028** | **0.39** | **0.05** | **0.74** | **0.008** | **0.39** | **0.05** | **0.74** | **0.010** |
| ***Triglycerides*** | | | | | | | | | | | | | | | |
| Total triglycerides | -0.07 | 0.24 | -0.55 | 0.01 | -0.47 | 0.49 | 1.000 | 0.03 | -0.45 | 0.51 | 1.000 | 0.03 | -0.44 | 0.49 | 1.000 |
| VLDL triglycerides | -0.10 | 0.46 | -0.33 | -0.09 | -0.57 | 0.39 | 1.000 | -0.07 | -0.56 | 0.41 | 1.000 | -0.07 | -0.54 | 0.39 | 1.000 |
| LDL triglycerides | -0.07 | 0.55 | -1.00 | 0.22 | -0.22 | 0.67 | 0.994 | 0.25 | -0.18 | 0.69 | 0.947 | 0.25 | -0.18 | 0.69 | 0.945 |
| HDL triglycerides | -0.06 | 0.93 | 0.06 | 0.45 | -0.13 | 1.02 | 0.400 | 0.47 | -0.09 | 1.04 | 0.259 | 0.47 | -0.09 | 1.03 | 0.255 |
| ***Phospholipids*** | | | | | | | | | | | | | | | |
| Total phospholipids | -0.09 | 0.69 | 0.09 | 0.21 | -0.26 | 0.68 | 0.999 | 0.25 | -0.22 | 0.71 | 0.984 | 0.25 | -0.21 | 0.71 | 0.982 |
| VLDL phospholipids | -0.10 | 1.07 | -0.20 | -0.18 | -0.58 | 0.21 | 0.999 | -0.17 | -0.57 | 0.23 | 1.000 | -0.17 | -0.56 | 0.22 | 1.000 |
| LDL phospholipids | 0.03 | 0.88 | 0.01 | -0.35 | -0.76 | 0.05 | 0.197 | -0.33 | -0.74 | 0.08 | 0.350 | -0.33 | -0.73 | 0.08 | 0.339 |
| HDL phospholipids | -0.34 | 0.77 | 0.02 | **0.56** | **0.15** | **0.98** | **0.000** | **0.60** | **0.19** | **1.01** | **0.000** | **0.60** | **0.18** | **1.01** | **0.000** |
| ***Cholesteryl esters*** | | | | | | | | | | | | | | | |
| Total cholesteryl esters | 0.07 | 0.78 | 0.20 | -0.14 | -0.58 | 0.30 | 1.000 | -0.11 | -0.55 | 0.33 | 1.000 | -0.11 | -0.55 | 0.33 | 1.000 |
| VLDL cholesteryl esters | -0.11 | 1.12 | 0.02 | -0.30 | -0.68 | 0.07 | 0.296 | -0.29 | -0.67 | 0.08 | 0.398 | -0.29 | -0.66 | 0.08 | 0.389 |
| LDL cholesteryl esters | -0.01 | 1.17 | 0.02 | -0.33 | -0.73 | 0.07 | 0.267 | -0.31 | -0.71 | 0.09 | 0.448 | -0.31 | -0.70 | 0.09 | 0.424 |
| HDL cholesteryl esters | -0.20 | 0.44 | 0.17 | 0.34 | -0.01 | 0.68 | 0.061 | **0.37** | **0.03** | **0.71** | **0.020** | **0.37** | **0.02** | **0.71** | **0.023** |
| ***Free cholesterol*** | | | | | | | | | | | | | | | |
| Total free cholesterol | 0.04 | 0.79 | 0.03 | -0.18 | -0.58 | 0.23 | 1.000 | -0.15 | -0.56 | 0.25 | 1.000 | -0.15 | -0.55 | 0.25 | 1.000 |
| VLDL free cholesterol | -0.11 | 1.09 | -0.17 | -0.22 | -0.60 | 0.17 | 0.963 | -0.21 | -0.60 | 0.18 | 0.985 | -0.21 | -0.59 | 0.18 | 0.981 |
| LDL free cholesterol | 0.06 | 1.73 | 0.12 | -0.37 | -0.82 | 0.08 | 0.299 | -0.35 | -0.80 | 0.11 | 0.455 | -0.35 | -0.80 | 0.11 | 0.448 |
| HDL free cholesterol | -0.28 | 0.41 | -0.10 | **0.43** | **0.07** | **0.79** | **0.004** | **0.46** | **0.10** | **0.81** | **0.001** | **0.46** | **0.10** | **0.81** | **0.001** |
| ***Total lipids*** | | | | | | | | | | | | | | | |
| Total lipids | -0.01 | 1.03 | -0.42 | -0.01 | -0.45 | 0.43 | 1.000 | 0.02 | -0.42 | 0.46 | 1.000 | 0.02 | -0.41 | 0.45 | 1.000 |
| VLDL total lipids | -0.09 | 0.70 | -0.34 | -0.17 | -0.59 | 0.26 | 1.000 | -0.15 | -0.58 | 0.27 | 1.000 | -0.15 | -0.57 | 0.26 | 1.000 |
| LDL total lipids | 0.02 | 1.01 | 0.00 | -0.32 | -0.72 | 0.08 | 0.347 | -0.30 | -0.70 | 0.11 | 0.552 | -0.29 | -0.69 | 0.10 | 0.534 |
| HDL total lipids | -0.33 | 0.66 | -0.02 | **0.50** | **0.12** | **0.88** | **0.001** | **0.53** | **0.15** | **0.91** | **0.000** | **0.53** | **0.15** | **0.92** | **0.000** |

| | | | | Crude | | | | Adjusted | | | | Adjusted + body fat % | | | |
|---|---|---|---|---|---|---|---|---|---|---|---|---|---|---|---|
| | | | | | 99.95% CI | | | | 99.95% CI | | | | 99.95% CI | | |
| **Metabolite** | $g_1$ | $g_2$ | $\lambda$ | **Est.** | **Lower** | **Upper** | ***P*** | **Est.** | **Lower** | **Upper** | ***P*** | **Est.** | **Lower** | **Upper** | ***P*** |
| ***Particles*** | | | | | | | | | | | | | | | |
| Total particles | -0.22 | 0.57 | 0.27 | 0.32 | -0.18 | 0.82 | 0.823 | 0.37 | -0.12 | 0.86 | 0.494 | 0.37 | -0.12 | 0.86 | 0.496 |
| VLDL particles | -0.13 | 1.00 | -0.34 | -0.14 | -0.54 | 0.25 | 1.000 | -0.13 | -0.53 | 0.27 | 1.000 | -0.13 | -0.52 | 0.26 | 1.000 |
| LDL particles | -0.02 | 1.52 | -0.18 | **-0.40** | **-0.76** | **-0.05** | **0.010** | **-0.39** | **-0.75** | **-0.03** | **0.020** | **-0.39** | **-0.74** | **-0.03** | **0.018** |
| HDL particles | -0.26 | 0.54 | 0.30 | 0.39 | -0.09 | 0.88 | 0.328 | 0.44 | -0.04 | 0.92 | 0.124 | 0.44 | -0.04 | 0.92 | 0.127 |
| ***Particle diameter*** | | | | | | | | | | | | | | | |
| VLDL size | 0.00 | 0.42 | -6.00 | -0.24 | -0.69 | 0.22 | 0.989 | -0.23 | -0.68 | 0.23 | 0.996 | -0.23 | -0.66 | 0.21 | 0.992 |
| LDL size | 0.03 | 0.06 | 6.00 | 0.12 | -0.54 | 0.78 | 1.000 | 0.12 | -0.54 | 0.77 | 1.000 | 0.12 | -0.54 | 0.77 | 1.000 |
| HDL size | -0.10 | -0.09 | -6.00 | **0.35** | **0.05** | **0.65** | **0.007** | **0.35** | **0.04** | **0.65** | **0.009** | **0.34** | **0.04** | **0.64** | **0.010** |
| ***Phosphoglycerides*** | | | | | | | | | | | | | | | |
| Phosphoglycerides | -0.11 | 0.69 | -0.14 | 0.35 | -0.16 | 0.86 | 0.687 | 0.39 | -0.11 | 0.89 | 0.395 | 0.39 | -0.11 | 0.89 | 0.389 |
| Tri-/phosphoglyceride ratio | -0.07 | 0.03 | -0.37 | -0.09 | -0.54 | 0.35 | 1.000 | -0.09 | -0.53 | 0.36 | 1.000 | -0.08 | -0.52 | 0.35 | 1.000 |
| Total cholines | -0.08 | 0.70 | 0.03 | 0.28 | -0.22 | 0.78 | 0.962 | 0.32 | -0.17 | 0.80 | 0.797 | 0.32 | -0.17 | 0.80 | 0.793 |
| Phosphatidylcholines | -0.09 | 0.73 | -0.06 | 0.40 | -0.12 | 0.92 | 0.421 | 0.44 | -0.07 | 0.94 | 0.206 | 0.44 | -0.07 | 0.94 | 0.203 |
| Sphingomyelins | 0.06 | 0.63 | 0.07 | -0.04 | -0.46 | 0.39 | 1.000 | -0.01 | -0.43 | 0.42 | 1.000 | -0.01 | -0.43 | 0.42 | 1.000 |
| ***Apolipoproteins*** | | | | | | | | | | | | | | | |
| Apolipoprotein B | 0.00 | 1.60 | -0.15 | -0.36 | -0.71 | 0.00 | 0.050 | -0.34 | -0.70 | 0.02 | 0.089 | -0.34 | -0.70 | 0.02 | 0.085 |
| Apolipoprotein A-I | -0.34 | 0.70 | 0.03 | **0.51** | **0.08** | **0.93** | **0.005** | **0.55** | **0.13** | **0.97** | **0.001** | **0.55** | **0.12** | **0.97** | **0.001** |
| Apo B/A-I ratio | -0.03 | 2.69 | -0.23 | **-0.54** | **-0.86** | **-0.21** | **0.000** | **-0.55** | **-0.88** | **-0.22** | **0.000** | **-0.55** | **-0.88** | **-0.22** | **0.000** |
| ***Fatty acids*** | | | | | | | | | | | | | | | |
| Total fatty acids | 0.03 | 0.77 | -1.29 | 0.08 | -0.39 | 0.55 | 1.000 | 0.11 | -0.35 | 0.56 | 1.000 | 0.11 | -0.35 | 0.56 | 1.000 |
| Degree of unsaturation | 0.11 | 0.58 | -0.04 | 0.15 | -0.33 | 0.63 | 1.000 | 0.13 | -0.35 | 0.61 | 1.000 | 0.13 | -0.35 | 0.61 | 1.000 |
| Omega-3 fatty acids | 0.18 | -0.10 | 0.09 | 0.29 | -0.17 | 0.75 | 0.856 | 0.29 | -0.17 | 0.76 | 0.841 | 0.29 | -0.17 | 0.76 | 0.840 |
| Omega-6 fatty acids | 0.11 | 1.76 | -0.59 | -0.01 | -0.51 | 0.49 | 1.000 | 0.01 | -0.48 | 0.51 | 1.000 | 0.01 | -0.48 | 0.50 | 1.000 |
| Polyunsaturated fatty acids | 0.08 | 1.07 | -0.66 | 0.07 | -0.40 | 0.54 | 1.000 | 0.10 | -0.37 | 0.56 | 1.000 | 0.10 | -0.36 | 0.56 | 1.000 |
| Monounsaturated fatty acids | 0.02 | 0.74 | -0.98 | -0.02 | -0.50 | 0.45 | 1.000 | 0.00 | -0.47 | 0.47 | 1.000 | 0.00 | -0.46 | 0.47 | 1.000 |
| Saturated fatty acids | -0.05 | 1.03 | -1.28 | 0.13 | -0.37 | 0.62 | 1.000 | 0.16 | -0.32 | 0.64 | 1.000 | 0.16 | -0.31 | 0.64 | 1.000 |
| Linoleic acid | 0.08 | 0.79 | -0.24 | -0.09 | -0.68 | 0.51 | 1.000 | -0.08 | -0.67 | 0.51 | 1.000 | -0.08 | -0.68 | 0.51 | 1.000 |
| Docosahexaenoic acid | 0.11 | 0.09 | -0.39 | 0.37 | -0.11 | 0.86 | 0.412 | 0.38 | -0.11 | 0.87 | 0.401 | 0.38 | -0.11 | 0.87 | 0.404 |
| ***Fatty acid ratios (%)*** | | | | | | | | | | | | | | | |
| Omega-3 ratio | 0.31 | 0.43 | 0.24 | 0.29 | -0.22 | 0.80 | 0.958 | 0.28 | -0.24 | 0.80 | 0.979 | 0.28 | -0.24 | 0.80 | 0.980 |
| Omega-6 ratio | 0.16 | 0.42 | 5.81 | -0.15 | -0.68 | 0.38 | 1.000 | -0.19 | -0.71 | 0.34 | 1.000 | -0.19 | -0.71 | 0.34 | 1.000 |
| Polyunsaturated ratio | 0.15 | 0.73 | 5.13 | -0.03 | -0.56 | 0.50 | 1.000 | -0.07 | -0.60 | 0.46 | 1.000 | -0.07 | -0.59 | 0.45 | 1.000 |
| Monounsaturated ratio | -0.01 | 1.23 | -0.52 | -0.13 | -0.61 | 0.35 | 1.000 | -0.11 | -0.59 | 0.38 | 1.000 | -0.11 | -0.59 | 0.37 | 1.000 |
| Saturated ratio | -0.24 | 1.71 | -1.10 | 0.24 | -0.43 | 0.92 | 1.000 | 0.29 | -0.38 | 0.96 | 1.000 | 0.29 | -0.38 | 0.96 | 1.000 |
| Linoleic acid ratio | 0.20 | 0.23 | 3.06 | -0.21 | -0.85 | 0.42 | 1.000 | -0.26 | -0.90 | 0.38 | 1.000 | -0.26 | -0.89 | 0.38 | 1.000 |
| Docosahexaenoic acid ratio | 0.15 | 0.17 | -0.02 | 0.32 | -0.17 | 0.82 | 0.793 | 0.31 | -0.19 | 0.81 | 0.879 | 0.31 | -0.19 | 0.81 | 0.873 |
| Polyu/monounsaturated ratio | 0.02 | 1.15 | 1.27 | 0.07 | -0.41 | 0.56 | 1.000 | 0.05 | -0.44 | 0.53 | 1.000 | 0.04 | -0.43 | 0.52 | 1.000 |
| Omega-6 to omega-3 ratio | -0.27 | 0.31 | -0.19 | -0.34 | -0.85 | 0.17 | 0.772 | -0.33 | -0.85 | 0.18 | 0.811 | -0.33 | -0.85 | 0.18 | 0.814 |

| | | | | | Crude 99.95% CI | | | | Adjusted 99.95% CI | | | | Adjusted + body fat % 99.95% CI | | |
|---|---|---|---|---|---|---|---|---|---|---|---|---|---|---|---|
| **Metabolite** | $g_1$ | $g_2$ | $\lambda$ | **Est.** | **Lower** | **Upper** | ***P*** | **Est.** | **Lower** | **Upper** | ***P*** | **Est.** | **Lower** | **Upper** | ***P*** |
| ***Amino acids*** | | | | | | | | | | | | | | | |
| Alanine | 0.08 | 0.07 | -0.13 | -0.01 | -0.62 | 0.59 | 1.000 | 0.00 | -0.60 | 0.61 | 1.000 | 0.00 | -0.59 | 0.59 | 1.000 |
| Glutamine | 0.14 | -0.06 | 0.71 | -0.59 | -1.23 | 0.05 | 0.122 | -0.60 | -1.24 | 0.05 | 0.107 | -0.60 | -1.24 | 0.05 | 0.108 |
| Glycine | 0.02 | 0.11 | -0.63 | **-0.46** | **-0.80** | **-0.11** | **0.001** | **-0.46** | **-0.80** | **-0.12** | **0.001** | **-0.46** | **-0.81** | **-0.11** | **0.001** |
| Histidine | 0.11 | 0.69 | 0.43 | 0.30 | -0.29 | 0.89 | 0.991 | 0.30 | -0.28 | 0.89 | 0.990 | 0.30 | -0.28 | 0.89 | 0.990 |
| Total BCAA | -0.19 | 0.05 | -0.48 | -0.18 | -0.79 | 0.43 | 1.000 | -0.20 | -0.82 | 0.42 | 1.000 | -0.20 | -0.82 | 0.42 | 1.000 |
| Isoleucine | -0.22 | 1.20 | 0.23 | -0.29 | -0.92 | 0.33 | 0.999 | -0.30 | -0.93 | 0.33 | 0.998 | -0.30 | -0.93 | 0.33 | 0.998 |
| Leucine | -0.15 | 0.02 | -0.09 | -0.22 | -0.82 | 0.37 | 1.000 | -0.23 | -0.84 | 0.37 | 1.000 | -0.23 | -0.83 | 0.37 | 1.000 |
| Valine | -0.09 | -0.11 | -0.40 | -0.09 | -0.71 | 0.53 | 1.000 | -0.12 | -0.75 | 0.50 | 1.000 | -0.12 | -0.75 | 0.51 | 1.000 |
| Phenylalanine | -0.07 | -0.44 | -0.01 | 0.11 | -0.52 | 0.75 | 1.000 | 0.10 | -0.54 | 0.74 | 1.000 | 0.10 | -0.54 | 0.74 | 1.000 |
| Tyrosine | 0.04 | 0.18 | -0.02 | -0.16 | -0.77 | 0.45 | 1.000 | -0.12 | -0.72 | 0.49 | 1.000 | -0.12 | -0.72 | 0.48 | 1.000 |
| ***Glycolysis-related*** | | | | | | | | | | | | | | | |
| Glucose | -0.44 | 0.53 | -1.22 | -0.17 | -0.71 | 0.38 | 1.000 | -0.11 | -0.65 | 0.43 | 1.000 | -0.11 | -0.65 | 0.43 | 1.000 |
| Lactate | 0.09 | 1.12 | -0.68 | 0.43 | -0.26 | 1.12 | 0.881 | 0.42 | -0.28 | 1.12 | 0.914 | 0.42 | -0.27 | 1.10 | 0.898 |
| Pyruvate | 0.16 | 0.56 | -0.02 | 0.26 | -0.36 | 0.88 | 1.000 | 0.23 | -0.39 | 0.85 | 1.000 | 0.23 | -0.39 | 0.85 | 1.000 |
| Citrate | -0.17 | 0.82 | -0.84 | -0.01 | -0.53 | 0.52 | 1.000 | -0.05 | -0.58 | 0.49 | 1.000 | -0.05 | -0.58 | 0.49 | 1.000 |
| Glycerol | -0.02 | 0.75 | -0.11 | 0.09 | -0.51 | 0.69 | 1.000 | 0.13 | -0.47 | 0.73 | 1.000 | 0.13 | -0.48 | 0.73 | 1.000 |
| ***Ketone bodies*** | | | | | | | | | | | | | | | |
| 3-Hydroxybutyrate | 0.96 | 0.75 | -6.00 | -0.15 | -0.82 | 0.52 | 1.000 | -0.16 | -0.84 | 0.51 | 1.000 | -0.16 | -0.84 | 0.51 | 1.000 |
| Acetate | 0.03 | 0.20 | -0.33 | -0.17 | -0.87 | 0.52 | 1.000 | -0.18 | -0.87 | 0.51 | 1.000 | -0.18 | -0.87 | 0.51 | 1.000 |
| Acetoacetate | -0.05 | -0.27 | -0.34 | -0.03 | -0.74 | 0.68 | 1.000 | -0.05 | -0.77 | 0.68 | 1.000 | -0.05 | -0.77 | 0.67 | 1.000 |
| Acetone | -0.04 | 0.35 | -1.13 | -0.07 | -0.79 | 0.65 | 1.000 | -0.11 | -0.83 | 0.62 | 1.000 | -0.11 | -0.83 | 0.61 | 1.000 |
| ***Miscellaneous*** | | | | | | | | | | | | | | | |
| Creatinine | 0.03 | 0.46 | 0.05 | 0.00 | -0.41 | 0.41 | 1.000 | 0.00 | -0.42 | 0.41 | 1.000 | 0.00 | -0.41 | 0.41 | 1.000 |
| Albumin | 0.16 | 0.27 | 2.27 | -0.42 | -0.96 | 0.11 | 0.382 | -0.42 | -0.96 | 0.12 | 0.409 | -0.42 | -0.96 | 0.12 | 0.420 |
| Glycoprotein acetyls | 0.21 | 0.42 | -0.31 | 0.00 | -0.47 | 0.46 | 1.000 | 0.00 | -0.46 | 0.47 | 1.000 | 0.00 | -0.46 | 0.46 | 1.000 |
| ***Chylomicrons and extremely large VLDL*** | | | | | | | | | | | | | | | |
| Particles | 0.01 | -0.12 | -0.15 | -0.11 | -0.63 | 0.41 | 1.000 | -0.10 | -0.62 | 0.43 | 1.000 | -0.10 | -0.61 | 0.42 | 1.000 |
| Total lipids | -0.08 | -0.14 | -0.15 | -0.12 | -0.63 | 0.39 | 1.000 | -0.11 | -0.63 | 0.41 | 1.000 | -0.11 | -0.62 | 0.39 | 1.000 |
| Phospholipids | -0.08 | 0.12 | -0.12 | -0.13 | -0.67 | 0.41 | 1.000 | -0.12 | -0.67 | 0.42 | 1.000 | -0.12 | -0.66 | 0.41 | 1.000 |
| Cholesterol | -0.04 | 0.13 | 0.02 | -0.12 | -0.59 | 0.35 | 1.000 | -0.11 | -0.58 | 0.36 | 1.000 | -0.11 | -0.57 | 0.35 | 1.000 |
| Cholesteryl esters | -0.04 | 0.52 | 0.18 | -0.15 | -0.63 | 0.33 | 1.000 | -0.14 | -0.62 | 0.34 | 1.000 | -0.14 | -0.61 | 0.33 | 1.000 |
| Free cholesterol | -0.24 | 0.20 | -0.18 | 0.06 | -0.49 | 0.61 | 1.000 | 0.07 | -0.49 | 0.62 | 1.000 | 0.07 | -0.49 | 0.62 | 1.000 |
| Triglycerides | -0.31 | -0.62 | 0.08 | -0.23 | -0.86 | 0.41 | 1.000 | -0.23 | -0.87 | 0.40 | 1.000 | -0.23 | -0.86 | 0.39 | 1.000 |
| ***Very large VLDL*** | | | | | | | | | | | | | | | |
| Particles | -0.06 | 1.01 | 0.32 | -0.12 | -0.57 | 0.32 | 1.000 | -0.11 | -0.56 | 0.34 | 1.000 | -0.11 | -0.55 | 0.33 | 1.000 |
| Total lipids | -0.03 | 0.84 | 0.25 | -0.18 | -0.63 | 0.26 | 1.000 | -0.17 | -0.62 | 0.28 | 1.000 | -0.17 | -0.60 | 0.27 | 1.000 |
| Phospholipids | -0.09 | 1.13 | 0.39 | -0.18 | -0.62 | 0.26 | 1.000 | -0.17 | -0.61 | 0.27 | 1.000 | -0.17 | -0.60 | 0.26 | 1.000 |
| Cholesterol | -0.05 | 1.07 | 0.27 | -0.27 | -0.67 | 0.13 | 0.750 | -0.26 | -0.66 | 0.14 | 0.828 | -0.26 | -0.65 | 0.14 | 0.796 |
| Cholesteryl esters | -0.07 | 1.14 | 0.32 | -0.33 | -0.71 | 0.06 | 0.221 | -0.32 | -0.71 | 0.07 | 0.285 | -0.32 | -0.70 | 0.06 | 0.259 |

| | | | | Crude | | | | Adjusted | | | | Adjusted + body fat % | | | |
|---|---|---|---|---|---|---|---|---|---|---|---|---|---|---|---|
| | | | | | 99.95% CI | | | | 99.95% CI | | | | 99.95% CI | | |
| **Metabolite** | ***g*₁** | ***g*₂** | ***λ*** | **Est.** | **Lower** | **Upper** | ***P*** | **Est.** | **Lower** | **Upper** | ***P*** | **Est.** | **Lower** | **Upper** | ***P*** |
| ***Very large VLDL continue*** | | | | | | | | | | | | | | | |
| Free cholesterol | -0.06 | 1.05 | 0.32 | -0.18 | -0.61 | 0.25 | 1.000 | -0.17 | -0.61 | 0.26 | 1.000 | -0.17 | -0.59 | 0.25 | 1.000 |
| Triglycerides | -0.20 | 1.82 | 0.33 | -0.12 | -0.61 | 0.38 | 1.000 | -0.10 | -0.59 | 0.40 | 1.000 | -0.10 | -0.58 | 0.38 | 1.000 |
| ***Large VLDL*** | | | | | | | | | | | | | | | |
| Particles | -0.10 | 0.51 | 0.06 | -0.16 | -0.60 | 0.29 | 1.000 | -0.14 | -0.59 | 0.31 | 1.000 | -0.14 | -0.57 | 0.29 | 1.000 |
| Total lipids | -0.14 | 0.91 | 0.10 | -0.19 | -0.64 | 0.27 | 1.000 | -0.17 | -0.63 | 0.28 | 1.000 | -0.17 | -0.62 | 0.27 | 1.000 |
| Phospholipids | -0.05 | 1.52 | 0.42 | -0.16 | -0.60 | 0.28 | 1.000 | -0.14 | -0.59 | 0.30 | 1.000 | -0.14 | -0.57 | 0.29 | 1.000 |
| Cholesterol | -0.10 | 1.03 | 0.29 | -0.21 | -0.62 | 0.20 | 0.990 | -0.20 | -0.62 | 0.21 | 0.997 | -0.20 | -0.61 | 0.20 | 0.995 |
| Cholesteryl esters | -0.11 | 1.05 | 0.32 | -0.25 | -0.66 | 0.15 | 0.853 | -0.25 | -0.65 | 0.16 | 0.911 | -0.24 | -0.64 | 0.15 | 0.890 |
| Free cholesterol | -0.09 | 1.03 | 0.29 | -0.17 | -0.59 | 0.25 | 1.000 | -0.16 | -0.59 | 0.27 | 1.000 | -0.16 | -0.57 | 0.26 | 1.000 |
| Triglycerides | -0.16 | 1.01 | -0.04 | -0.21 | -0.69 | 0.28 | 1.000 | -0.19 | -0.68 | 0.30 | 1.000 | -0.19 | -0.66 | 0.28 | 1.000 |
| ***Medium VLDL*** | | | | | | | | | | | | | | | |
| Particles | -0.14 | 1.08 | -0.10 | -0.28 | -0.66 | 0.10 | 0.549 | -0.27 | -0.66 | 0.12 | 0.660 | -0.27 | -0.65 | 0.11 | 0.614 |
| Total lipids | -0.15 | 0.86 | -0.17 | -0.26 | -0.67 | 0.15 | 0.837 | -0.25 | -0.67 | 0.16 | 0.899 | -0.25 | -0.65 | 0.15 | 0.863 |
| Phospholipids | -0.13 | 1.20 | 0.00 | -0.30 | -0.68 | 0.09 | 0.419 | -0.29 | -0.67 | 0.10 | 0.533 | -0.28 | -0.66 | 0.09 | 0.488 |
| Cholesterol | -0.06 | 1.00 | 0.22 | **-0.41** | **-0.79** | **-0.03** | **0.020** | **-0.40** | **-0.78** | **-0.01** | **0.032** | **-0.40** | **-0.78** | **-0.02** | **0.031** |
| Cholesteryl esters | 0.00 | 0.75 | 0.33 | **-0.48** | **-0.89** | **-0.07** | **0.007** | **-0.47** | **-0.89** | **-0.05** | **0.012** | **-0.47** | **-0.88** | **-0.05** | **0.012** |
| Free cholesterol | -0.12 | 1.21 | 0.05 | -0.31 | -0.69 | 0.06 | 0.252 | -0.30 | -0.68 | 0.08 | 0.346 | -0.30 | -0.67 | 0.07 | 0.315 |
| Triglycerides | -0.16 | 0.78 | -0.23 | -0.15 | -0.63 | 0.32 | 1.000 | -0.14 | -0.62 | 0.34 | 1.000 | -0.14 | -0.61 | 0.32 | 1.000 |
| ***Small VLDL*** | | | | | | | | | | | | | | | |
| Particles | -0.12 | 0.58 | -0.25 | -0.12 | -0.53 | 0.29 | 1.000 | -0.11 | -0.52 | 0.30 | 1.000 | -0.11 | -0.52 | 0.30 | 1.000 |
| Total lipids | -0.12 | 0.66 | -0.22 | -0.14 | -0.55 | 0.27 | 1.000 | -0.13 | -0.54 | 0.29 | 1.000 | -0.12 | -0.53 | 0.28 | 1.000 |
| Phospholipids | -0.13 | 1.03 | -0.10 | -0.26 | -0.63 | 0.12 | 0.727 | -0.25 | -0.63 | 0.13 | 0.817 | -0.24 | -0.62 | 0.13 | 0.802 |
| Cholesterol | -0.16 | 1.18 | 0.03 | -0.31 | -0.68 | 0.07 | 0.299 | -0.29 | -0.67 | 0.08 | 0.389 | -0.29 | -0.67 | 0.08 | 0.387 |
| Cholesteryl esters | -0.18 | 1.30 | 0.10 | -0.29 | -0.67 | 0.09 | 0.439 | -0.28 | -0.66 | 0.10 | 0.537 | -0.28 | -0.66 | 0.10 | 0.542 |
| Free cholesterol | -0.12 | 0.96 | -0.01 | -0.32 | -0.69 | 0.05 | 0.179 | -0.31 | -0.69 | 0.06 | 0.253 | -0.31 | -0.68 | 0.06 | 0.243 |
| Triglycerides | -0.02 | 0.13 | -0.24 | 0.10 | -0.40 | 0.60 | 1.000 | 0.11 | -0.39 | 0.61 | 1.000 | 0.11 | -0.38 | 0.60 | 1.000 |
| ***Very small VLDL*** | | | | | | | | | | | | | | | |
| Particles | 0.00 | 0.92 | -0.30 | -0.01 | -0.41 | 0.38 | 1.000 | 0.00 | -0.40 | 0.39 | 1.000 | 0.00 | -0.39 | 0.39 | 1.000 |
| Total lipids | -0.08 | 0.87 | -0.31 | 0.03 | -0.38 | 0.44 | 1.000 | 0.04 | -0.36 | 0.45 | 1.000 | 0.04 | -0.36 | 0.45 | 1.000 |
| Phospholipids | -0.12 | 1.03 | -0.17 | 0.05 | -0.38 | 0.47 | 1.000 | 0.06 | -0.36 | 0.48 | 1.000 | 0.06 | -0.36 | 0.48 | 1.000 |
| Cholesterol | 0.10 | 0.98 | 0.11 | -0.10 | -0.49 | 0.30 | 1.000 | -0.08 | -0.48 | 0.31 | 1.000 | -0.08 | -0.48 | 0.31 | 1.000 |
| Cholesteryl esters | 0.14 | 1.00 | 0.23 | -0.13 | -0.53 | 0.28 | 1.000 | -0.11 | -0.51 | 0.30 | 1.000 | -0.11 | -0.51 | 0.30 | 1.000 |
| Free cholesterol | -0.02 | 1.04 | -0.16 | -0.04 | -0.44 | 0.36 | 1.000 | -0.03 | -0.42 | 0.36 | 1.000 | -0.03 | -0.42 | 0.36 | 1.000 |
| Triglycerides | -0.10 | 0.62 | -0.25 | 0.25 | -0.25 | 0.75 | 0.995 | 0.27 | -0.22 | 0.76 | 0.978 | 0.27 | -0.22 | 0.76 | 0.977 |
| ***IDL*** | | | | | | | | | | | | | | | |
| Particles | 0.20 | 0.80 | 0.29 | -0.24 | -0.66 | 0.19 | 0.975 | -0.21 | -0.64 | 0.22 | 0.996 | -0.21 | -0.64 | 0.22 | 0.996 |
| Total lipids | 0.16 | 0.79 | 0.31 | -0.14 | -0.56 | 0.29 | 1.000 | -0.11 | -0.54 | 0.31 | 1.000 | -0.11 | -0.54 | 0.31 | 1.000 |
| Phospholipids | 0.08 | 0.78 | 0.13 | -0.19 | -0.60 | 0.22 | 0.999 | -0.17 | -0.58 | 0.24 | 1.000 | -0.17 | -0.58 | 0.24 | 1.000 |
| Cholesterol | 0.19 | 1.50 | 0.43 | -0.17 | -0.62 | 0.28 | 1.000 | -0.15 | -0.60 | 0.31 | 1.000 | -0.14 | -0.60 | 0.31 | 1.000 |
| Cholesteryl esters | 0.18 | 1.32 | 0.45 | -0.15 | -0.61 | 0.32 | 1.000 | -0.12 | -0.59 | 0.35 | 1.000 | -0.12 | -0.59 | 0.34 | 1.000 |

| | | | | Crude | | | | Adjusted | | | | Adjusted + body fat % | | | |
|---|---|---|---|---|---|---|---|---|---|---|---|---|---|---|---|
| | | | | | 99.95% CI | | | | 99.95% CI | | | | 99.95% CI | | |
| **Metabolite** | $g_1$ | $g_2$ | $\lambda$ | **Est.** | **Lower** | **Upper** | $P$ | **Est.** | **Lower** | **Upper** | $P$ | **Est.** | **Lower** | **Upper** | $P$ |
| ***IDL continue*** | | | | | | | | | | | | | | | |
| Free cholesterol | 0.22 | 1.78 | 0.36 | -0.24 | -0.67 | 0.19 | 0.960 | -0.22 | -0.65 | 0.21 | 0.993 | -0.22 | -0.64 | 0.21 | 0.993 |
| Triglycerides | -0.12 | 0.47 | -0.82 | 0.29 | -0.19 | 0.78 | 0.895 | 0.32 | -0.15 | 0.79 | 0.736 | 0.32 | -0.15 | 0.79 | 0.740 |
| ***Large LDL*** | | | | | | | | | | | | | | | |
| Particles | 0.04 | 1.52 | -0.25 | **-0.38** | **-0.76** | **0.00** | **0.047** | -0.37 | -0.75 | 0.01 | 0.071 | -0.37 | -0.75 | 0.01 | 0.067 |
| Total lipids | 0.03 | 0.87 | 0.04 | -0.27 | -0.70 | 0.15 | 0.821 | -0.25 | -0.67 | 0.18 | 0.945 | -0.25 | -0.67 | 0.18 | 0.941 |
| Phospholipids | 0.04 | 0.78 | 0.07 | -0.33 | -0.76 | 0.10 | 0.441 | -0.30 | -0.73 | 0.13 | 0.661 | -0.30 | -0.73 | 0.13 | 0.653 |
| Cholesterol | 0.02 | 1.07 | 0.06 | -0.29 | -0.72 | 0.14 | 0.750 | -0.26 | -0.70 | 0.17 | 0.900 | -0.26 | -0.69 | 0.17 | 0.893 |
| Cholesteryl esters | 0.00 | 0.95 | 0.05 | -0.28 | -0.71 | 0.15 | 0.799 | -0.25 | -0.68 | 0.18 | 0.930 | -0.25 | -0.68 | 0.17 | 0.923 |
| Free cholesterol | 0.07 | 1.81 | 0.14 | -0.32 | -0.78 | 0.14 | 0.680 | -0.29 | -0.76 | 0.17 | 0.840 | -0.29 | -0.75 | 0.17 | 0.836 |
| Triglycerides | -0.08 | 0.44 | -0.91 | 0.26 | -0.19 | 0.72 | 0.941 | 0.29 | -0.15 | 0.73 | 0.781 | 0.29 | -0.15 | 0.73 | 0.782 |
| ***Medium LDL*** | | | | | | | | | | | | | | | |
| Particles | -0.09 | 0.86 | -0.07 | **-0.39** | **-0.74** | **-0.03** | **0.017** | **-0.37** | **-0.73** | **-0.01** | **0.039** | **-0.37** | **-0.72** | **-0.01** | **0.037** |
| Total lipids | -0.04 | 1.11 | 0.00 | -0.37 | -0.75 | 0.00 | 0.055 | -0.35 | -0.73 | 0.03 | 0.116 | -0.35 | -0.72 | 0.03 | 0.106 |
| Phospholipids | -0.02 | 0.87 | -0.01 | -0.35 | -0.74 | 0.04 | 0.157 | -0.33 | -0.72 | 0.07 | 0.287 | -0.33 | -0.72 | 0.07 | 0.272 |
| Cholesterol | -0.05 | 1.22 | 0.06 | **-0.41** | **-0.79** | **-0.03** | **0.021** | **-0.39** | **-0.77** | **0.00** | **0.047** | **-0.39** | **-0.76** | **-0.01** | **0.042** |
| Cholesteryl esters | -0.08 | 1.21 | 0.02 | **-0.40** | **-0.77** | **-0.03** | **0.018** | **-0.38** | **-0.75** | **-0.01** | **0.041** | **-0.38** | **-0.74** | **-0.01** | **0.036** |
| Free cholesterol | 0.03 | 1.26 | 0.13 | -0.42 | -0.86 | 0.03 | 0.103 | -0.39 | -0.84 | 0.06 | 0.179 | -0.39 | -0.84 | 0.05 | 0.173 |
| Triglycerides | -0.05 | 0.57 | -1.00 | 0.17 | -0.26 | 0.61 | 1.000 | 0.20 | -0.23 | 0.63 | 0.999 | 0.20 | -0.22 | 0.62 | 0.998 |
| ***Small LDL*** | | | | | | | | | | | | | | | |
| Particles | -0.01 | 0.96 | -0.09 | **-0.46** | **-0.84** | **-0.07** | **0.005** | **-0.44** | **-0.83** | **-0.05** | **0.011** | **-0.44** | **-0.82** | **-0.05** | **0.010** |
| Total lipids | 0.03 | 1.39 | -0.07 | **-0.42** | **-0.79** | **-0.06** | **0.007** | **-0.40** | **-0.77** | **-0.04** | **0.016** | **-0.40** | **-0.77** | **-0.04** | **0.015** |
| Phospholipids | 0.05 | 1.50 | 0.12 | **-0.41** | **-0.77** | **-0.05** | **0.008** | **-0.40** | **-0.76** | **-0.04** | **0.016** | **-0.40** | **-0.76** | **-0.04** | **0.015** |
| Cholesterol | 0.01 | 1.19 | -0.03 | **-0.46** | **-0.84** | **-0.08** | **0.005** | **-0.44** | **-0.83** | **-0.05** | **0.010** | **-0.44** | **-0.82** | **-0.05** | **0.009** |
| Cholesteryl esters | -0.01 | 1.31 | -0.14 | **-0.43** | **-0.81** | **-0.06** | **0.007** | **-0.41** | **-0.79** | **-0.04** | **0.015** | **-0.41** | **-0.78** | **-0.04** | **0.013** |
| Free cholesterol | 0.01 | 0.96 | 0.29 | **-0.49** | **-0.91** | **-0.06** | **0.009** | **-0.47** | **-0.90** | **-0.04** | **0.016** | **-0.47** | **-0.90** | **-0.04** | **0.015** |
| Triglycerides | -0.06 | 0.63 | -1.05 | 0.07 | -0.38 | 0.53 | 1.000 | 0.10 | -0.36 | 0.55 | 1.000 | 0.10 | -0.35 | 0.54 | 1.000 |
| ***Very large HDL*** | | | | | | | | | | | | | | | |
| Particles | -0.27 | 1.39 | 0.12 | 0.23 | -0.09 | 0.55 | 0.604 | 0.23 | -0.10 | 0.55 | 0.649 | 0.23 | -0.10 | 0.55 | 0.673 |
| Total lipids | -0.29 | 1.64 | 0.33 | 0.23 | -0.10 | 0.56 | 0.627 | 0.23 | -0.10 | 0.56 | 0.707 | 0.22 | -0.10 | 0.55 | 0.716 |
| Phospholipids | -0.23 | 0.73 | 0.49 | 0.28 | -0.04 | 0.60 | 0.197 | 0.27 | -0.05 | 0.59 | 0.257 | 0.27 | -0.05 | 0.59 | 0.255 |
| Cholesterol | -0.24 | 1.15 | 0.23 | 0.16 | -0.17 | 0.48 | 0.997 | 0.16 | -0.17 | 0.49 | 0.998 | 0.15 | -0.17 | 0.48 | 0.999 |
| Cholesteryl esters | -0.21 | 1.00 | 0.25 | 0.17 | -0.15 | 0.49 | 0.982 | 0.17 | -0.15 | 0.49 | 0.983 | 0.17 | -0.15 | 0.49 | 0.986 |
| Free cholesterol | -0.32 | 1.31 | 0.25 | 0.11 | -0.26 | 0.48 | 1.000 | 0.10 | -0.28 | 0.47 | 1.000 | 0.10 | -0.27 | 0.47 | 1.000 |
| Triglycerides | 0.08 | 0.53 | -0.37 | 0.45 | -0.06 | 0.97 | 0.169 | 0.46 | -0.05 | 0.97 | 0.147 | 0.46 | -0.05 | 0.97 | 0.148 |
| ***Large HDL*** | | | | | | | | | | | | | | | |
| Particles | -0.18 | 0.29 | 0.31 | **0.35** | **0.06** | **0.64** | **0.004** | **0.36** | **0.07** | **0.66** | **0.002** | **0.36** | **0.07** | **0.66** | **0.003** |
| Total lipids | -0.16 | 0.13 | 0.34 | **0.38** | **0.08** | **0.67** | **0.001** | **0.39** | **0.10** | **0.69** | **0.001** | **0.39** | **0.09** | **0.69** | **0.001** |
| Phospholipids | -0.17 | 0.13 | 0.36 | **0.42** | **0.12** | **0.73** | **0.000** | **0.44** | **0.13** | **0.74** | **0.000** | **0.44** | **0.13** | **0.74** | **0.000** |
| Cholesterol | -0.15 | 0.16 | 0.39 | **0.29** | **0.00** | **0.58** | **0.043** | **0.31** | **0.02** | **0.60** | **0.027** | **0.30** | **0.01** | **0.60** | **0.031** |
| Cholesteryl esters | -0.14 | 0.13 | 0.40 | 0.28 | -0.01 | 0.57 | 0.072 | **0.29** | **0.00** | **0.58** | **0.044** | 0.29 | 0.00 | 0.58 | 0.051 |

| | | | | Crude | | | | Adjusted | | | | Adjusted + body fat % | | | |
|---|---|---|---|---|---|---|---|---|---|---|---|---|---|---|---|
| | | | | | 99.95% CI | | | | 99.95% CI | | | | 99.95% CI | | |
| **Metabolite** | $g_1$ | $g_2$ | $\lambda$ | **Est.** | **Lower** | **Upper** | ***P*** | **Est.** | **Lower** | **Upper** | ***P*** | **Est.** | **Lower** | **Upper** | ***P*** |
| ***Large HDL continue*** | | | | | | | | | | | | | | | |
| Free cholesterol | -0.19 | 0.51 | 0.40 | **0.34** | **0.04** | **0.64** | **0.012** | **0.35** | **0.05** | **0.65** | **0.008** | **0.35** | **0.04** | **0.65** | **0.010** |
| Triglycerides | -0.12 | 0.42 | 0.01 | **0.67** | **0.13** | **1.22** | **0.003** | **0.68** | **0.14** | **1.21** | **0.002** | **0.68** | **0.14** | **1.21** | **0.002** |
| ***Medium HDL*** | | | | | | | | | | | | | | | |
| Particles | -0.31 | 0.71 | 0.42 | **0.54** | **0.10** | **0.97** | **0.003** | **0.58** | **0.15** | **1.01** | **0.001** | **0.58** | **0.14** | **1.01** | **0.001** |
| Total lipids | -0.31 | 0.78 | 0.40 | **0.59** | **0.12** | **1.05** | **0.002** | **0.63** | **0.17** | **1.08** | **0.000** | **0.63** | **0.17** | **1.09** | **0.000** |
| Phospholipids | -0.33 | 0.85 | 0.30 | **0.63** | **0.14** | **1.13** | **0.002** | **0.68** | **0.19** | **1.17** | **0.000** | **0.68** | **0.19** | **1.17** | **0.000** |
| Cholesterol | -0.24 | 0.63 | 0.52 | **0.46** | **0.06** | **0.87** | **0.010** | **0.50** | **0.10** | **0.91** | **0.002** | **0.50** | **0.10** | **0.91** | **0.003** |
| Cholesteryl esters | -0.21 | 0.70 | 0.59 | **0.44** | **0.04** | **0.85** | **0.014** | **0.49** | **0.09** | **0.88** | **0.004** | **0.48** | **0.08** | **0.89** | **0.004** |
| Free cholesterol | -0.34 | 0.69 | 0.35 | **0.52** | **0.09** | **0.95** | **0.003** | **0.56** | **0.14** | **0.98** | **0.001** | **0.56** | **0.13** | **0.98** | **0.001** |
| Triglycerides | 0.04 | 1.44 | 0.44 | 0.46 | -0.12 | 1.03 | 0.369 | 0.48 | -0.08 | 1.05 | 0.221 | 0.48 | -0.08 | 1.05 | 0.217 |
| ***Small HDL*** | | | | | | | | | | | | | | | |
| Particles | 0.00 | 0.23 | 0.10 | 0.07 | -0.45 | 0.59 | 1.000 | 0.12 | -0.39 | 0.64 | 1.000 | 0.12 | -0.39 | 0.63 | 1.000 |
| Total lipids | -0.05 | 0.37 | 0.12 | 0.27 | -0.27 | 0.82 | 0.994 | 0.33 | -0.20 | 0.87 | 0.878 | 0.33 | -0.20 | 0.86 | 0.870 |
| Phospholipids | -0.10 | 0.46 | 0.03 | 0.38 | -0.18 | 0.95 | 0.736 | 0.44 | -0.12 | 0.99 | 0.369 | 0.44 | -0.11 | 0.99 | 0.359 |
| Cholesterol | -0.02 | 0.29 | 0.10 | 0.07 | -0.47 | 0.61 | 1.000 | 0.13 | -0.40 | 0.66 | 1.000 | 0.13 | -0.40 | 0.66 | 1.000 |
| Cholesteryl esters | 0.02 | 0.45 | 0.35 | 0.00 | -0.54 | 0.54 | 1.000 | 0.05 | -0.48 | 0.59 | 1.000 | 0.05 | -0.48 | 0.58 | 1.000 |
| Free cholesterol | -0.12 | 0.59 | -0.11 | 0.30 | -0.23 | 0.84 | 0.960 | 0.35 | -0.17 | 0.87 | 0.742 | 0.35 | -0.17 | 0.87 | 0.734 |
| Triglycerides | 0.21 | 1.85 | 0.51 | 0.19 | -0.28 | 0.67 | 1.000 | 0.22 | -0.25 | 0.69 | 0.999 | 0.22 | -0.25 | 0.68 | 0.999 |

Note. K-effective test number: 51.507.
